# Assessing Genotype-Phenotype Correlations with Deep Learning in Colorectal Cancer: A Multi-Centric Study

**DOI:** 10.1101/2025.02.04.25321660

**Authors:** Marco Gustav, Marko van Treeck, Nic G. Reitsam, Zunamys I. Carrero, Chiara M. Loeffler, Asier Rabasco Meneghetti, Bruno Märkl, Lisa A. Boardman, Amy J. French, Ellen L. Goode, Andrea Gsur, Stefanie Brezina, Marc J. Gunter, Neil Murphy, Pia Hönscheid, Christian Sperling, Sebastian Foersch, Robert Steinfelder, Tabitha Harrison, Ulrike Peters, Amanda Phipps, Jakob Nikolas Kather

**Author notes:** Correspondence to: Jakob Nikolas Kather, MD, MSc Professor of Clinical Artificial Intelligence Else Kroener Fresenius Center for Digital Health Technical University Dresden Fetscherstrasse 74 01307 Dresden, Germany http://www.kather.ai.

## Abstract

**Background:** Deep Learning (DL) has emerged as a powerful tool to predict genetic biomarkers directly from digitized Hematoxylin and Eosin (H&E) slides in colorectal cancer (CRC). However, few studies have systematically investigated the predictability of biomarkers beyond routinely available alterations such as microsatellite instability (MSI), and *BRAF* and *KRAS* mutations.

**Methods:** Our primary dataset comprised H&E slides of CRC tumors across five cohorts totaling 1,376 patients who underwent comprehensive panel sequencing, with an additional 536 patients from two public datasets for validation. We developed a DL model using a single transformer model to predict multiple genetic alterations directly from the slides. The model’s performance was compared against conventional single-target models, and potential confounders were analyzed.

**Findings:** The multi-target model was able to predict numerous biomarkers from pathology slides, matching and partly exceeding single-target transformers. The Area Under the Receiver Operating Characteristic curve (AUROC, mean ± std) on the primary external validation cohorts was: *BRAF* (0·78 ± 0·01), hypermutation (0·88 ± 0·01), MSI (0·93 ± 0·01), *RNF43* (0·86 ± 0·01); this biomarker predictability was mirrored across metrics and co-occurrence analyses. However, biomarkers with high AUROCs largely correlated with MSI, with model predictions depending considerably on MSI-associated morphology upon pathological examination.

**Interpretation:** Our study demonstrates that multi-target transformers can predict the biomarker status for numerous genetic alterations in CRC directly from H&E slides. However, their pre-dictability is mainly associated with MSI phenotype, despite indications of slight biomarker-inherent contributions to a phenotype. Our findings underscore the need to analyze confounders in AI-based oncology biomarkers. To enable this, we developed a validated model applicable to other cancers and larger, diverse datasets.

**Funding:** The German Federal Ministry of Health, the Max-Eder-Programme of German Cancer Aid, the German Federal Ministry of Education and Research, the German Academic Exchange Service, and the EU.

## Introduction

Exome sequencing, including targeted panel sequencing, plays a key role in precision oncology of colorectal cancer (CRC)^1,2^ but remains inaccessible to many CRC patients worldwide due to the need for cost-intensive laboratory equipment and complex data analysis.^3^ In contrast, Hematoxylin and Eosin (H&E) stained histopathology slides are a standard diagnostic tool available for almost every cancer patient globally. The advent of deep learning (DL) has unlocked these slides as a quantifiable data resource. Recent studies have extensively suggested that DL can predict molecular biomarkers directly from digitized H&E slides, including microsatellite instability (MSI)^4–10^, hypermutation status, and gene mutations such as *TP53*, *BRAF*, and *KRAS.*^7,8,10–13^ When applied as pre-screening tools, such DL systems can streamline the diagnostic workflow^9,14^, identifying those cases that need further testing and ruling out others.^15^

Previous DL studies in CRC have predominantly focused on specific genetic alterations as potential biomarkers, referred to as ‘prediction targets’ in computational pathology, with only few studies adopting a pan-molecular alteration approach.^11,16^ Such efforts have often been constrained by the challenge of obtaining diverse datasets with extensive sequencing, and even then, traditional methods require training separate models for individual prediction targets^5–8,12,13^, making them labor- and resource-intensive. Addressing both challenges, this study introduces the first pan-biomarker DL approach for CRC, utilizing a single model to predict multiple molecular targets. The transformer-based model was trained and validated on a unified dataset from the Genetics and Epidemiology of Colorectal Cancer Consortium (GECCO), which consolidates sequencing data from diverse cohorts.^17^ Furthermore, its generalizability was evaluated using two extensively studied CRC cohorts.

As DL has demonstrated robust capability in linking phenotype to genotype, particularly for pre-dicting MSI^5–9^, we leverage the strengths of our dataset and model to extend this analysis to multiple prediction targets. Specifically, we analyzed the co-occurrence of genetic alterations with MSI, evaluated the ability of DL to predict these alterations, and examined the corresponding slide morphology, relating it to features known to be relevant for MSI detection. Our study included genetic alterations investigated in prior DL-based studies and clinically relevant mutations, such as *BMPR2*, *RNF43* and *BRAF* as well as MSI and hypermutation status.

## Materials and Methods

### Patient Samples

GECCO is an international collaboration utilizing genotype and sequencing data from a growing resource of over 60 studies.^17^ Our experiments were conducted on five cohorts within GECCO, comprising N=1,376 patients with complete data and digitized whole-slide images (WSIs): European Prospective Investigation into Cancer (EPIC, N=183)^18^, Colorectal Cancer Study of Austria (CORSA, N=158)^19^, Iowa Women’s Health Study (IWHS, N=390)^20^, Cancer Risk Assessment study (CRA, N=321) and Women’s Health Initiative (WHI, N=324)^21^ (Fig. 1A-C). These studies provided digitized WSIs with unified demographic, clinical, and lifestyle data (relevant data for this study in Tab. S1), establishing them as the primary dataset for this study. Centralized targeted tumor sequencing of up to 356 genes was conducted on all samples, with MSI and hypermutation status also assessed. The analysis targeted non-silent mutations and mutational signatures using panel sequencing of 1·8 Megabases (Mb), with tumor and normal sequencing coverage of 975× and 273×, respectively. To ensure generalizability of our model and comparability with existing literature, we utilized a secondary external dataset with two well-characterized publicly available CRC cohorts from The Cancer Genome Atlas (TCGA, N=426)^22^ and the Clinical Proteomic Tumor Analysis Consortium (CPTAC, N=110, fresh-frozen tissue samples)^23^ (Fig. 1C). We included patients with WSIs and ground truth data on the primary genetic alterations analyzed in this study. The MSI status for TCGA was defined per Liu et al.^24^, with high grade MSI categorized as MSI and low grade MSI and microsatellite stable (MSS) grouped as MSS^25^. Ground truth labels were derived from the clinical data accessible at https://portal.gdc.cancer.gov/ (accessed on Nov 20, 2024).

**Fig. 1:**
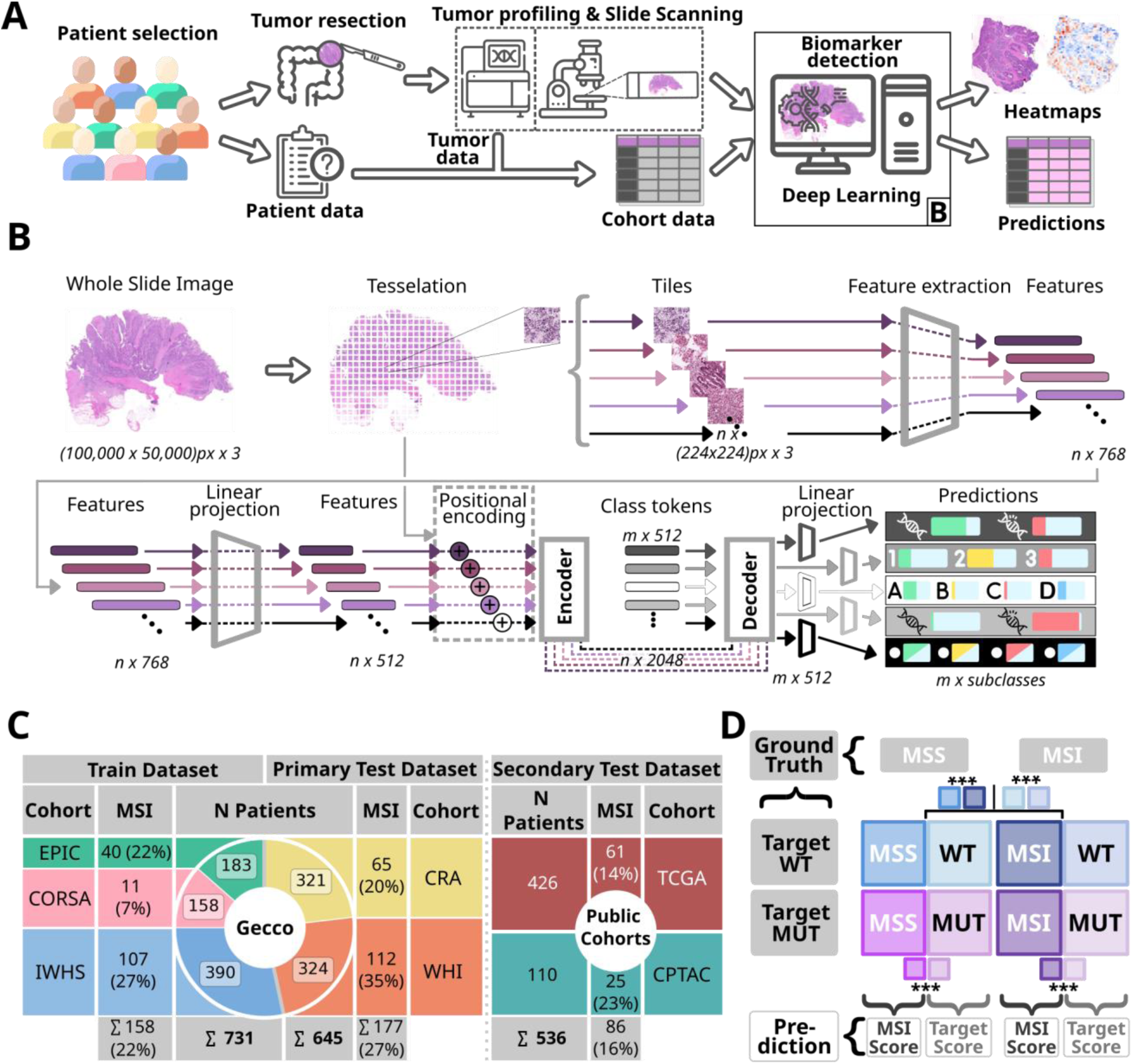
Experimental design, cohort characterization, and schematic for predictive analysis. **A**. Tissue samples from colorectal cancer (CRC) patients across five independent cohorts were obtained via surgical resection, with associated demographic, clinical, and sequencing data collected. Upon Hematoxylin and Eosin (H&E) staining, tumor tissues are digitized into Whole Slide Images (WSIs) for profiling genetic alterations. The WSIs are then used to train and test a deep learning (DL) algorithm for biomarker detection, to simultaneously predict multiple mutational statuses and provide heatmap explanations. **B**. The DL pipeline tessellates the WSIs into smaller tiles while rejecting background and blurry areas, extracting n feature vectors from n tiles. Feature vectors are compressed and processed in a multi-target transformer, employing an attention mechanism in an encoder-decoder structure for class token learning. The transformer generates individual scores for the respective amount of classes per target. The code is able to comprise positional tile embedding (dashed lines), which did not result in improved performance and were therefore excluded from our study. **C**. Overview of the five GECCO and two public cohorts, including patient numbers, slides, extracted features, and MSI case proportions. The cohorts are divided into train datasets and test datasets. **D**. Schematic for interpreting result plots and statistics, delineating dataset partitioning based on microsatellite (MSS: microsatellite stability, MSI: microsatellite instability) and gene mutational status (MUT: mutated, WT: wild type). The diagram illustrates distinct groups by color, with the left side representing MSI prediction scores and the right side for prediction target scores. True ground truth labels of samples guide the group organization, with model-generated scores depicted in corresponding colors.

### Experimental Design

Our study included genetic alterations investigated in prior DL-based studies, clinically relevant mutations, and genetic alterations strongly associated with MSI in the TCGA CRC cohort^26^, including *APC*, *BMPR2*, *BRAF*, *KRAS*, *RNF43*, *TP53*, *ZNRF3*.^7,8,11–13,26,27^ Together with MSI status and hypermutation, these exemplify the main prediction targets among the broad target set utilized from the GECCO cohorts (Tab. S2). Continuous prediction targets were discretized into classes using predefined thresholds to achieve balanced case distribution. Only prediction targets with at least 20 samples per class were included in model training to ensure robust analysis.

Model training involved 731 patients from three GECCO cohorts (EPIC, CORSA, IWHS; Fig. 1C) using seven-fold cross-validation to balance training and validation sets, ensure class distribution, and identify a median-performing model. The resulting seven models were deployed on external test sets. The primary test set consisted of 645 patients from two GECCO cohorts (CRA, WHI; Fig. 1C), ensuring a diverse and representative split while addressing missing cases (Tab. S1). Exclusively female cohorts were included in both datasets (IWHS for training, WHI for testing). To assess generalizability and enable comparison to the literature, selected models were validated on a secondary test set comprising TCGA and CPTAC cohorts. ‘External validation’ and ‘test set’ typically refer to the primary test set unless stated otherwise; cohort-wise analyses are explicitly noted. In total, we trained eleven models using the same training set: a primary model incorporating the broad target set (Tab. S2) and a secondary model excluding MSI as a target to evaluate its impact on other predictions—both tested on the primary and secondary test sets. Additionally, we trained nine single-target models, each dedicated to one of the main prediction targets, and evaluated them on the primary test set. All models were evaluated in direct comparison to the primary multi-target model and compared to the literature.

### Data Analysis of Genetic Alterations

In CRC, there is a pronounced co-occurrence of genetic alterations with MSI.^26,27^ We employed two distinct methods to investigate the interplay and dependencies between various genetic alterations within the five GECCO cohorts: First, we used hierarchical clustering to group molecular alterations in our dataset using the ‘Euclidean’ metric and ‘Ward’ procedure ^28^, using only alterations with complete data (Tab. S1). Subsequently, we applied association rule mining^29^ to reveal the co-occurrence of gene mutations and conditions such as MSI, considering an initiating genetic alteration (’antecedent’) and a potentially resulting alteration (’consequent’). The presence of an initiating alteration statistically increases the probability of observing a resulting alteration, without implying a cause-effect biological sequence. We used six complementary metrics to assess co-occurrence: ‘Support’, ‘Confidence’, ‘Lift’, ‘Leverage’, ‘Conviction’, ‘Zhang’s Metric’.

### Image Processing and Deep Learning Techniques

According to the framework of Wagner et al.^25^, the digitized WSIs were tessellated into tiles of 224×224 pixels, corresponding to 256×256 micrometers (Fig. 1B). Tiles predominantly containing background (brightness value ≥ 224) and blurred regions, identified using Canny edge detection^30^ (thresholds: 40, 100) with fewer than two percent of the tile’s pixels identified as edges were excluded. We used the pre-trained CTransPath feature extractor^25,31^ to extract a 768-dimensional feature representation for each tile, which we used as an input for a subsequent transformer model (Fig. 1B). Advancing upon previous work^25^ we developed a model with an encoder-decoder architecture to simultaneously predict multiple targets from tile embeddings (Fig. 1B). Specifically, the 768-dimensional tile features are projected into a 512-dimensional space using a fully connected layer to reduce complexity and enhance computational efficiency for the subsequent model. The encoded tokens are decoded in class tokens^32^, each with a dimension of 1×512, with one token dedicated for each prediction target. The decoded tokens are fed through a fully connected layer to generate target-specific predictions with scores ranging from zero (negative prediction) to one (positive prediction) for each class and patient. To address class imbalance during training, cross-entropy loss is calculated per target, weighted by the inverse mutation 5 frequency, and summed to ensure proportional importance of rare mutations. The training and deployment procedures were performed on a NVIDIA RTX A6000 featuring 48 GB of GPU memory. All source codes for preprocessing and DL are available in our open source repositories (Tab. S3). TRIPOD reporting guidelines were used.

### Explainability

To investigate the model’s detection of relevant regions for predicting alterations in CRC, we generated heatmaps using Grad-CAM^33^, focusing on the fold with the median AUROC for MSI detection to ensure consistent comparisons. These heatmaps highlight the contribution of each tile to patient-level predictions, enabling analysis of whether the model relies on distinct patterns and areas in WSIs for different targets or similar features across targets. Notably, a high number of positively contributing tiles does not necessarily correspond to a high final score due to the nonlinear aggregation process, and the model may also incorporate global cues not captured by the heatmap. To examine morphology in greater detail, we identified highly predictive tiles (‘top tiles’) for representative cases, selected based on their scores and attention values assigned by the model. For direct comparability, we included top tiles for each prediction target alongside those for MSI from the same slide. As an additional mode of explainability, we analyzed class token interactions in the decoder during deployment of the primary model to assess overlap for the main prediction targets. Grad-CAM was used to compute activations, capturing class token-score interactions, which were aggregated into a cross-correlation matrix, averaged across patients, and visualized as a heatmap.

### Statistical Analysis

We used descriptive statistics to summarize sociodemographic and clinicopathologic characteristics across cohorts (Tab. S1). Model performance was evaluated using AUROC during training and testing, with additional metrics and target-specific prediction scores (ranging from 0·00 to 1·00) analyzed. For the main prediction targets we applied a two-sided DeLong test to compare AUROCs of single-target models and the primary multi-target model on the primary external dataset, utilizing mean prediction scores across seven folds per target. Mean and median AUROC values for each target were calculated over the seven folds. DeLong tests were also applied to compare the primary model (including MSI as a target) with the secondary model (excluding MSI) within each of the four external cohorts, based on mean prediction scores from the seven models.

Given the rarity of mutations and the limitations of metrics such as AUROC and the Area Under the Precision-Recall Curve (AUPRC) in capturing imbalanced distributions^34^, we employed diverse metrics and analyzed individual prediction scores for the main prediction targets, stratified by MSI status. Average prediction scores across seven folds were calculated per patient and categorized into four subgroups per target based on microsatellite and mutational status: MSS/WT and MSS/MUT represent MSS tumors with wild-type (WT) and mutated (MUT) targets, respectively, while MSI/WT and MSI/MUT represent MSI tumors with wild-type and mutated targets (Fig. 1D). Model mutation detection performance was evaluated through comparisons on training and testing sets. After testing for normality with the Shapiro-Wilk test, the Mann-Whitney U test was used to assess statistical significance between MSI and prediction target scores within each subgroup. Additionally, four comparisons using the Wilcoxon test evaluated the model’s ability to identify prediction target mutations by comparing score distributions for MSS/WT vs MSS/MUT and MSI/WT vs MSI/MUT. Finally, subgroup comparisons based on prediction target status were performed: wild type (MSS/WT vs MSI/WT) or mutated (MSS/MUT vs MSI/MUT).

## Results

### Multi-Target Transformers Match and Partly Exceed Single-Target Models While Enabling Simultaneous Prediction of Genetic Alterations

Our approach aimed to reproduce prior DL-studies predicting genetic alterations in CRC from H&E slides^5–9,11–13,35,36^, expanding the scope to a broader set of alterations using a multi-target transformer architecture (Fig. 1B). We compared the performance of our transformer for selected targets including the main prediction targets (Fig. 2) with the external validation AUROCs reported in the literature for single-target models, which used varying model architectures.

**Fig. 2:**
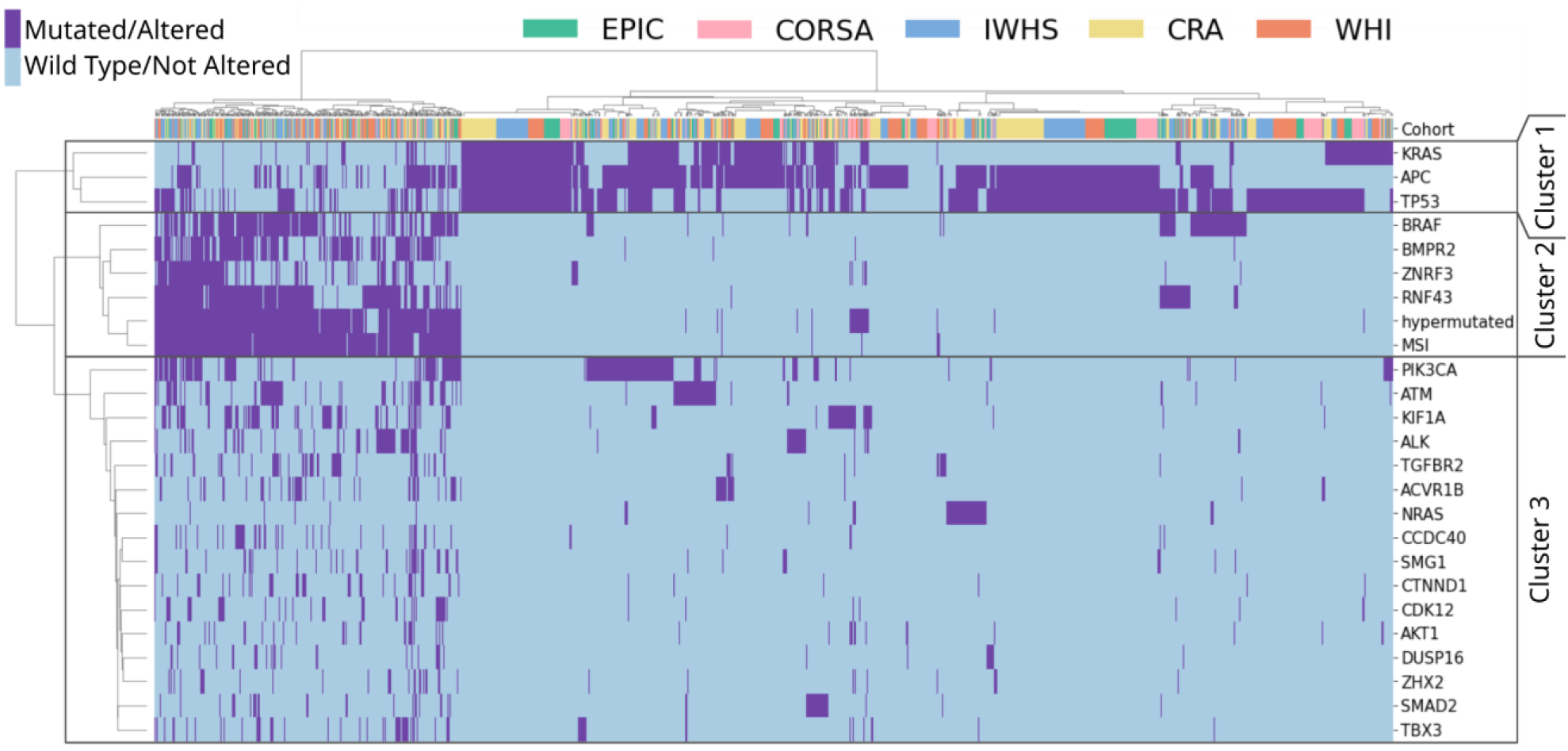
Analysis of genetic alterations co-occurrence in CRC for GECCO cohorts. Hierarchical clustering analysis was conducted on the ground truth of genetic alterations with fully available mutational information. Each row corresponds to a genetic alteration, and each column represents a patient from the dataset. The top row indicates the distribution of patients from various cohorts within genetic clusters. The distance calculation was performed using the ‘Euclidean’ metric, and the ‘Ward’ method was applied to clustering. Three unique genetic clusters were created and marked. The patient clustering shows a diverse distribution of samples across all five cohorts and genetic clusters (top row).

For MSI detection, single- and multi-target transformers achieved AUROCs of 0·91 (±0·02) and 0·93 (±0·01), respectively, on the primary test set (p<0·05; refer to Tab. S4, Fig. 3A here and in the following). The primary model achieved AUROCs from 0·87 (±0·01, TCGA) to 0·94 (±0·01, WHI) (Tab. S5-S8, Fig. S1; referred to here and in the following), consistent with the reported literature range of 0·77–0·96.^4,7,8,10,12,13,25,36^ The following summarizes model performance for external validation for selected targets, with AUROCs for single- and multi-target transformers (Tab. S4, Fig. 3A) and comparisons between primary (multi-target with MSI) and secondary (multi-target without MSI) models (Tab. S5-S12, Fig. S2). In *BRAF* MUT detection, the the AUROC was 0·72 (±0·06, single-target) and 0·78 (±0·01, multi-target, p<0·05). The AUROCs for primary vs. secondary models were lowest on WHI (0·77 (±0·01) vs. 0·76 (±0·01, p>0·05)) to highest on CRA (0·83 (±0·02) vs. 0·82 (±0·03, p>0·05)). These results align with the literature range of 0·66 to 0·88.^7,8,10–13,16,25,36^ In *RNF43* MUT detection, the AUROC was 0·80 (±0·05, single-target) and 0·86 (±0·01, multi-target, p<0·05). The AUROCs for primary vs. secondary models ranged from 0·80 (±0·01) vs. 0·79 (±0·01, p>0·05) on TCGA to 0·87 (±0·02) vs. 0·85 (±0·01, p>0·05) on CRA. These results exceed the literature range of 0·63 to 0·72.^11,16^ For KRAS MUT detection, the AUROC was 0·65 (±0·02) for both single- and multi-target models (p>0·05). The primary vs. secondary models ranged from 0·56 (±0·02) vs. 0·55 (±0·02, TCGA, p>0·05) to 0·69 (±0·06) vs. 0·72 (±0·03, CPTAC, p>0·05). These results fall below and within the literature range of 0·60 to 0·80.^8,11,12,16,25,36^ Interestingly, CPTAC had no *KRAS* MUT cases with MSI, while TCGA showed the highest number of such cases among all cohorts, with up to 19 instances. For hypermutation and *TP53* MUT detection, the results fell within the literature ranges of 0·81–0·87^8,10,13^ and 0·60–0·75^8,10,11,16^, with no significant differences between models. For *APC* mutational status prediction, the results ranged both above and below the single available literature AUROC of 0·67^11,16^, with no statistical differences between models, highlighting the limited number of comparable values. For *BMPR2* and *ZNRF43* there was no comparable data in the literature (AUROCs shown in Tab. S4, Fig. 3A, Fig. S2). Additional prediction targets with AUROCs ≥0·75 are detailed in Tab. S4.

**Fig. 3:**
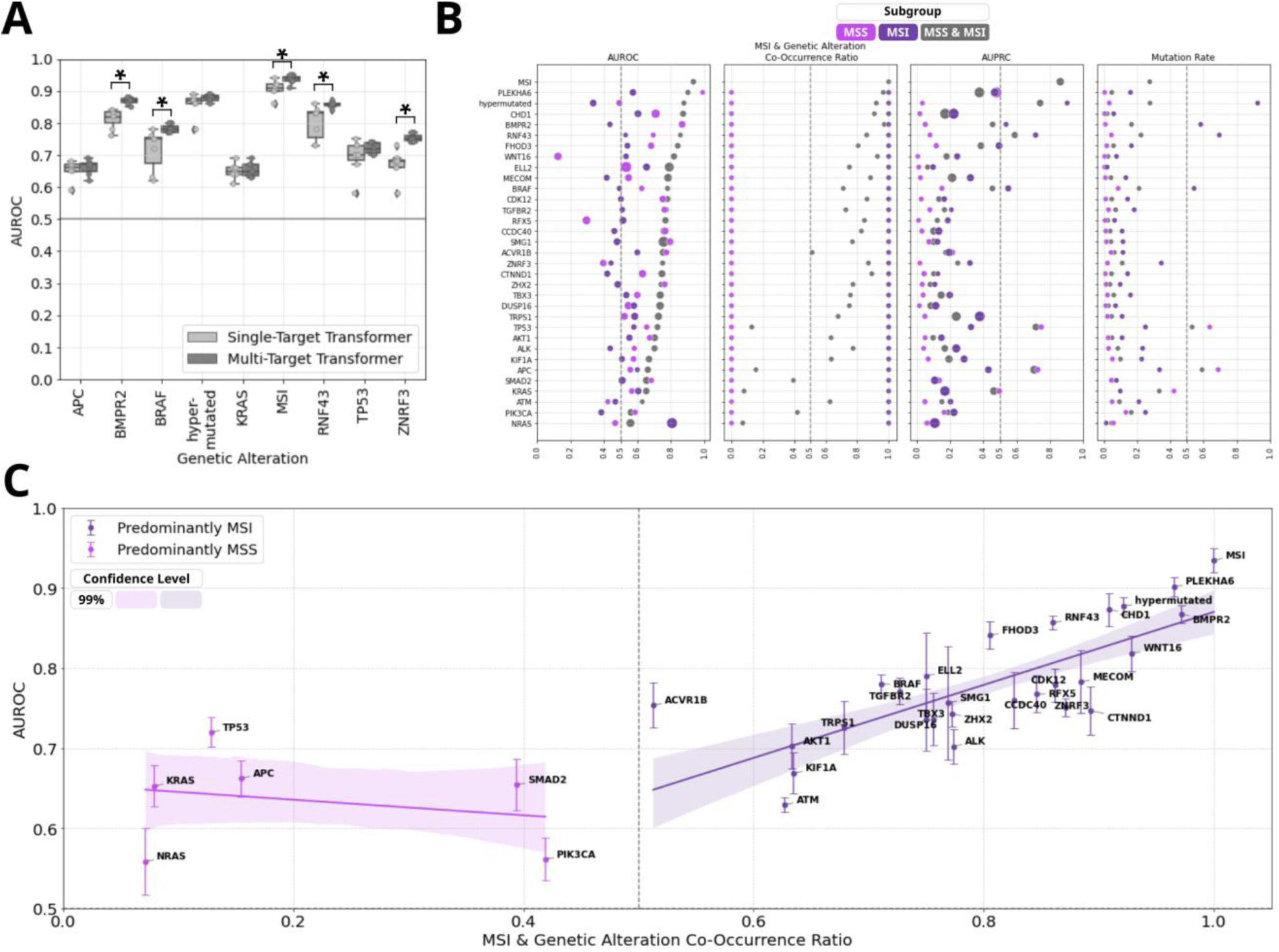
Evaluation of the performance of the Multi-Target Transformer on selected prediction targets for the external cohorts from GECCO. **A.** The comparison of Single-Target Transformer versus Multi-Target Transformers shows the Area Under the Receiver Operating Characteristic curve (AUROC) from each of the 7 folds of external cross-validation, with the median value highlighted with a horizontal line in each box. The figure includes selected representative potential biomarkers of genetic alterations associated with MSS (Fig. 2, genetic Cluster 1) and MSI (Fig. 2, genetic Cluster 2). The test set cohorts consist of CRA and WHI (Fig. 1C). The horizontal line positioned at an AUROC of 0·50 represents a random guess of the model. Significance was determined through a two-sided DeLong test with a p-value threshold of less than 0·05. **B.** Performance metrics of Multi-Target Transformers for external validation. The mean (center of dot) and standard deviation (diameter of dot) for relevant selected prediction targets for the whole external set, as well as the MSI and MSS subgroups, are displayed based on the 7 folds of cross-validation. The threshold for binary classification is pre-defined as 0·50· The evaluation metrics include the Area Under the Receiver Operating Characteristic Curve (AUROC), and the Area Under the Precision-Recall Curve (AUPRC), along with the corresponding mutation rates in external cohorts. The Mutation Rate refers to the fraction of instances with a specific mutation in the subgroup. The MSI & Genetic Alteration Co-Occurrence Ratio is the fraction of cases harboring MSI among all cases with a particular genetic mutation. The data is sorted for AUROC and shown in Tab. S14 and Tab. S17-S18. An extended version of this panel with more metrics is shown in Fig. S3B. **C.** Distribution of Areas under the Receiver Operating Characteristic Curve (AUROCs, mean ± standard deviation) for selected prediction targets and their co-occurrence with MSI with corresponding values and further metrics shown in Tab. S14. An extended version of this panel with MSS/MSI-subgroup specific AUROCs is shown in Fig. S3C.

### Enhanced Predictive Performance for Genetic Alterations Associated with MSI

Hierarchical clustering identified two primary genetic clusters: Cluster 2, comprising MSI-associated genes (e.g., *BRAF*, *BMPR2*, *ZNRF3*, *RNF43*) with strong co-occurrence with MSI and hypermutation, and Cluster 1, including MSS-associated genes such as *TP53*, *KRAS*, and *APC* (Fig. 2). Association rule mining reinforced robust MSI correlations in Cluster 2 (e.g., *BMPR2*, *RNF43*), contrasting with inverse relationships in Cluster 1 MSS-linked genes (e.g., *TP53*, *KRAS*) (Fig. S3A, Tab. S13).

MSI-associated alterations showed higher AUROC values for predicting genetic mutations compared to MSS-associated alterations (Cluster 1) (Fig. 3B-C, Tab. S14, Fig. S1). Additional metrics (Fig. 3B, Fig. S3B, Tab. S14) and patient-level prediction score distributions were analyzed for MSI and genetic alterations within Clusters 1 and 2 (Fig. 1D, Fig. 4, Tab. S15-S16). Prediction scores quantified model certainty, with high scores indicating mutation presence, low scores indicating absence, and scores near 0·5 reflecting indecision. Comprehensive summaries supported internal (Fig. S4) and external validation (Fig. S5). For MSS-associated Cluster 1 genes (*TP53*, *APC*, *KRAS*) the external validation AUROCs ranged from 0·65 to 0·72, with MSI scores accurately reflecting ground truth: low for MSS and high for MSI cases (Fig. 2, Fig. 4A, Tab. S4). High MSI scores tended to align with WT target predictions, while low scores aligned with MUT predictions. As a result, discrepancies were observed in MSS/WT and MSI/MUT subgroups, where target predictions deviated from ground truth. Genetic Cluster 2, including MSI, hypermutation, and the genes *BMPR2*, *ZNRF3*, *RNF43*, and *BRAF*, demonstrated higher AUROCs (0·75–0·88) during external validation (Fig. 2, Fig. 4B, Tab. S4). In MSS patients, mutations in biomarkers like BMPR2 (n=3) and ZNRF3 (n=9) were rare, and MSS/MUT subgroups showed less distinct prediction scores for RNF43 (n=20) and BRAF (n=39), reflecting higher prediction uncertainty. MSI scores aligned with the ground truth (MSS/MSI), but unlike Cluster 1, alteration scores in all subgroups correlated with MSI scores. This produced accurate prediction trends for MSS/WT and MSI/MUT subgroups (low scores for WT, high for MUT), while MSS/MUT and MSI/WT subgroups showed deviations from the target ground truth. These findings, partially reflected in AUROC results (Fig. 3B, Fig. S3B), suggest less effective differentiation between MUT and WT in MSS and MSI subgroups compared to the combined group. Despite the strong influence of MSI-associated morphology on predictions, aligning the prediction target scores with MSS (Cluster 1) or MSI-high (Cluster 2), intrinsic phenotypes for some alterations cannot be fully confirmed or excluded. Analyses of MSI-high and MSS subgroups (Tab. S17-S18, Fig. 3B, Fig. S3B-S3C) revealed AUROCs of 0·60–0·70 for genes like *BRAF*, *RNF43*, and *TP53* in MSS patients, indicating modest discrimination between MUT and WT. Statistical differences in score distributions for MSI and these genetic alterations further supported these findings (Fig. 4A-B).

**Fig. 4:**
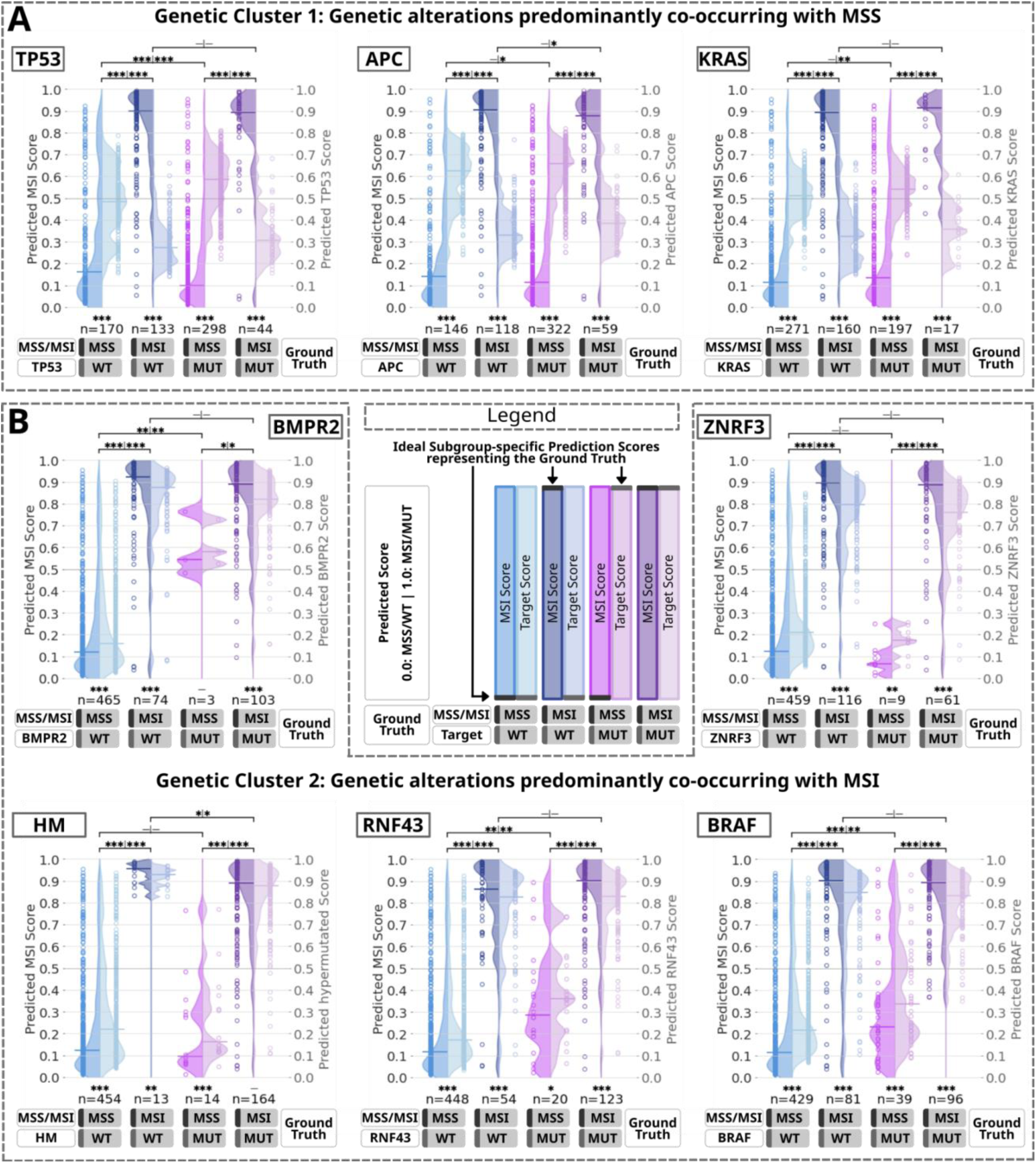
Evaluation of prediction scores based on the multi-target transformer in external validation on the GECCO test set subgrouped by the co-occurrence of the prediction targets with MSI. A.-B. Violin plots representing individual patient scores from the test set cohorts for MSI and representative genetic alterations in four subgroups based on microsatellite and alteration mutational status. The left y-axis represents the MSI score scale (left violin halfs) and the right y-axis corresponds to the prediction target scores (right violin halfs). The legend displays gray horizontal lines in the concept violins that represent the optimal position of the prediction scores based on ground truth. The selection of prediction targets includes *TP53*, *APC*, and *KRAS* from genetic Cluster 1 (**A**.), and *BMPR2*, *ZNRF3*, Hypermutation (HM), *RNF43*, and *BRAF* from genetic Cluster 2 (**B**.) (Fig. 2). The data encompasses both external cohorts CRA and WHI (Fig. 1C). Each dot represents the mean value of individual patient prediction scores calculated from 7 folds, with the horizontal line on each side of the violin indicating the median of all individual mean patient scores. A horizontal line at 0·50 denotes the line of model uncertainty. The sample count for each subgroup is indicated below the violins. Statistical significance is denoted in the figures as follows: * for p < 0·05, ** for p < 0·01, *** for p < 0·001, with more details provided in Fig. 1D. After testing for normal distribution (Tab. S20), the Mann-Whitney U test was used for within-group comparisons, and the Wilcoxon test was used for between-group comparisons. Abbreviations: HM: Hypermutation; MSI: Microsatellite instability; MSS: Microsatellite stability; MUT: Mutated; WT: Wild type.

### MSI-Associated Morphological Patterns Drive Predictions Across Prediction Targets

To investigate the morphological patterns underlying the prediction of genetic alterations in CRC from H&E slides using DL, we analyzed WSI heatmaps (Fig. 5, Fig. S6-S10) and highly predictive tiles (‘top tiles’, Fig. 6, Fig. S11-S23) for key prediction targets (Cluster 1: *TP53*, *APC*, *KRAS*; Cluster 2: *RNF43*, *BRAF*, hypermutated) in 25 cases from CRA and WHI test cohorts. This was assessed via manual histopathological review of WSI heatmaps and 20 top tiles per target (Tab. S19). We found that our model predominantly focused on tumor regions, with minimal relevance attributed to pen marks or non-tumor areas (e.g., Fig. 5A-D, Fig. S6B-D, Fig. S7A-D). Pen marks, present on most slides, were rarely highlighted and appeared faintly in top tiles (e.g., Fig. S13, Fig. S22), though slight highlighting in heatmaps occurred in rare cases (e.g., Fig. 5C, Fig. S9D). Regions of high model attention primarily consisted of tumor tissue, not normal intestinal mucosa or uninvolved connective/adipose tissue, despite the model not being explicitly trained for tumor detection.

**Fig. 5:**
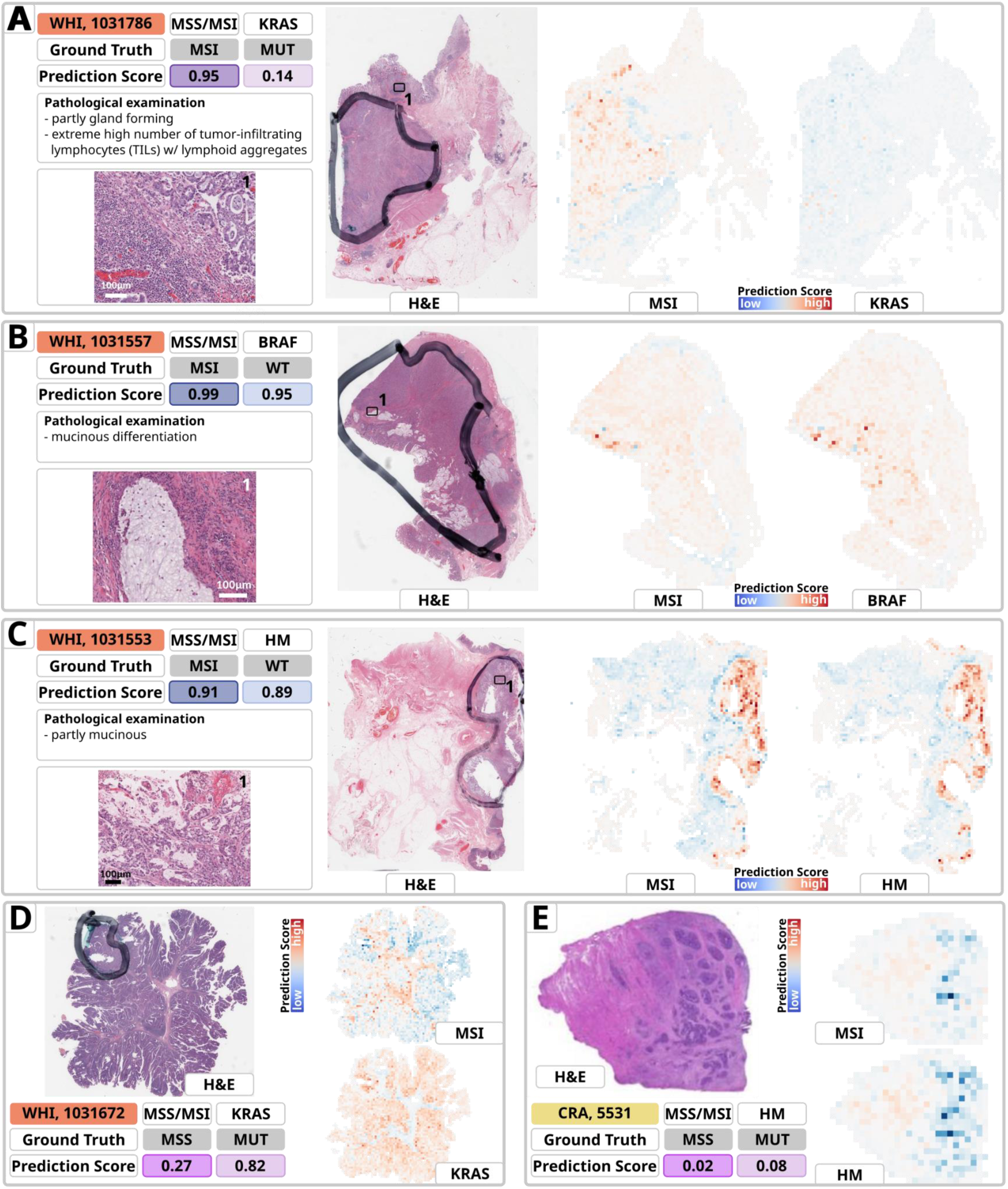
Heatmaps of representative samples for prediction of MSI, *KRAS, BRAF* and Hypermutation (HM) from the external GECCO validation dataset. The heatmaps are derived from the model with the median AUROC for MSI detection and the majority of prediction targets evaluated by sevenfold cross-validation. The cohort, Sample-ID, ground truth and prediction scores for MSI, along with the individual mutational status of the target, a brief pathological evaluation and magnified views of specific areas are provided for in-depth analysis. The heatmaps indicate relevant areas for the various predictions. The red areas are of high importance and indicate a mutant type (MUT), while the blue areas are of low importance and indicate a wild type (WT). The color intensity showcases the model’s attention to that distinct area. **A.** The tumor exhibits both gland-forming and more solid components and extremely high numbers of tumor-infiltrating lymphocytes (TILs) with dense lymphoid aggregates. The pathological examination confirms the plausibility of a high MSI score indicating MSI which is also the ground truth. A low *KRAS* score indicates *KRAS* WT but the ground truth is *KRAS* MUT. The heatmap highlights similar tumor areas but with diverging scores: where MSI map is red indicating high score, *KRAS* map is blue indicating low score. **B.** The presence of mucinous differentiation in a MSI, *BRAF* WT case results in high MSI and *BRAF* scores. The MSI score is pathologically plausible whereas the *BRAF* score indicates a contrary prediction tendency than the ground truth holds. For both predictions, the model focuses on similar tumor areas with similar scores indicating MSI/*BRAF* MUT. **C.** Partly mucinous morphology indicates the possibility of MSI, with a high score predicting MSI. HM is also predicted MUT with a high score, even though HM is WT for this sample. Both heatmaps primarily label the tumor and the same region with comparable significance. **D.** Villous adenoma with high grade dysplasia is a common precursor lesion associated with high frequency of *KRAS* mutations ^41,42^ The heatmaps highlight similar large scale tumor areas but with converging scores: where the MSI map is red indicating a high score, the *KRAS* map is blue indicating low score. **E**. The tumor area appears to be mainly MSS, and the heatmap predicts a low score, indicating this. Although it is being mutated in the ground truth, it is still predicted as non-hypermutated. This is a rare MSS case with HM ^54^ Both heatmaps predominantly mark the tumor area and the same region with comparable relevance. Abbreviations: HM: Hypermutation; MSI: Microsatellite instability; MSS: Microsatellite stability; MUT: Mutated; WT: Wild type; w/: with

**Fig. 6:**
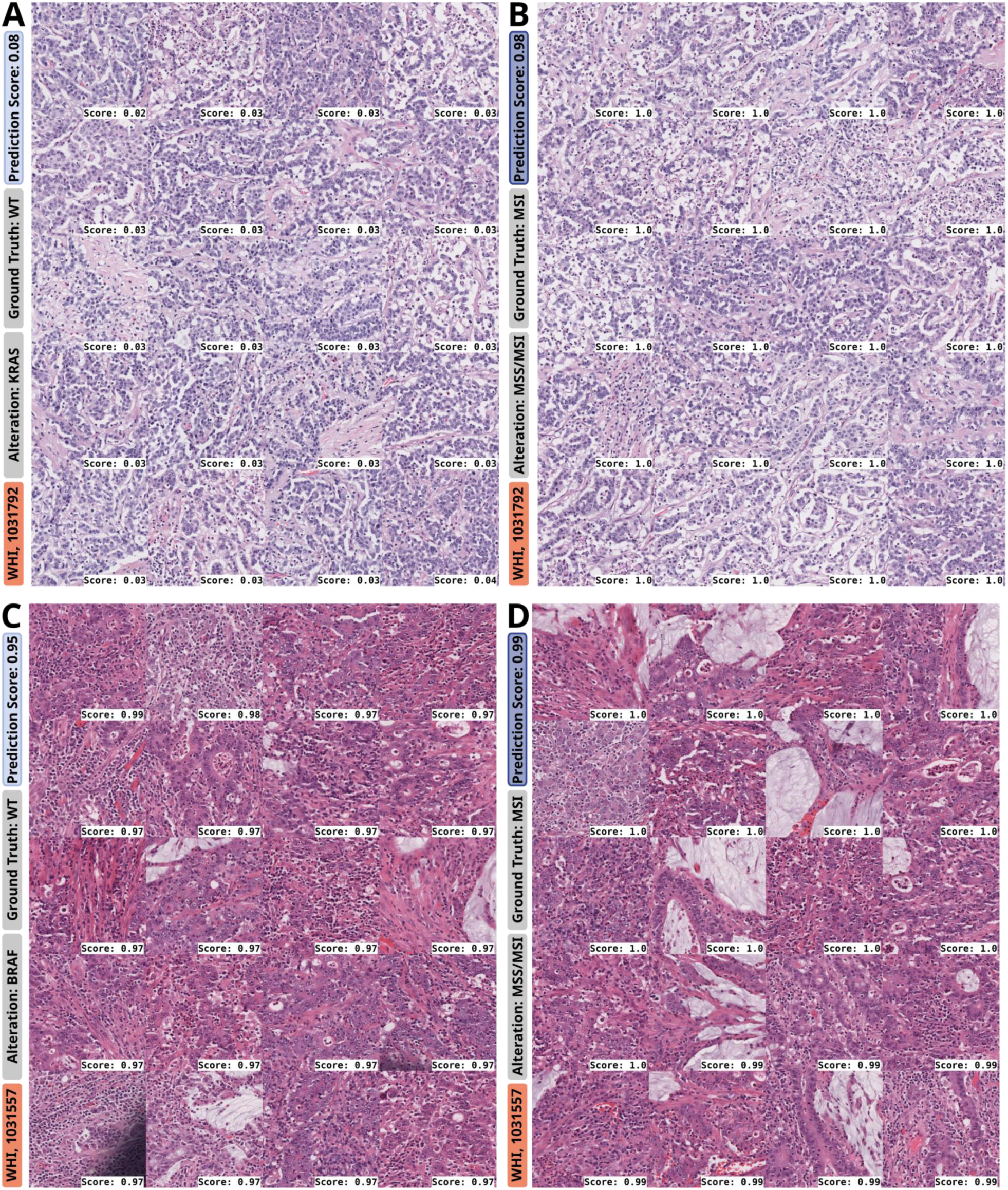
Top tiles for prediction of genetic alterations (left column) and MSI (right column) for two selected slides from the GECCO test set. **A.-B.** WHI, 1031792: Medullary carcinoma with sheets of tumor cells, low stroma content, high number of tumor-infiltrating lymphocytes in a *KRAS* WT and MSI case, leading to high MSI prediction scores (**B.**) and low *KRAS* MUT prediction scores (**A.**). *KRAS* mutations show a lower frequency in MSI CRCs. Medullary carcinoma is a key morphological feature of MSI CRCs. **C.-D.** WHI 1031557: Top tiles for *BRAF* MUT as well as MSI prediction, both predictions with high prediction scores, both displaying a mixed morphology with partly medullary, partly mucinous, partly gland-forming histology, and high number of tumorinfiltrating/associated lymphocytes. Medullary growth pattern with lymphocytic infiltration and mucinous differentiation are typical features of a MSI-like morphology. Accordingly, the case was correctly predicted as MSI with a really high prediction score (**D.**). As *BRAF* MUT and MSI often co-occur and share morphologic overlap, the case was misclassified with regards to *BRAF*-status, resulting in high prediction scores for *BRAF* MUT (**C.**), even though the ground truth was *BRAF* WT. Abbreviations: CRC: Colorectal cancer; MSI: Microsatellite instability; MSS: Microsatellite stability; MUT: Mutated; WT: Wild type.

Frequent MSI-associated patterns in most MSI cases and Cluster 2 gene mutations included medullary growth, high tumor-infiltrating lymphocytes (TILs), and mucinous differentiation, consistent with prior studies.^37–39^ However, some MSI cases in our cohort and the literature display atypical morphologies not commonly linked to MSI (e.g., Fig. S8D, Fig. S9D).^40^ In MSS-related Cluster 1, *KRAS* mutation predictions highlighted luminal tumor regions, especially villous adenomas with high-grade dysplasia, as equally important as invasive adenocarcinoma areas (Fig. S16A-B, Fig. S17A-B). This aligns with the known prevalence of *KRAS* mutations in villous adenomas^41^ and carcinomas with close proximity to polyps^42^, reflecting the algorithm’s ability to capture intratumoral heterogeneity. Tiles with high MSI relevance displayed medullary carcinoma patterns, including tumor cell sheets and high TILs, which corresponded to low *KRAS* MUT prediction scores (Fig. 6A-B). Alterations such as *TP53* and *APC*, less common in MSI-high cases, relied on MSS-like morphology (e.g., gland-forming conventional adenocarcinoma with dirty necrosis^40^; Fig. S18-S20), while MSI-associated alterations (e.g., *BRAF*, *RNF43*, hypermutation) depended on MSI-related features, including medullary patterns, mucinous differentiation (with signet-ring cells; Fig. S23B), and high TILs^40^ (Fig. 6C-D, Fig. S12-S15, Fig. S21, Tab. S19). To explore target-specific patterns beyond MSI, a pathological review of top tiles from selected slides (Fig. S22-S23) identified tumor budding as a morphological feature associated with *BRAF* mutations in MSS cases (Fig. S23A-C).^43^

## Discussion

Previous research in CRC has focused on high AUROCs for biomarker detection, showing MSI as the most detectable, while often neglecting co-occurrence with other mutations and lacking indepth analyses of multiple genetic alterations and their links to MSI-associated morphologies. To address these limitations, we developed a DL model to predict multiple genetic alterations in CRC from H&E slides, using five GECCO cohorts with unified sequencing data with additional validation on two public CRC-datasets, TCGA and CPTAC. The multi-target model achieved performance within the literature range, outperforming single-target models for some alterations while enabling the efficient and sustainable simultaneous prediction of numerous prediction targets. This in-depth analysis was made possible by the comprehensive dataset and novel model architecture, both of which are shared open-source with the scientific community.

Our cohort, comprising multiple sub-cohorts, is broadly representative of CRC patients^44^, particularly in Western populations, with data primarily from the US and Europe. It includes a higher proportion of female patients (IWHS, WHI) and disproportionately includes White individuals, while TCGA had the highest proportion of Black/African American patients, and CPTAC was limited to fresh-frozen colon samples with the smallest cohort. MSI frequency ranged from 7–35%, aligning with reported rates of 15–20%^45^ (Tab. S1). Variability in target mutation frequencies across cohorts, remained within plausible ranges, as did the experimental results. However, small sample sizes for some prediction targets likely contributed to higher variability (Fig. S2), underscoring the need for larger, more diverse datasets. The genetic clusters analyzed included key CRC biomarkers (e.g., *TP53*, *APC*, *KRAS*, *BRAF*)^7,8,11–13^, with mutational profiles and MSI co-occurrences aligning with prior studies^26,27,46^.

In line with the literature MSI was the most predictable alteration^7,8,12,13,25,36^, though some MSI cases scored low due to a lack of associated morphological features, which is plausible and pathological assessment has shown. For the detection of multiple genetic alterations apart from MSI, our multi-target model outperformed several of our own single-target models and models from the literature, such as for detection of *RNF43*, using the AUROC as a common performance measure.^4–9,35,36^ Notably, hypermutation prediction demonstrates a strong interdependence with 10 MSI due to their frequent co-occurrence and close biological linkage, with shared morphological features leading to lower scores for hypermutated MSS cases. *BRAF* mutations, critical in CRC for their prognostic and predictive significance^47^, showed detectable phenotypic changes, including mucinous differentiation and poorly differentiated clusters. The histopathological evaluation revealed features such as mucinous differentiation and stroma-rich patterns dominating *BRAF* MUT and *BRAF* WT tiles, respectively^12,47^ Interestingly, we could identify mucinous/signet-ring differentiation relevant for *BRAF* predictions.^47^ However, these distinct features overlapped with MSI-associated morphology, confounding specificity and occasionally leading to misclassification in MSS cases. In contrast, some top tiles for *BRAF* MUT prediction showed poorly differentiated clusters and tumor budding, which has already been linked to *BRAF* mutations^48^, and is associated with MSS.^43^ While mutations like *BRAF* induce distinct phenotypic changes detectable by DL, MSI morphology often overshadows fine-grained, target-specific patterns. Excluding MSI as a prediction target did not affect the predictability of other prediction targets, and independent class token behavior with no cross-token information flow in the decoder, a key feature of the model architecture, was demonstrated (Fig. S24). In summary, we have shown that CRC mutation predictability is primarily driven by MSI-associated morphology, the most discriminative feature for mutation detection, though subgroup analyses of MSI and MSS suggest subtle mutation-specific patterns warrant further study.

This study has several limitations. Despite its extent, the dataset was limiting the detection of rare alterations and subtle mutation-specific morphologies, highlighting the need for larger and more diverse datasets. Underrepresentation of Black/African-American individuals in the GECCO dataset and missing or incomplete annotations further constrain generalizability and accuracy. While monochrome tiles were excluded during preprocessing, some pen markings remained. These markings, though occasionally attracting slight model attention and potentially introducing bias, were largely transparent, limited to a few tiles, and may hold biologically relevant information, particularly near tumor borders, warranting further investigation. Additionally, our study emphasizes the need for advanced explainability methods to validate DL-detected pathological patterns and link them to specific prediction targets, fostering a bidirectional exchange between pathology and DL. This could clarify whether MSI-associated morphology dominates model predictions due to the absence of other target-specific features or its overwhelming influence. Approaches such as modified loss functions could help mitigate MSI dominance and enhance detection of subtle features^49^ and integrating multimodal data could improve generalizability, accuracy, and understanding of mutation-specific morphologies. Our model expands on single-target transformers and raises important questions regarding potential bias toward MSI-associated targets due to their larger subgroup size; however, as our model performs within the range reported in the literature, further investigation is required to elucidate the influence of MSI- and non-MSI-associated features, providing insights into the mechanisms of single- and multi-target transformers. Similar to prior research^34,50^, we show that AUROC can be a misleading metric for evaluating biomarker detection; thus, we report it for comparability but include additional metrics. For future studies, we recommend using extensive, diverse datasets, analyzing multiple prediction targets and their interactions with various metrics (e.g. Accuracy, Precision, Sensitivity, Specificity, F1 Score), and addressing potential confounding factors by leveraging approaches such as our proposed model architecture.

From a practical point of view, our multitarget DL-based biomarker prediction provides significant value for research by identifying which prediction targets can be reliably predicted and the morphological basis for these predictions, guiding future studies and highlighting prediction targets that merit further investigation. For some alterations, such as *BRAF*, existing surrogate immunohistochemical assays could be applied^51^ to predicted regions in follow-up studies to deepen understanding of the biological mechanisms behind predictions. Clinically, this approach streamlines colorectal cancer testing by eliminating the need for separate workflows and serving as a costeffective prescreening tool, particularly in resource-limited settings or earlier CRC stages. Biologically, it sheds light on how genetic alterations influence tumor morphology and underscores the dominant role of surrogate markers like MSI in CRC histology. As the comprehensive analysis of co-occurrences in CRC is scarcely explored in relation to DL and is rare for other cancer types^52^, this efficient, scalable method can be extended to any cancer type, enabling simultaneous analysis of multiple prediction targets and advancing both research and precision oncology.

In conclusion, our study highlights the utility of multi-target transformers for detecting biomarker-specific patterns in CRC H&E images, with MSI phenotype as the dominant factor influencing predictability. The model’s reliance on MSI- and MSS-associated morphology underscores the importance of morphology and co-occurrence patterns over AUROC metrics. By enabling the efficient investigation of numerous prediction targets simultaneously, this model facilitates a more comprehensive understanding of biomarker interactions. These findings emphasize the need to consider individual prediction scores and tumor-specific confounders in biomarker studies, laying the foundation for future research to address these effects across other cancer types.

## List of abbreviations

AUROC: Area under the Receiver Operating Characteristic Curve
AUPRC: Area Under the Precision-Recall Curve
CIN: Chromosomal instability
CPTAC: Clinical Proteomic Tumor Analysis Consortium
CRC: Colorectal cancer
DL: Deep Learning
H&E: Hematoxylin and eosin
Mb: Megabases
MSI: Microsatellite instability
MSS: Microsatellite stable
MUT: Mutated
NOS: Not otherwise specified
px: Pixel
ROC: Receiver Operating Characteristic Curve
TCGA: The Cancer Genome Atlas
TILs: Tumor infiltrating lymphocytes
ViT: Vision Transformer
WSI: Whole Slide Image
WT: Wild type

## Declarations

### Ethics approval and consent to participate

This study was performed in accordance with the Declaration of Helsinki. This study is a retrospective analysis of scanned images of anonymized tissue samples of various cohorts of cancer patients. Data were collected and anonymized and ethical approval was obtained. The overall analysis was approved by the Ethics board of the Medical Faculty of Technical University Dresden under the ID BO-EK-444102022·

### Contributions

MG and JNK conceptualized the study. CS, PH, LAB, AJF, ELG, AG, MJG, PL, NM, ST and UP provided clinical and digital histopathological data. MG curated the source data and conducted the literature research. MvT implemented the source code for the deep learning pipeline and the heatmaps. MG developed the code for data analysis and visualization. MG planned and conducted the experiments, visualizations and analysis of data and results. ARM assisted with the statistical evaluation. MG, CMLL, ZIC and NGR interpreted the data. NGR and BM did the pathological examination of the samples and analyzed the heatmaps and top/bottom tiles. MG wrote the first draft of the manuscript. All authors revised the manuscript draft, jointly interpreted the data and agreed to the submission of this article. All authors had access to all the data, and MG and JNK have verified the data.

### Data sharing

All source code we used to conduct this study is publicly available with publication. The code for image preprocessing is publicly available at https://github.com/KatherLab. The tesselation script is available at https://github.com/KatherLab/preprocessing-ng. Extraction of CTransPath features was conducted with scripts from https://github.com/KatherLab/marugoto. The scripts for the multi-target transformer and the heatmaps used for explainability can be accessed at https://github-com/LocalToasty/barspoon-transformer/. The saved checkpoints for the trained models with the lowest validation loss for all seven folds can be obtained from https://github.com/gustavmarco/barspoon-transformer/releases/tag/gustav2024<u>·</u> Links to the exact repository versions used can be found in Tab. S3. The digitized whole slide images (WSIs) for the TCGA ^22^ cohort are publicly accessible at https://portal.gdc.cancer.gov/ and https://www.cbioportal.org/. The digitized WSIs for the CPTAC cohort are publicly accessible at The Cancer Imaging Archive (TCIA) ^53^. The molecular and clinical data for TCGA and CPTAC is publicly accessible at https://portal.gdc.cancer.gov/ and https://www.cbioportal.org/. The datasets from the GECCO consortium are available from the corresponding author on reasonable request. All data generated or analyzed during this study are included in this published article and its supplementary information files.

## Acknowledgements

JNK is supported by the German Federal Ministry of Health (DEEP LIVER, ZMVI1-2520DAT111), the Max-Eder-Programme of German Cancer Aid (grant #70113864), the German Federal Ministry of Education and Research (PEARL, 01KD2104C; CAMINO, 01EO2101; SWAG, 01KD2215A; TRANSFORM LIVER, 031L0312A; TANGERINE, 01KT2302 through ERA-NET Transcan), the German Academic Exchange Service (SECAI, 57616814), the German Federal Joint Committee (Transplant.KI, 01VSF21048) the European Union’s Horizon Europe and innovation programme (ODELIA, 101057091; GENIAL, 101096312) and the National Institute for Health and Care Research (NIHR, NIHR213331) Leeds Biomedical Research Centre. The views expressed are those of the author(s) and not necessarily those of the NHS, the NIHR or the Department of Health and Social Care. SF is supported by the German Federal Ministry of Education and Research (SWAG, 01KD2215C), the German Cancer Aid (DECADE, 70115166 and TargHet, 70115995) and the German Research Foundation (504101714). The Genetics and Epidemiology of Colorectal Cancer Consortium (GECCO) is funded by: National Cancer Institute, National Institutes of Health, U.S. Department of Health and Human Services (U01 CA137088, R01 CA488857, P20 CA252733). Genotyping/Sequencing services were provided by the Center for Inherited Disease Research (CIDR) contract number HHSN268201700006I. This research was funded in part through the NIH/NCI Cancer Center Support Grant P30 CA015704· Scientific Computing Infrastructure at Fred Hutch funded by ORIP grant S10OD028685· The CORSA study was funded by Austrian Research Funding Agency (FFG) BRIDGE (grant 829675, to Andrea Gsur), the “Herzfelder’sche Familienstiftung” (grant to Andrea Gsur) and was supported by COST Action BM1206· CRA was supported by the National Institutes of Health grant R01 CA068535· The coordination of EPIC is financially supported by the International Agency for Research on Cancer (IARC) and also by the Department of Epidemiology and Biostatistics, School of Public Health, Imperial College London which has additional infrastructure support provided by the NIHR Imperial Biomedical Research Centre (BRC). The national cohorts are supported by: Danish Cancer Society (Denmark); Ligue Contre le Cancer, Institut Gustave Roussy, Mutuelle Générale de l’Education Nationale, Institut National de la Santé et de la Recherche Médicale (INSERM) (France); German Cancer Aid, German Cancer Research Center (DKFZ), German Institute of Human Nutrition Potsdam-Rehbruecke (DIfE), Federal Ministry of Education and Research (BMBF) (Germany); Associazione Italiana per la Ricerca sul Cancro-AIRC-Italy, Compagnia di SanPaolo and National Research Council (Italy); Dutch Ministry of Public Health, Welfare and Sports (VWS), Netherlands Cancer Registry (NKR), LK Research Funds, Dutch Prevention Funds, Dutch ZON (Zorg Onderzoek Nederland), World Cancer Research Fund (WCRF), Statistics Netherlands (The Netherlands); Health Research Fund (FIS) - Instituto de Salud Carlos III (ISCIII), Regional Governments of Andalucía, Asturias, Basque Country, Murcia and Navarra, and the Catalan Institute of Oncology - ICO (Spain); Swedish Cancer Society, Swedish Research Council and and Region Skåne and Region Västerbotten (Sweden); Cancer Research UK (14136 to EPIC-Norfolk; C8221/A29017 to EPIC-Oxford), Medical Research Council (1000143 to EPIC-Norfolk; MR/M012190/1 to EPIC-Oxford) (United Kingdom). The IWHS study was supported by NIH grants CA107333 (R01 grant awarded to P.J. Limburg) and HHSN261201000032C (N01 contract awarded to the University of Iowa). The WHI program is funded by the National Heart, Lung, and Blood Institute, National Institutes of Health, U.S. Department of Health and Human Services through contracts 75N92021D00001, 75N92021D00002, 75N92021D00003, 75N92021D00004, 75N92021D00005. We kindly thank all individuals who agreed to participate in the CORSA study. Furthermore, we thank all cooperating physicians and students and the Biobank Graz of the Medical University of Graz. We also acknowledge the TCGA Research Network and the Clinical Proteomic Tumor Analysis Consortium (CPTAC), which generated the data on which some of the results shown in this study are based. Where authors are identified as personnel of the International Agency for Research on Cancer/World Health Organization, the authors alone are responsible for the views expressed in this article and they do not necessarily represent the decisions, policy or views of the International Agency for Research on Cancer/World Health Organization. The authors thank the WHI investigators and staff for their dedication, and the study participants for making the program possible. A full listing of WHI investigators can be found at: http://www.whi.org/researchers/Documents%20%20Write%20a%20Paper/WHI%20Investigator%20Short%20List.pdf. The readability and language of the work were improved using ChatGPT-4o and DeepL.

