## Supplement for "Assessing Genotype-Phenotype Correlations with Deep Learning in Colorectal Cancer: A Multi-Centric Study"

### Supplementary Methods

#### Data Analysis of Genetic Alterations

We applied hierarchical clustering[1,2] to group genes and conditions, such as MSI and hypermutation, based on their mutational patterns. This method organizes data into a hierarchy of clusters, where similarities are quantified using the commonly applied 'Euclidean' metric, which measures distances in a multi-dimensional space. To optimize cluster formation, we used the 'Ward' method[3], which minimizes within-cluster variance, resulting in more balanced and interpretable clusters. To mitigate the influence of missing values, we restricted clustering to prediction targets with complete data (Table S1), ensuring robust identification of clusters. This approach revealed groups of related genetic alterations sharing similar mutational patterns among the prediction targets.

Subsequently, we employed association rule mining[4] to identify patterns of co-occurrence among gene mutations and conditions, such as MSI. This method evaluates the relationship between an initiating genetic alteration, referred to as the ‘antecedent’, and a potentially resulting alteration, termed the ‘consequent’, using several established metrics. Support[5] quantifies the frequency with which the antecedent and consequent co-occur within the dataset, while confidence[5] measures the conditional probability of observing the consequent given the presence of the antecedent, thereby reflecting the predictive strength of the association. Lift[5] assesses the degree of association by comparing the observed frequency of co-occurrence to what would be expected under statistical independence. Leverage[6] calculates the difference between the observed co-occurrence frequency and the expected frequency, highlighting the actual impact of the association. Conviction[7] gauges the reliability of the rule, with higher values indicating greater dependence of the consequent on the antecedent. Zhang’s Metric[8] further evaluates the relevance of the relationship by accounting for both the presence and absence of the antecedent and consequent, offering a more balanced assessment of the association's significance. These metrics must be interpreted cautiously and in conjunction to ensure meaningful conclusions, particularly given the inherent complexity of biological data and potential limitations arising from missing or sparse data.

### Supplementary Tables

**Tab. S1: Sociodemographic and clinicopathological patient characteristics of all cohorts.** We selected relevant and representative targets from all targets within the study as described in the manuscript. The use of the dataset as a train or test dataset is also indicated. 'N' denotes quantity, 'IQR' is interquartile range, 'WT' is wild type, 'MUT' is mutated, and 'NaN' indicates missing information.

| | | **GECCO (primary dataset)** | | | | | **Public Cohorts** | |
|---|---|---|---|---|---|---|---|---|
| | | **EPIC** | **CORSA** | **IWHS** | **CRA** | **WHI** | **TCGA** | **CPTAC** |
| Dataset | | train | train | train | test | test | test | test |
| N Patients | | 183 | 158 | 390 | 321 | 324 | 426 | 110 |
| Age | Median | 62 | 69 | 63 | 67 | 65 | 67 | 66 |
| | IQR | 13 | 16 | 6 | 15 | 10 | 18 | 18 |
| | <50 | 17 (9%) | 10 (6%) | 0 (0%) | 29 (9%) | 0 (0%) | 59 (14%) | 5 (5%) |
| | NaN | 0 (0%) | 1 (1%) | 0 (0%) | 0 (0%) | 0 (0%) | 1 (0%) | 2 (2%) |
| Sex | Male | 83 (45%) | 99 (63%) | 0 (0%) | 183 (57%) | 0 (0%) | 212 (50%) | 44 (40%) |
| | Female | 100 (55%) | 59 (37%) | **390 (100%)** | 138 (43%) | **324 (100%)** | 213 (50%) | 66 (60%) |
| | NaN | 0 (0%) | 0 (0%) | 0 (0%) | 0 (0%) | 0 (0%) | 1 (0%) | 0 (0%) |
| Race | White | 183 (100%) | 158 (100%) | 385 (99%) | 106 (33%) | 292 (90%) | 244 (57%) | 81 (74%) |
| | Black/African-American | 0 (0%) | 0 (0%) | 0 (0%) | 4 (1%) | 14 (4%) | 57 (13%) | 7 (6%) |
| | American Indian/Alaska Native | 0 (0%) | 0 (0%) | 0 (0%) | 0 (0%) | 1 (0%) | 1 (0%) | 1 (1%) |
| | Asian | 0 (0%) | 0 (0%) | 0 (0%) | 0 (0%) | 3 (1%) | 12 (3%) | 18 (16%) |
| | NaN | 0 (0%) | 0 (0%) | 5 (1%) | 211 (66%) | 14 (4%) | 112 (26%) | 3 (3%) |
| Cancer Site | Colon | 96 (52%) | 99 (63%) | 323 (83%) | 223 (69%) | 287 (89%) | 317 (74%) | 110 (100%) |
| | Rectum | 47 (26%) | 50 (32%) | 63 (16%) | 98 (31%) | 32 (10%) | 105 (25%) | 0 (0%) |
| | NaN | 40 (22%) | 9 (6%) | 4 (1%) | 0 (0%) | 5 (2%) | 4 (1%) | 0 (0%) |
| T-Stage | T1 | 1 (1%) | 11 (7%) | 0 (0%) | 23 (7%) | 24 (7%) | 14 (3%) | 0 (0%) |
| | T2 | 1 (1%) | 29 (18%) | 0 (0%) | 43 (13%) | 56 (17%) | 72 (17%) | 17 (15%) |
| | T3 | 4 (2%) | 95 (60%) | 0 (0%) | 172 (54%) | 170 (52%) | 292 (69%) | 79 (72%) |
| | T4 | 3 (2%) | 18 (11%) | 0 (0%) | 20 (6%) | 68 (21%) | 47 (11%) | 14 (13%) |
| | TX | 0 (0%) | 1 (1%) | 0 (0%) | 2 (1%) | 0 (0%) | 0 (0%) | 0 (0%) |
| | NaN | 174 (95%) | 4 (3%) | 390 (100%) | 61 (19%) | 6 (2%) | 1 (0%) | 0 (0%) |
| M-Stage | M0 | 8 (4%) | 72 (46%) | 0 (0%) | 214 (67%) | 292 (90%) | 311 (73%) | 102 (93%) |
| | M1 | 1 (1%) | 20 (13%) | 0 (0%) | 34 (11%) | 26 (8%) | 55 (13%) | 7 (6%) |
| | MX | 0 (0%) | 0 (0%) | 0 (0%) | 12 (4%) | 0 (0%) | 54 (13%) | 0 (0%) |
| | NaN | 174 (95%) | 66 (42%) | 390 (100%) | 61 (19%) | 6 (2%) | 6 (1%) | 1 (1%) |
| N-Stage | N0 | 8 (4%) | 74 (47%) | 0 (0%) | 157 (49%) | 191 (59%) | 235 (55%) | 59 (54%) |
| | N1 | 1 (1%) | 48 (30%) | 0 (0%) | 59 (18%) | 68 (21%) | 115 (27%) | 35 (32%) |
| | N2 | 0 (0%) | 27 (17%) | 0 (0%) | 43 (13%) | 51 (16%) | 74 (17%) | 16 (15%) |
| | NX | 0 (0%) | 3 (2%) | 0 (0%) | 1 (0%) | 0 (0%) | 1 (0%) | 0 (0%) |
| | NaN | 174 (95%) | 6 (4%) | 390 (100%) | 61 (19%) | 14 (4%) | 1 (0%) | 0 (0%) |
| ACVR1B | WT | 177 (97%) | 150 (95%) | 369 (95%) | 306 (95%) | 300 (93%) | 405 (95%) | 107 (97%) |

| | | GECCO (primary dataset) | | | | | Public Cohorts | |
|---|---|---|---|---|---|---|---|---|
| | | **EPIC** | **CORSA** | **IWHS** | **CRA** | **WHI** | **TCGA** | **CPTAC** |
| | MUT | 6 (3%) | 8 (5%) | 21 (5%) | 15 (5%) | 24 (7%) | 21 (5%) | 3 (3%) |
| | NaN | 0 (0%) | 0 (0%) | 0 (0%) | 0 (0%) | 0 (0%) | 0 (0%) | 0 (0%) |
| AKT1 | WT | 176 (96%) | 150 (95%) | 383 (98%) | 310 (97%) | 305 (94%) | 417 (98%) | 109 (99%) |
| | MUT | 7 (4%) | 8 (5%) | 7 (2%) | 11 (3%) | 19 (6%) | 9 (2%) | 1 (1%) |
| | NaN | 0 (0%) | 0 (0%) | 0 (0%) | 0 (0%) | 0 (0%) | 0 (0%) | 0 (0%) |
| ALK | WT | 169 (92%) | 148 (94%) | 362 (93%) | 295 (92%) | 297 (92%) | 398 (93%) | 102 (93%) |
| | MUT | 14 (8%) | 10 (6%) | 28 (7%) | 26 (8%) | 27 (8%) | 28 (7%) | 8 (7%) |
| | NaN | 0 (0%) | 0 (0%) | 0 (0%) | 0 (0%) | 0 (0%) | 0 (0%) | 0 (0%) |
| APC | WT | 64 (35%) | 72 (46%) | 163 (42%) | 90 (28%) | 174 (54%) | 91 (21%) | 27 (25%) |
| | MUT | 119 (65%) | 86 (54%) | 227 (58%) | 231 (72%) | 150 (46%) | 335 (79%) | 83 (7y5%) |
| | NaN | 0 (0%) | 0 (0%) | 0 (0%) | 0 (0%) | 0 (0%) | 0 (0%) | 0 (0%) |
| ATM | WT | 162 (89%) | 148 (94%) | 331 (85%) | 289 (90%) | 297 (92%) | 369 (87%) | 101 (92%) |
| | MUT | 21 (11%) | 10 (6%) | 59 (15%) | 32 (10%) | 27 (8%) | 57 (13%) | 9 (8%) |
| | NaN | 0 (0%) | 0 (0%) | 0 (0%) | 0 (0%) | 0 (0%) | 0 (0%) | 0 (0%) |
| BMPR2 | WT | 157 (86%) | 145 (92%) | 324 (83%) | 276 (86%) | 263 (81%) | 395 (93%) | 96 (87%) |
| | MUT | 26 (14%) | 13 (8%) | 66 (17%) | 45 (14%) | 61 (19%) | 31 (7%) | 14 (13%) |
| | NaN | 0 (0%) | 0 (0%) | 0 (0%) | 0 (0%) | 0 (0%) | 0 (0%) | 0 (0%) |
| BRAF | WT | 132 (72%) | 148 (94%) | 271 (69%) | 242 (75%) | 268 (83%) | 370 (87%) | 92 (84%) |
| | MUT | 51 (28%) | 10 (6%) | 119 (31%) | 79 (25%) | 56 (17%) | 56 (13%) | 18 (16%) |
| | NaN | 0 (0%) | 0 (0%) | 0 (0%) | 0 (0%) | 0 (0%) | 0 (0%) | 0 (0%) |
| CCDC40 | WT | 176 (96%) | 152 (96%) | 373 (96%) | 312 (97%) | 310 (96%) | 411 (96%) | 103 (94%) |
| | MUT | 7 (4%) | 6 (4%) | 17 (4%) | 9 (3%) | 14 (4%) | 15 (4%) | 7 (6%) |
| | NaN | 0 (0%) | 0 (0%) | 0 (0%) | 0 (0%) | 0 (0%) | 0 (0%) | 0 (0%) |
| CDK12 | WT | 178 (97%) | 152 (96%) | 378 (97%) | 307 (96%) | 309 (95%) | 399 (94%) | 107 (97%) |
| | MUT | 5 (3%) | 6 (4%) | 12 (3%) | 14 (4%) | 15 (5%) | 27 (6%) | 3 (3%) |
| | NaN | 0 (0%) | 0 (0%) | 0 (0%) | 0 (0%) | 0 (0%) | 0 (0%) | 0 (0%) |
| CHD1 | WT | 169 (92%) | 0 (0%) | 356 (91%) | 310 (97%) | 0 (0%) | 404 (95%) | 103 (94%) |
| | MUT | 14 (8%) | 0 (0%) | 34 (9%) | 11 (3%) | 0 (0%) | 22 (5%) | 7 (6%) |
| | NaN | 0 (0%) | 158 (100%) | 0 (0%) | 0 (0%) | 324 (100%) | 0 (0%) | 0 (0%) |
| CTNND1 | WT | 177 (97%) | 156 (99%) | 369 (95%) | 310 (97%) | 307 (95%) | 406 (95%) | 108 (98%) |
| | MUT | 6 (3%) | 2 (1%) | 21 (5%) | 11 (3%) | 17 (5%) | 20 (5%) | 2 (2%) |
| | NaN | 0 (0%) | 0 (0%) | 0 (0%) | 0 (0%) | 0 (0%) | 0 (0%) | 0 (0%) |
| DUSP16 | WT | 179 (98%) | 155 (98%) | 377 (97%) | 312 (97%) | 317 (98%) | 425 (100%) | 104 (95%) |
| | MUT | 4 (2%) | 3 (2%) | 13 (3%) | 9 (3%) | 7 (2%) | 1 (0%) | 6 (5%) |
| | NaN | 0 (0%) | 0 (0%) | 0 (0%) | 0 (0%) | 0 (0%) | 0 (0%) | 0 (0%) |
| ELL2 | WT | 176 (96%) | 0 (0%) | 378 (97%) | 317 (99%) | 0 (0%) | 416 (98%) | 105 (95%) |
| | MUT | 7 (4%) | 0 (0%) | 12 (3%) | 4 (1%) | 0 (0%) | 10 (2%) | 5 (5%) |
| | NaN | 0 (0%) | 158 (100%) | 0 (0%) | 0 (0%) | 324 (100%) | 0 (0%) | 0 (0%) |
| FHOD3 | WT | 164 (90%) | 0 (0%) | 335 (86%) | 285 (89%) | 0 (0%) | 395 (93%) | 96 (87%) |
| | MUT | 19 (10%) | 0 (0%) | 55 (14%) | 36 (11%) | 0 (0%) | 31 (7%) | 14 (13%) |
| | NaN | 0 (0%) | 158 (100%) | 0 (0%) | 0 (0%) | 324 (100%) | 0 (0%) | 0 (0%) |
| Hypermutated (HM) | not HM | 143 (78%) | 127 (80%) | 286 (73%) | 254 (79%) | 213 (66%) | 358 (84%) | 0 (0%) |
| | HM | 40 (22%) | 31 (20%) | 104 (27%) | 67 (21%) | 111 (34%) | 68 (16%) | 0 (0%) |
| | NaN | 0 (0%) | 0 (0%) | 0 (0%) | 0 (0%) | 0 (0%) | 0 (0%) | 110 (100%) |
| KIF1A | WT | 163 (89%) | 135 (85%) | 353 (91%) | 286 (89%) | 296 (91%) | 399 (94%) | 101 (92%) |
| | MUT | 20 (11%) | 23 (15%) | 37 (9%) | 35 (11%) | 28 (9%) | 27 (6%) | 9 (8%) |
| | NaN | 0 (0%) | 0 (0%) | 0 (0%) | 0 (0%) | 0 (0%) | 0 (0%) | 0 (0%) |
| KRAS | WT | 124 (68%) | 101 (64%) | 266 (68%) | 205 (64%) | 226 (70%) | 238 (56%) | 74 (67%) |

| | | GECCO (primary dataset) | | | | | Public Cohorts | |
|---|---|---|---|---|---|---|---|---|
| | | **EPIC** | **CORSA** | **IWHS** | **CRA** | **WHI** | **TCGA** | **CPTAC** |
| | MUT | 59 (32%) | 57 (36%) | 124 (32%) | 116 (36%) | 98 (30%) | 188 (44%) | 36 (33%) |
| | NaN | 0 (0%) | 0 (0%) | 0 (0%) | 0 (0%) | 0 (0%) | 0 (0%) | 0 (0%) |
| MECOM | WT | 173 (95%) | 0 (0%) | 350 (90%) | 295 (92%) | 0 (0%) | 403 (95%) | 102 (93%) |
| | MUT | 10 (5%) | 0 (0%) | 40 (10%) | 26 (8%) | 0 (0%) | 23 (5%) | 8 (7%) |
| | NaN | 0 (0%) | 158 (100%) | 0 (0%) | 0 (0%) | 324 (100%) | 0 (0%) | 0 (0%) |
| MSS/ MSI | MSS | 143 (78%) | 147 (93%) | 283 (73%) | 256 (80%) | 212 (65%) | 365 (86%) | 84 (76%) |
| | MSI | 40 (22%) | 11 (7%) | 107 (27%) | 65 (20%) | 112 (35%) | 61 (14%) | 25 (23%) |
| | NaN | 0 (0%) | 0 (0%) | 0 (0%) | 0 (0%) | 0 (0%) | 0 (0%) | 1 (1%) |
| NRAS | WT | 174 (95%) | 148 (94%) | 376 (96%) | 305 (95%) | 312 (96%) | 395 (93%) | 103 (94%) |
| | MUT | 9 (5%) | 10 (6%) | 14 (4%) | 16 (5%) | 12 (4%) | 31 (7%) | 7 (6%) |
| | NaN | 0 (0%) | 0 (0%) | 0 (0%) | 0 (0%) | 0 (0%) | 0 (0%) | 0 (0%) |
| PIK3CA | WT | 148 (81%) | 143 (91%) | 291 (75%) | 262 (82%) | 278 (86%) | 307 (72%) | 87 (79%) |
| | MUT | 35 (19%) | 15 (9%) | 99 (25%) | 59 (18%) | 46 (14%) | 119 (28%) | 23 (21%) |
| | NaN | 0 (0%) | 0 (0%) | 0 (0%) | 0 (0%) | 0 (0%) | 0 (0%) | 0 (0%) |
| PLEKHA6 | WT | 169 (92%) | 0 (0%) | 346 (89%) | 292 (91%) | 0 (0%) | 402 (94%) | 102 (93%) |
| | MUT | 14 (8%) | 0 (0%) | 44 (11%) | 29 (9%) | 0 (0%) | 24 (6%) | 8 (7%) |
| | NaN | 0 (0%) | 158 (100%) | 0 (0%) | 0 (0%) | 324 (100%) | 0 (0%) | 0 (0%) |
| RFX5 | WT | 176 (96%) | 0 (0%) | 369 (95%) | 308 (96%) | 0 (0%) | 411 (96%) | 105 (95%) |
| | MUT | 7 (4%) | 0 (0%) | 21 (5%) | 13 (4%) | 0 (0%) | 15 (4%) | 5 (5%) |
| | NaN | 0 (0%) | 158 (100%) | 0 (0%) | 0 (0%) | 324 (100%) | 0 (0%) | 0 (0%) |
| RNF43 | WT | 149 (81%) | 148 (94%) | 294 (75%) | 268 (83%) | 234 (72%) | 386 (91%) | 93 (85%) |
| | MUT | 34 (19%) | 10 (6%) | 96 (25%) | 53 (17%) | 90 (28%) | 40 (9%) | 17 (15%) |
| | NaN | 0 (0%) | 0 (0%) | 0 (0%) | 0 (0%) | 0 (0%) | 0 (0%) | 0 (0%) |
| SMAD2 | WT | 174 (95%) | 153 (97%) | 376 (96%) | 306 (95%) | 306 (94%) | 404 (95%) | 100 (91%) |
| | MUT | 9 (5%) | 5 (3%) | 14 (4%) | 15 (5%) | 18 (6%) | 22 (5%) | 10 (9%) |
| | NaN | 0 (0%) | 0 (0%) | 0 (0%) | 0 (0%) | 0 (0%) | 0 (0%) | 0 (0%) |
| SMG1 | WT | 172 (94%) | 150 (95%) | 377 (97%) | 308 (96%) | 311 (96%) | 408 (96%) | 102 (93%) |
| | MUT | 11 (6%) | 8 (5%) | 13 (3%) | 13 (4%) | 13 (4%) | 18 (4%) | 8 (7%) |
| | NaN | 0 (0%) | 0 (0%) | 0 (0%) | 0 (0%) | 0 (0%) | 0 (0%) | 0 (0%) |
| TBX3 | WT | 174 (95%) | 150 (95%) | 376 (96%) | 302 (94%) | 306 (94%) | 407 (96%) | 102 (93%) |
| | MUT | 9 (5%) | 8 (5%) | 14 (4%) | 19 (6%) | 18 (6%) | 19 (4%) | 8 (7%) |
| | NaN | 0 (0%) | 0 (0%) | 0 (0%) | 0 (0%) | 0 (0%) | 0 (0%) | 0 (0%) |
| TGFBR2 | WT | 176 (96%) | 152 (96%) | 364 (93%) | 304 (95%) | 297 (92%) | 411 (96%) | 104 (95%) |
| | MUT | 7 (4%) | 6 (4%) | 26 (7%) | 17 (5%) | 27 (8%) | 15 (4%) | 6 (5%) |
| | NaN | 0 (0%) | 0 (0%) | 0 (0%) | 0 (0%) | 0 (0%) | 0 (0%) | 0 (0%) |
| TP53 | WT | 64 (35%) | 57 (36%) | 158 (41%) | 109 (34%) | 194 (60%) | 150 (35%) | 51 (46%) |
| | MUT | 119 (65%) | 101 (64%) | 232 (59%) | 212 (66%) | 130 (40%) | 276 (65%) | 59 (54%) |
| | NaN | 0 (0%) | 0 (0%) | 0 (0%) | 0 (0%) | 0 (0%) | 0 (0%) | 0 (0%) |
| TRPS1 | WT | 171 (93%) | 0 (0%) | 353 (91%) | 293 (91%) | 0 (0%) | 381 (89%) | 92 (84%) |
| | MUT | 12 (7%) | 0 (0%) | 37 (9%) | 28 (9%) | 0 (0%) | 45 (11%) | 18 (16%) |
| | NaN | 0 (0%) | 158 (100%) | 0 (0%) | 0 (0%) | 324 (100%) | 0 (0%) | 0 (0%) |
| WNT16 | WT | 175 (96%) | 0 (0%) | 371 (95%) | 307 (96%) | 0 (0%) | 420 (99%) | 99 (90%) |
| | MUT | 8 (4%) | 0 (0%) | 19 (5%) | 14 (4%) | 0 (0%) | 6 (1%) | 11 (10%) |
| | NaN | 0 (0%) | 158 (100%) | 0 (0%) | 0 (0%) | 324 (100%) | 0 (0%) | 0 (0%) |
| ZHX2 | WT | 174 (95%) | 152 (96%) | 381 (98%) | 313 (98%) | 310 (96%) | 413 (97%) | 106 (96%) |
| | MUT | 9 (5%) | 6 (4%) | 9 (2%) | 8 (2%) | 14 (4%) | 13 (3%) | 4 (4%) |

| | | GECCO (primary dataset) | | | | | Public Cohorts | |
|---|---|---|---|---|---|---|---|---|
| | | **EPIC** | **CORSA** | **IWHS** | **CRA** | **WHI** | **TCGA** | **CPTAC** |
| | NaN | 0 (0%) | 0 (0%) | 0 (0%) | 0 (0%) | 0 (0%) | 0 (0%) | 0 (0%) |
| ZNRF3 | WT | 162 (89%) | 150 (95%) | 340 (87%) | 288 (90%) | 287 (89%) | 413 (97%) | 101 (92%) |
| | MUT | 21 (11%) | 8 (5%) | 50 (13%) | 33 (10%) | 37 (11%) | 13 (3%) | 9 (8%) |
| | NaN | 0 (0%) | 0 (0%) | 0 (0%) | 0 (0%) | 0 (0%) | 0 (0%) | 0 (0%) |

**Tab. S2: List of included targets from data provided by GECCO.** The table includes target names and explanations where clarification is required. These targets were used in both the primary and secondary models.

| Target 1 | Explanation 1 | Target 2 | Explanation 2 | Target 3 | Explanation 3 |
|---|---|---|---|---|---|
| ABCA8 | | LIMCH1 | | SETD2 | |
| ACVR1B | | LMO7 | | SIN3A | |
| ACVR2A | | LRRN3 | | SLC12A5 | |
| AKAP7 | | MAMDC4 | | SLC1A3 | |
| AKT1 | | MAML2 | | SMAD2 | |
| ALK | | MAP2K4 | | SMAD3 | |
| AMER1 | | MAP2K7 | | SMAD4 | |
| APC | | MAST2 | | SMARCA4 | |
| APC_NM_000038_truncated_first1600AA | Mutated yes/no. Truncating mutations within the first 1,600 amino acids for transcript NM000038 exclusively. | MBD6 | | SMG1 | |
| ARID1A | | MECOM | | SOS1 | |
| ARID1B | | MLH1 | | SOX9 | |
| ARID2 | | MLH3 | | SYNE1 | |
| ARID3A | | MMR | Pathway mutated yes/no in any genes:MLH1, MLH3, MSH2, MSH6, PMS2 | SYT3 | |
| ASXL1 | | MSH6 | | T2A | Number of T -> A and A -> T transversions |
| ATG2A | | MSI | | T2C | Number of T -> C and A -> G transitions/transversions |
| ATM | | MTOR | | T2G | Number of T -> G and A -> C transversions |
| ATXN1 | | MTUS2 | | TAF1L | |
| AXIN1 | | MUC4 | | TAF3 | |
| AXIN2 | | MUTYH | | TBX3 | |
| B2M | | MXRA5 | | TCERG1 | |
| BCL9 | | MYH9 | | TCF7L2 | |
| BCL9L | | NCAPD3 | | TCHH | |
| BCOR | | NFE2L3 | | TET1 | |
| BIRC6 | | NLGN4X | | TET2 | |
| BMPR2 | | NOD2 | | TET3 | |
| BRAF | | NRAS | | TEX14 | |
| BRAF_NM_004333_V600 | Mutated yes/no of all nonsynonymous SNVs in BRAF transcript NM_004333 at codon 600. | NRAS_NM_002524_Oncogenic_known | Mutated yes/no of all nonsynonymous SNVs in NRAS transcript NM_002524 codons 12,13 and 61. | TGFBR1 | |
| C2A | Number of C -> A and G -> T transversions | NRG1 | | TGFBR2 | |
| C2G | Number of C -> G and G -> C transversions | OSBPL6 | | TGF_beta | Pathway mutated yes/no in any genes: ACVR1B, ACVR2A, BMPR1A, BMPR2, GDF5, SMAD2, SMAD3, SMAD4, TGFBR1, TGFBR2 |
| C2T | Number of C -> T and G -> A transitions | PATIENT | | TGIF1 | |
| CACNG3 | | PAX5 | | TLR9 | |
| CALD1 | | PBRM1 | | TNRC6B | |
| CASP8 | | PCBP1 | | TP53 | |
| CCDC13 | | PCDH10 | | TP53BP1 | |
| CCDC40 | | PCDHA3 | | TP53_NM_000546_non_silent | TP53_NM_000546_Oncogenic_known or TP53_NM_000546_other_non_silent mutated yes/no. |
| CDH1 | | PCDHGA7 | | TPR | |
| CDK12 | | PCDHGA9 | | TRPS1 | |
| CDKN2A | | PCDHGB1 | | TSHZ2 | |
| CHD1 | | PIK3CA | | TYRO3 | |
| CHD4 | | PIK3CA_NM_006218_nonsynonymous_SNV | Mutated yes/no of all nonsynonymous SNVs in transcript NM_006218 (PIK3CA). | USP9X | |
| CPEB2 | | PIK3R1 | | UTP20 | |
| CRTC1 | | PIK3R1_NM_181523_non_silent | Mutated yes/no of all non-silent mutations called for PIK3R1 transcript NM_181523. | WNT | Pathway mutated yes/no in any genes: AMER1, APC, ARID1A, AXIN1, AXIN2, CTNNB1, FBXW7, RNF43, SOX9, TCF7, TCF7L2, ZNRF3 |
| CSMD1 | | PLEKHA6 | | WNT16 | |
| CTCF | | PLK1 | | XPO6 | |
| CTNNB1 | | POLD1 | | XYLT2 | |
| CTNND1 | | POLE | | ZBTB20 | |
| CUX1 | | POLQ | | ZBTB7A | |
| DAB2 | | PTEN | | ZDHHC8 | |

| | | | | | |
|---|---|---|---|---|---|
| DAPK1 | | PTEN_NM_000314_non_silent | Mutated yes/no of all non-silent mutations called for PTEN transcript NM_000314. | ZFHX3 | |
| DCAF4L1 | | RB1 | | ZFP36L2 | |
| DCC | | RBM10 | | ZHX2 | |
| DCHS1 | | RECQL5 | | ZNF512B | |
| DNMT1 | | RFX5 | | ZNF521 | |
| DOCK3 | | RGMB | | ZNRF3 | |
| DPYD | | RGS12 | | alcohol_ref | Alcohol use, at the reference time |
| DUSP16 | | RNF43 | | alcoholc2 | Alcohol use (nondrinker vs 1-28g/day) |
| DYNC1H1 | | RNF43_NM_001305544_truncating | Mutated yes/no of all truncating mutations called for transcript NM_001305544. | alcoholc2_1-28g/d | |
| ELF3 | | RTK_RAS | Pathway mutated yes/no in any genes:BRAF, ERBB2, ERBB3, KRAS, NRAS | alcoholc2_nondrinker | |
| ELL2 | | RTK_RAS_EM_TS_combined | | asp_ref(2) | Regular aspirin/NSAID use at referent time, definition1 |
| ELMO1 | | RYR1 | | asp_ref_missing | |
| ENAM | | S1 | | aspirin | Aspirin use? |
| EP300 | | S10 | | aspirin_ever | Aspirin, ever used regularly? |
| EP400 | | S11 | | diab | Ever diagnosed with diabetes by a doctor? |
| ERBB2 | | S12 | | horm_ref | Any postmenopausal hormone use? |
| ERBB3 | | S14 | | hrt_ref_pm(x) | Any Post-menopausal HRT use at reference time, definition1 |
| ERCC5 | | S15 | | hypermutated | |
| ESR1 | | S16 | | log_n_snv | Log transformed number of SNVs called from tumor-normal bam pair |
| FAN1 | | S17 | | log_total_mutations | Log transformed total number of SNVs and InDels mutations |
| FAT1 | | S18 | | n_indel | Total number of InDels mutations |
| FBLN2 | | S19 | | n_snv | Total number of SNVs called from tumor-normal bam pair (only pass-filter SNVs) |
| FBXW7 | | S1PR4 | | non_silent_2_silent_ratio | Non-silent to silent ratio. |
| FGFR1 | | S20 | | nsaids | Non-aspirin NSAIDS use? |
| FHOD3 | | S21 | | nsaids_ever | Non-aspirin NSAIDS, ever used regularly? |
| GDF5 | | S22 | | p53 | Pathway mutated yes/no in any genes: ATM, TP53 |
| GNAS | | S23 | | smk_ever | Ever smoked cigarettes? |
| GPATCH8 | | S24 | | smoke | Smoking status |
| GPC5 | | S25 | | smoke_Former smoker | |
| HCN1 | | S26 | | smoke_Never smoker | |
| HGF | | S28 | | smoke_Smoker | |
| IGF2_PI3K | Pathway mutated yes/no in any genes: IGF2, PIK3CA, PIK3R1, PTEN | S29 | | white_nonhisp | Non-Hispanic white |
| ING1 | | S3 | | | |
| KDM6A | | S30 | | | |
| KIF1A | | S4 | | | |
| KLF3 | | S5 | | | |
| KMT2B | | S6 | | | |
| KMT2C | | S7 | | | |
| KMT2D | | S8 | | | |
| KRAS | | S9 | | | |
| KRAS_NM_033360_Oncogenic_known | Mutated yes/no of all nonsynonymous SNVs in KRAS transcript NM_033360 codons 12, 13, 61, 117, 146. | SALL4 | | | |
| | | SCN5A | | | |

**Tab. S3: Links to directories containing the code used for the study and trained models.**

| Process | Link |
|---|---|
| Tessellation | https://github.com/KatherLab/preprocessing-ng/tree/1f5fdebf669363cf67bb422bb4cb0f91218d9c29 |
| Feature extraction with CTransPath | https://github.com/KatherLab/marugoto/tree/d401e1157635273cd4a99ca6e60c83db7ea09a22 |
| Multi-Target Transformer model, Heatmaps | https://github.com/gustavmarco/barspoon-transformer/tree/08b30a6cf80c8ed44c59fecb5a367b307da3284a |
| Top-tiles | https://github.com/LocalToasty/barspoon-transformer/tree/276d288cfeaaebf44c4d7f4e03dfe003c5bc7bc3 |
| Trained models from 7 folds | https://github.com/gustavmarco/barspoon-transformer/releases/tag/gustav2024 |

**Tab. S4: Comparative analysis of the internal and external performance of Multi-Target and Single-Target Transformers.** The performance is evaluated using the mean Area Under Precision Recall Curve (AUROC) from the 7 folds of the cross-validation for relevant selected prediction targets. AUROCs greater than 0·75 are highlighted in bold. The table includes results for the Single-Target Transformer for high relevance prediction targets from genetic Clusters 1-2 (Fig. 2). A two-sided DeLong test was conducted, indicating the fold results of Multi-Target versus Single-Target Transformers.

| | **Multi-Target Transformer** | | | **Single-Target Transformer** | | | **DeLong test p-value** |
|---|---|---|---|---|---|---|---|
| **Target** | **AUROC** internal | **Mean (std) AUROC**external | **Median AUROC**external | **AUROC**internal | **Mean (std) AUROC**external | **Median AUROC**external | |
| ACVR1B | 0·68 | **0·75 (± 0·03)** | **0·77** | | | | |
| AKT1 | **0·75** | 0·70 (± 0·03) | 0·70 | | | | |
| ALK | 0·63 | 0·70 (± 0·02) | 0·71 | | | | |
| APC | 0·6 | 0·66 (± 0·02) | 0·67 | 0·58 | 0·65 (± 0·03) | 0·66 | 0·7142 |
| ATM | 0·59 | 0·63 (± 0·01) | 0·63 | | | | |
| BMPR2 | **0·82** | **0·87 (± 0·01)** | **0·87** | **0·80** | **0·81 (± 0·03)** | **0·82** | 0·0001 |
| BRAF | **0·79** | **0·78 (± 0·01)** | **0·78** | **0·75** | **0·72 (± 0·06)** | **0·75** | <0·0001 |
| CCDC40 | 0·68 | 0·76 (± 0·04) | **0·78** | | | | |
| CDK12 | 0·69 | 0·78 (± 0·02) | **0·78** | | | | |
| CHD1 | **0·78** | **0·87 (± 0·02)** | **0·88** | | | | |
| CTNND1 | **0·75** | 0·75 (± 0·03) | 0·73 | | | | |
| DUSP16 | 0·61 | 0·74 (± 0·04) | 0·74 | | | | |
| ELL2 | **0·76** | **0·79 (± 0·05)** | **0·81** | | | | |
| FHOD3 | **0·77** | **0·84 (± 0·02)** | **0·84** | | | | |
| hypermu-tated | **0·80** | **0·88 (± 0·01)** | **0·88** | **0·82** | **0·86 (± 0·03)** | **0·87** | 0·2383 |
| KIF1A | 0·67 | 0·67 (± 0·03) | 0·67 | | | | |
| KRAS | 0·63 | 0·65 (± 0·03) | 0·65 | 0·61 | 0·65 (± 0·02) | 0·65 | 0·5590 |
| MECOM | **0·76** | **0·78 (± 0·04)** | **0·78** | | | | |
| MSI | **0·84** | **0·93 (± 0·01)** | **0·94** | **0·87** | **0·91 (± 0·02)** | **0·91** | 0·0015 |
| NRAS | 0·52 | 0·56 (± 0·04) | 0·55 | | | | |
| PIK3CA | 0·61 | 0·56 (± 0·03) | 0·55 | | | | |
| PLEKHA6 | 0·74 | **0·90 (± 0·01)** | **0·90** | | | | |
| RFX5 | **0·79** | **0·77 (± 0·02)** | **0·77** | | | | |
| RNF43 | **0·81** | **0·86 (± 0·01)** | **0·86** | **0·80** | **0·80 (± 0·05)** | **0·83** | 0·0021 |
| SMAD2 | 0·50 | 0·65 (± 0·03) | 0·65 | | | | |
| SMG1 | 0·58 | **0·76 (± 0·07)** | **0·78** | | | | |
| TBX3 | 0·58 | 0·74 (± 0·03) | 0·73 | | | | |
| TGFBR2 | 0·67 | **0·77 (± 0·02)** | **0·77** | | | | |
| TP53 | 0·65 | 0·72 (± 0·02) | 0·72 | 0·64 | 0·69 (± 0·05) | 0·70 | 0·3747 |
| TRPS1 | 0·63 | 0·73 (± 0·03) | 0·72 | | | | |
| WNT16 | **0·75** | **0·82 (± 0·02)** | **0·82** | | | | |
| ZHX2 | 0·66 | 0·74 (± 0·02) | 0·74 | | | | |
| ZNRF3 | **0·78** | **0·75 (± 0·01)** | **0·75** | 0·67 | 0·67 (± 0·04) | 0·68 | 0·0042 |

**Tab. S5: Performance metrics of Multi-Target Transformers including MSI as a target (primary model) for external validation on CRA.** Results are presented as mean and standard deviation across seven cross-validation folds for selected prediction targets. Metrics include Matthews Correlation Coefficient (MCC), Area Under the Receiver Operating Characteristic Curve (AUROC), and Area Under the Precision-Recall Curve (AUPRC), with mutation rates in the cohort. Binary classification thresholds were pre-defined at 0·5. Data is sorted by AUROC.

| **Target** | **Accuracy** | **Precision** | **Sensitivity** | **Specificity** | **F1 Score** | **MCC** | **AUROC** | **AUPRC** | **Mutation Rate** | **(Target MUT + MSI) / Target MUT** |
|---|---|---|---|---|---|---|---|---|---|---|
| MSI | 0·85 (±0·05) | 0·61 (±0·11) | 0·85 (±0·09) | 0·85 (±0·08) | 0·70 (±0·06) | 0·63 (±0·08) | 0·92 (±0·02) | 0·75 (±0·06) | 0·20 | 1·00 |
| hypermutated | 0·79 (±0·08) | 0·52 (±0·10) | 0·88 (±0·06) | 0·77 (±0·11) | 0·65 (±0·07) | 0·56 (±0·08) | 0·90 (±0·01) | 0·73 (±0·05) | 0·21 | 0·97 |
| PLEKHA6 | 0·77 (±0·05) | 0·28 (±0·04) | 0·91 (±0·08) | 0·76 (±0·06) | 0·42 (±0·04) | 0·42 (±0·03) | 0·90 (±0·01) | 0·38 (±0·06) | 0·09 | 0·97 |
| BMPR2 | 0·80 (±0·06) | 0·41 (±0·07) | 0·87 (±0·10) | 0·78 (±0·08) | 0·55 (±0·06) | 0·50 (±0·06) | 0·89 (±0·02) | 0·48 (±0·02) | 0·14 | 0·98 |
| CHD1 | 0·76 (±0·07) | 0·12 (±0·03) | 0·84 (±0·15) | 0·76 (±0·08) | 0·20 (±0·03) | 0·26 (±0·02) | 0·87 (±0·02) | 0·17 (±0·06) | 0·03 | 0·91 |
| RNF43 | 0·80 (±0·04) | 0·45 (±0·06) | 0·74 (±0·10) | 0·81 (±0·06) | 0·55 (±0·03) | 0·46 (±0·04) | 0·87 (±0·02) | 0·48 (±0·02) | 0·17 | 0·81 |
| FHOD3 | 0·79 (±0·06) | 0·33 (±0·06) | 0·77 (±0·14) | 0·79 (±0·08) | 0·45 (±0·04) | 0·40 (±0·04) | 0·84 (±0·02) | 0·38 (±0·03) | 0·11 | 0·81 |
| BRAF | 0·80 (±0·02) | 0·59 (±0·07) | 0·66 (±0·10) | 0·84 (±0·05) | 0·61 (±0·03) | 0·48 (±0·02) | 0·83 (±0·02) | 0·61 (±0·03) | 0·25 | 0·65 |
| CDK12 | 0·64 (±0·09) | 0·09 (±0·01) | 0·84 (±0·13) | 0·63 (±0·10) | 0·17 (±0·02) | 0·20 (±0·04) | 0·82 (±0·03) | 0·18 (±0·06) | 0·04 | 0·86 |
| WNT16 | 0·77 (±0·08) | 0·14 (±0·04) | 0·76 (±0·14) | 0·77 (±0·09) | 0·24 (±0·05) | 0·26 (±0·04) | 0·82 (±0·02) | 0·18 (±0·02) | 0·04 | 0·93 |
| SMG1 | 0·67 (±0·11) | 0·10 (±0·03) | 0·80 (±0·16) | 0·67 (±0·12) | 0·17 (±0·04) | 0·20 (±0·04) | 0·81 (±0·03) | 0·14 (±0·02) | 0·04 | 0·77 |
| ZNRF3 | 0·77 (±0·04) | 0·28 (±0·04) | 0·75 (±0·06) | 0·77 (±0·05) | 0·41 (±0·04) | 0·36 (±0·04) | 0·81 (±0·02) | 0·32 (±0·03) | 0·10 | 0·91 |
| AKT1 | 0·57 (±0·22) | 0·08 (±0·04) | 0·82 (±0·14) | 0·57 (±0·23) | 0·14 (±0·07) | 0·16 (±0·11) | 0·79 (±0·08) | 0·14 (±0·05) | 0·03 | 0·82 |
| ELL2 | 0·71 (±0·08) | 0·03 (±0·00) | 0·75 (±0·14) | 0·71 (±0·09) | 0·06 (±0·01) | 0·11 (±0·01) | 0·79 (±0·05) | 0·06 (±0·01) | 0·01 | 0·75 |
| DUSP16 | 0·46 (±0·21) | 0·05 (±0·02) | 0·89 (±0·13) | 0·45 (±0·22) | 0·09 (±0·03) | 0·12 (±0·06) | 0·78 (±0·08) | 0·11 (±0·04) | 0·03 | 0·78 |
| MECOM | 0·75 (±0·05) | 0·20 (±0·04) | 0·70 (±0·09) | 0·75 (±0·06) | 0·31 (±0·05) | 0·28 (±0·07) | 0·78 (±0·04) | 0·21 (±0·04) | 0·08 | 0·88 |
| RFX5 | 0·71 (±0·09) | 0·11 (±0·02) | 0·77 (±0·11) | 0·71 (±0·10) | 0·18 (±0·03) | 0·21 (±0·04) | 0·77 (±0·02) | 0·12 (±0·01) | 0·04 | 0·85 |
| ACVR1B | 0·63 (±0·12) | 0·09 (±0·02) | 0·73 (±0·09) | 0·62 (±0·13) | 0·16 (±0·03) | 0·16 (±0·05) | 0·74 (±0·04) | 0·22 (±0·06) | 0·05 | 0·53 |
| CTNND1 | 0·70 (±0·08) | 0·08 (±0·02) | 0·68 (±0·05) | 0·71 (±0·08) | 0·14 (±0·03) | 0·15 (±0·05) | 0·73 (±0·06) | 0·11 (±0·03) | 0·03 | 0·82 |
| ALK | 0·46 (±0·24) | 0·13 (±0·04) | 0·86 (±0·12) | 0·43 (±0·27) | 0·22 (±0·05) | 0·17 (±0·08) | 0·73 (±0·05) | 0·22 (±0·05) | 0·08 | 0·73 |
| TGFBR2 | 0·65 (±0·10) | 0·10 (±0·01) | 0·69 (±0·17) | 0·65 (±0·11) | 0·18 (±0·02) | 0·16 (±0·04) | 0·73 (±0·03) | 0·14 (±0·03) | 0·05 | 0·65 |
| TRPS1 | 0·48 (±0·17) | 0·14 (±0·05) | 0·80 (±0·07) | 0·45 (±0·20) | 0·23 (±0·07) | 0·15 (±0·09) | 0·73 (±0·03) | 0·24 (±0·05) | 0·09 | 0·68 |
| CCDC40 | 0·65 (±0·12) | 0·05 (±0·01) | 0·67 (±0·20) | 0·65 (±0·13) | 0·10 (±0·02) | 0·11 (±0·04) | 0·72 (±0·07) | 0·08 (±0·05) | 0·03 | 0·67 |
| TBX3 | 0·59 (±0·13) | 0·11 (±0·01) | 0·75 (±0·17) | 0·58 (±0·14) | 0·18 (±0·02) | 0·17 (±0·03) | 0·72 (±0·04) | 0·15 (±0·05) | 0·06 | 0·68 |
| APC | 0·68 | 0·81 | 0·75 | 0·52 | 0·75 | 0·28 | 0·71 | 0·85 | 0·72 | 0·12 |

| | | | | | | | | | | |
|---|---|---|---|---|---|---|---|---|---|---|
| | (±0·10) | (±0·05) | (±0·22) | (±0·24) | (±0·14) | (±0·05) | (±0·02) | (±0·01) | | |
| TP53 | 0·62 (±0·03) | 0·78 (±0·05) | 0·61 (±0·16) | 0·65 (±0·22) | 0·67 (±0·07) | 0·26 (±0·03) | 0·69 (±0·02) | 0·79 (±0·03) | 0·66 | 0·10 |
| KIF1A | 0·47 (±0·19) | 0·16 (±0·03) | 0·80 (±0·15) | 0·43 (±0·23) | 0·26 (±0·03) | 0·16 (±0·06) | 0·67 (±0·04) | 0·22 (±0·06) | 0·11 | 0·57 |
| ZHX2 | 0·59 (±0·08) | 0·04 (±0·01) | 0·62 (±0·23) | 0·59 (±0·09) | 0·07 (±0·02) | 0·07 (±0·05) | 0·67 (±0·03) | 0·07 (±0·05) | 0·02 | 0·62 |
| KRAS | 0·59 (±0·06) | 0·46 (±0·07) | 0·53 (±0·24) | 0·62 (±0·21) | 0·46 (±0·11) | 0·17 (±0·07) | 0·65 (±0·04) | 0·51 (±0·05) | 0·36 | 0·08 |
| ATM | 0·65 (±0·08) | 0·16 (±0·03) | 0·57 (±0·10) | 0·66 (±0·10) | 0·25 (±0·03) | 0·15 (±0·05) | 0·63 (±0·02) | 0·19 (±0·03) | 0·10 | 0·62 |
| PIK3CA | 0·55 (±0·10) | 0·24 (±0·02) | 0·66 (±0·20) | 0·52 (±0·17) | 0·34 (±0·04) | 0·15 (±0·06) | 0·62 (±0·03) | 0·25 (±0·02) | 0·18 | 0·39 |
| SMAD2 | 0·44 (±0·16) | 0·06 (±0·01) | 0·70 (±0·15) | 0·42 (±0·17) | 0·11 (±0·02) | 0·06 (±0·05) | 0·61 (±0·06) | 0·09 (±0·04) | 0·05 | 0·33 |
| NRAS | 0·48 (±0·16) | 0·05 (±0·01) | 0·53 (±0·20) | 0·48 (±0·17) | 0·09 (±0·02) | 0·00 (±0·05) | 0·51 (±0·06) | 0·06 (±0·02) | 0·05 | 0·00 |

**Tab. S6: Performance metrics of Multi-Target Transformers including MSI as a target (primary model) for external validation on WHI.** Results are presented as mean and standard deviation across seven cross-validation folds for selected prediction targets. Metrics include Matthews Correlation Coefficient (MCC), Area Under the Receiver Operating Characteristic Curve (AUROC), and Area Under the Precision-Recall Curve (AUPRC), with mutation rates in the cohort. Binary classification thresholds were pre-defined at 0·5. Data is sorted by AUROC.

| **Target** | **Accuracy** | **Precision** | **Sensitivity** | **Specificity** | **F1 Score** | **MCC** | **AUROC** | **AUPRC** | **Mutation Rate** | **(Target MUT + MSI) / Target MUT** |
|---|---|---|---|---|---|---|---|---|---|---|
| MSI | 0·84 (±0·04) | 0·73 (±0·07) | 0·91 (±0·04) | 0·81 (±0·07) | 0·80 (±0·03) | 0·69 (±0·05) | 0·94 (±0·01) | 0·91 (±0·02) | 0·35 | 1.00 |
| hypermutated | 0·78 (±0·02) | 0·64 (±0·04) | 0·84 (±0·03) | 0·75 (±0·05) | 0·72 (±0·02) | 0·56 (±0·03) | 0·86 (±0·01) | 0·76 (±0·02) | 0·34 | 0·89 |
| RNF43 | 0·75 (±0·04) | 0·53 (±0·05) | 0·86 (±0·03) | 0·70 (±0·07) | 0·66 (±0·03) | 0·51 (±0·04) | 0·84 (±0·01) | 0·65 (±0·02) | 0·28 | 0·89 |
| BMPR2 | 0·71 (±0·05) | 0·39 (±0·04) | 0·90 (±0·07) | 0·67 (±0·07) | 0·54 (±0·02) | 0·45 (±0·02) | 0·84 (±0·01) | 0·46 (±0·03) | 0·19 | 0·97 |
| TGFBR2 | 0·63 (±0·05) | 0·17 (±0·01) | 0·88 (±0·08) | 0·60 (±0·06) | 0·28 (±0·01) | 0·27 (±0·02) | 0·79 (±0·01) | 0·21 (±0·02) | 0·08 | 0·78 |
| CCDC40 | 0·60 (±0·10) | 0·08 (±0·01) | 0·83 (±0·23) | 0·59 (±0·12) | 0·15 (±0·02) | 0·17 (±0·05) | 0·78 (±0·03) | 0·15 (±0·07) | 0·04 | 0·93 |
| TBX3 | 0·47 (±0·16) | 0·09 (±0·02) | 0·91 (±0·05) | 0·44 (±0·17) | 0·17 (±0·03) | 0·17 (±0·06) | 0·78 (±0·03) | 0·17 (±0·05) | 0·06 | 0·83 |
| ZHX2 | 0·61 (±0·05) | 0·09 (±0·01) | 0·91 (±0·09) | 0·60 (±0·05) | 0·17 (±0·01) | 0·21 (±0·03) | 0·78 (±0·00) | 0·11 (±0·01) | 0·04 | 0·86 |
| BRAF | 0·66 (±0·04) | 0·32 (±0·02) | 0·81 (±0·06) | 0·63 (±0·06) | 0·45 (±0·02) | 0·34 (±0·02) | 0·77 (±0·01) | 0·41 (±0·04) | 0·17 | 0·80 |
| ACVR1B | 0·60 (±0·06) | 0·13 (±0·01) | 0·80 (±0·17) | 0·58 (±0·07) | 0·23 (±0·02) | 0·20 (±0·05) | 0·75 (±0·04) | 0·17 (±0·03) | 0·07 | 0·50 |
| CDK12 | 0·56 (±0·08) | 0·09 (±0·01) | 0·89 (±0·08) | 0·54 (±0·09) | 0·16 (±0·02) | 0·18 (±0·03) | 0·75 (±0·03) | 0·13 (±0·02) | 0·05 | 0·87 |
| CTNND1 | 0·63 (±0·06) | 0·11 (±0·02) | 0·79 (±0·08) | 0·62 (±0·07) | 0·19 (±0·03) | 0·19 (±0·04) | 0·75 (±0·02) | 0·11 (±0·02) | 0·05 | 0·94 |
| TP53 | 0·67 (±0·03) | 0·60 (±0·05) | 0·62 (±0·16) | 0·70 (±0·15) | 0·59 (±0·04) | 0·33 (±0·03) | 0·73 (±0·01) | 0·64 (±0·03) | 0·40 | 0·18 |
| SMG1 | 0·48 (±0·18) | 0·07 (±0·02) | 0·85 (±0·17) | 0·47 (±0·19) | 0·12 (±0·04) | 0·13 (±0·08) | 0·72 (±0·10) | 0·10 (±0·04) | 0·04 | 0·77 |
| DUSP16 | 0·45 (±0·18) | 0·03 (±0·01) | 0·76 (±0·14) | 0·44 (±0·19) | 0·06 (±0·01) | 0·06 (±0·03) | 0·70 (±0·05) | 0·09 (±0·06) | 0·02 | 0·71 |
| SMAD2 | 0·38 (±0·19) | 0·08 (±0·01) | 0·88 (±0·09) | 0·35 (±0·20) | 0·14 (±0·02) | 0·11 (±0·05) | 0·70 (±0·02) | 0·14 (±0·04) | 0·06 | 0·44 |
| ZNRF3 | 0·62 (±0·05) | 0·19 (±0·02) | 0·71 (±0·05) | 0·61 (±0·07) | 0·30 (±0·02) | 0·21 (±0·03) | 0·70 (±0·01) | 0·23 (±0·02) | 0·11 | 0·84 |
| ALK | 0·39 (±0·23) | 0·11 (±0·02) | 0·83 (±0·17) | 0·35 (±0·26) | 0·19 (±0·03) | 0·11 (±0·06) | 0·68 (±0·01) | 0·16 (±0·02) | 0·08 | 0·81 |
| KIF1A | 0·45 (±0·20) | 0·12 (±0·02) | 0·80 (±0·15) | 0·42 (±0·23) | 0·21 (±0·03) | 0·13 (±0·05) | 0·67 (±0·02) | 0·19 (±0·02) | 0·09 | 0·71 |
| KRAS | 0·62 (±0·05) | 0·42 (±0·04) | 0·61 (±0·13) | 0·63 (±0·12) | 0·49 (±0·04) | 0·23 (±0·04) | 0·66 (±0·02) | 0·43 (±0·03) | 0·30 | 0·08 |
| AKT1 | 0·39 (±0·16) | 0·08 (±0·01) | 0·80 (±0·16) | 0·37 (±0·18) | 0·14 (±0·01) | 0·09 (±0·03) | 0·64 (±0·02) | 0·09 (±0·01) | 0·06 | 0·53 |
| ATM | 0·57 (±0·10) | 0·13 (±0·02) | 0·68 (±0·06) | 0·55 (±0·11) | 0·21 (±0·03) | 0·13 (±0·05) | 0·64 (±0·02) | 0·14 (±0·02) | 0·08 | 0·63 |
| APC | 0·57 (±0·03) | 0·53 (±0·03) | 0·59 (±0·24) | 0·56 (±0·19) | 0·53 (±0·18) | 0·15 (±0·07) | 0·60 (±0·02) | 0·52 (±0·02) | 0·46 | 0·21 |
| NRAS | 0·69 (±0·16) | 0·05 (±0·02) | 0·44 (±0·25) | 0·70 (±0·18) | 0·08 (±0·04) | 0·05 (±0·05) | 0·59 (±0·05) | 0·09 (±0·06) | 0·04 | 0·17 |
| PIK3CA | 0·43 | 0·15 | 0·66 | 0·39 | 0·25 | 0·04 | 0·51 | 0·15 | 0·14 | 0·46 |

| | | | | | | | | |
|---|---|---|---|---|---|---|---|---|
| | (±0·09) | (±0·01) | (±0·11) | (±0·12) | (±0·01) | (±0·03) | (±0·03) | (±0·02) |

**Tab. S7: Performance metrics of Multi-Target Transformers including MSI as a target (primary model) for external validation on TCGA.** Results are presented as mean and standard deviation across seven cross-validation folds for selected prediction targets. Metrics include Matthews Correlation Coefficient (MCC), Area Under the Receiver Operating Characteristic Curve (AUROC), and Area Under the Precision-Recall Curve (AUPRC), with mutation rates in the cohort. Binary classification thresholds were pre-defined at 0·5. Data is sorted by AUROC.

| **Target** | **Accuracy** | **Precision** | **Sensitivity** | **Specificity** | **F1 Score** | **MCC** | **AUROC** | **AUPRC** | **Mutation Rate** | **(Target MUT + MSI) / Target MUT** |
|---|---|---|---|---|---|---|---|---|---|---|
| hypermutated | 0·77 (±0·08) | 0·42 (±0·09) | 0·84 (±0·07) | 0·75 (±0·11) | 0·55 (±0·06) | 0·47 (±0·06) | 0·89 (±0·01) | 0·63 (±0·02) | 0·16 | 0·85 |
| MSI | 0·78 (±0·09) | 0·40 (±0·10) | 0·80 (±0·07) | 0·77 (±0·11) | 0·52 (±0·07) | 0·46 (±0·07) | 0·87 (±0·01) | 0·62 (±0·02) | 0·14 | 1.00 |
| FHOD3 | 0·77 (±0·09) | 0·23 (±0·06) | 0·76 (±0·14) | 0·77 (±0·10) | 0·34 (±0·06) | 0·32 (±0·05) | 0·85 (±0·01) | 0·34 (±0·04) | 0·07 | 0·65 |
| WNT16 | 0·75 (±0·07) | 0·05 (±0·01) | 0·79 (±0·13) | 0·75 (±0·07) | 0·09 (±0·03) | 0·15 (±0·04) | 0·84 (±0·03) | 0·10 (±0·06) | 0·01 | 0·67 |
| ZHX2 | 0·73 (±0·04) | 0·09 (±0·01) | 0·82 (±0·11) | 0·72 (±0·04) | 0·16 (±0·03) | 0·21 (±0·04) | 0·83 (±0·05) | 0·16 (±0·06) | 0·03 | 0·69 |
| ELL2 | 0·73 (±0·08) | 0·07 (±0·01) | 0·80 (±0·08) | 0·73 (±0·09) | 0·13 (±0·03) | 0·18 (±0·04) | 0·82 (±0·03) | 0·15 (±0·07) | 0·02 | 0·40 |
| BMPR2 | 0·74 (±0·09) | 0·20 (±0·05) | 0·75 (±0·12) | 0·74 (±0·11) | 0·30 (±0·04) | 0·29 (±0·04) | 0·82 (±0·01) | 0·31 (±0·03) | 0·07 | 0·84 |
| PLEKHA6 | 0·72 (±0·10) | 0·15 (±0·05) | 0·78 (±0·05) | 0·71 (±0·11) | 0·25 (±0·06) | 0·25 (±0·06) | 0·80 (±0·03) | 0·22 (±0·04) | 0·06 | 0·96 |
| RNF43 | 0·73 (±0·08) | 0·23 (±0·05) | 0·72 (±0·09) | 0·74 (±0·10) | 0·35 (±0·05) | 0·30 (±0·04) | 0·80 (±0·01) | 0·33 (±0·02) | 0·09 | 0·75 |
| MECOM | 0·73 (±0·09) | 0·14 (±0·03) | 0·75 (±0·08) | 0·73 (±0·10) | 0·24 (±0·04) | 0·25 (±0·04) | 0·79 (±0·03) | 0·17 (±0·03) | 0·05 | 0·61 |
| BRAF | 0·73 (±0·07) | 0·30 (±0·07) | 0·72 (±0·07) | 0·74 (±0·09) | 0·42 (±0·05) | 0·34 (±0·06) | 0·78 (±0·02) | 0·40 (±0·03) | 0·13 | 0·64 |
| ZNRF3 | 0·73 (±0·06) | 0·08 (±0·02) | 0·71 (±0·06) | 0·73 (±0·06) | 0·14 (±0·03) | 0·17 (±0·04) | 0·78 (±0·02) | 0·12 (±0·05) | 0·03 | 0·85 |
| RFX5 | 0·72 (±0·11) | 0·09 (±0·03) | 0·65 (±0·13) | 0·72 (±0·12) | 0·15 (±0·03) | 0·16 (±0·03) | 0·75 (±0·02) | 0·23 (±0·07) | 0·04 | 0·80 |
| CDK12 | 0·66 (±0·09) | 0·12 (±0·03) | 0·67 (±0·10) | 0·66 (±0·10) | 0·21 (±0·03) | 0·17 (±0·03) | 0·74 (±0·03) | 0·21 (±0·05) | 0·06 | 0·59 |
| TGFBR2 | 0·67 (±0·09) | 0·08 (±0·02) | 0·68 (±0·06) | 0·67 (±0·10) | 0·13 (±0·04) | 0·14 (±0·04) | 0·74 (±0·03) | 0·12 (±0·02) | 0·04 | 0·60 |
| DUSP16 | 0·52 (±0·20) | 0·00 (±0·00) | 0·86 (±0·38) | 0·52 (±0·20) | 0·01 (±0·01) | 0·04 (±0·04) | 0·73 (±0·18) | 0·01 (±0·01) | 0.00 | 1.00 |
| TP53 | 0·63 (±0·06) | 0·79 (±0·04) | 0·59 (±0·15) | 0·70 (±0·15) | 0·67 (±0·08) | 0·29 (±0·07) | 0·72 (±0·03) | 0·81 (±0·03) | 0·65 | 0·08 |
| ACVR1B | 0·67 (±0·12) | 0·10 (±0·03) | 0·67 (±0·10) | 0·67 (±0·13) | 0·18 (±0·04) | 0·16 (±0·04) | 0·72 (±0·03) | 0·15 (±0·07) | 0·05 | 0·48 |
| ALK | 0·45 (±0·29) | 0·11 (±0·05) | 0·82 (±0·15) | 0·43 (±0·32) | 0·19 (±0·07) | 0·14 (±0·09) | 0·72 (±0·03) | 0·19 (±0·03) | 0·07 | 0·50 |
| CTNND1 | 0·69 (±0·11) | 0·10 (±0·03) | 0·65 (±0·06) | 0·69 (±0·12) | 0·17 (±0·04) | 0·16 (±0·04) | 0·72 (±0·02) | 0·14 (±0·03) | 0·05 | 0·40 |
| CCDC40 | 0·70 (±0·13) | 0·08 (±0·02) | 0·54 (±0·12) | 0·71 (±0·14) | 0·13 (±0·03) | 0·11 (±0·04) | 0·68 (±0·02) | 0·10 (±0·03) | 0·04 | 0·44 |
| APC | 0·68 (±0·11) | 0·85 (±0·01) | 0·73 (±0·17) | 0·51 (±0·15) | 0·77 (±0·11) | 0·23 (±0·06) | 0·67 (±0·01) | 0·86 (±0·01) | 0·79 | 0·08 |
| CHD1 | 0·72 (±0·07) | 0·09 (±0·01) | 0·49 (±0·11) | 0·73 (±0·08) | 0·16 (±0·01) | 0·11 (±0·02) | 0·66 (±0·02) | 0·09 (±0·01) | 0·05 | 0·55 |
| SMG1 | 0·65 (±0·17) | 0·08 (±0·03) | 0·59 (±0·18) | 0·66 (±0·18) | 0·13 (±0·05) | 0·11 (±0·08) | 0·64 (±0·11) | 0·10 (±0·04) | 0·04 | 0·50 |
| ATM | 0·66 | 0·22 | 0·55 | 0·68 | 0·30 | 0·17 | 0·64 | 0·24 | 0·13 | 0·37 |

| | | | | | | | | | | |
|---|---|---|---|---|---|---|---|---|---|---|
| | (±0·10) | (±0·03) | (±0·12) | (±0·13) | (±0·03) | (±0·04) | (±0·02) | (±0·03) | | |
| NRAS | 0·54 (±0·20) | 0·09 (±0·04) | 0·65 (±0·32) | 0·54 (±0·24) | 0·15 (±0·07) | 0·09 (±0·06) | 0·63 (±0·06) | 0·11 (±0·03) | 0·07 | 0·10 |
| SMAD2 | 0·46 (±0·19) | 0·07 (±0·01) | 0·72 (±0·16) | 0·44 (±0·21) | 0·13 (±0·02) | 0·08 (±0·03) | 0·63 (±0·05) | 0·09 (±0·02) | 0·05 | 0·09 |
| KIF1A | 0·55 (±0·24) | 0·09 (±0·02) | 0·62 (±0·21) | 0·55 (±0·27) | 0·16 (±0·03) | 0·09 (±0·05) | 0·63 (±0·03) | 0·11 (±0·03) | 0·06 | 0·41 |
| TRPS1 | 0·58 (±0·12) | 0·16 (±0·03) | 0·62 (±0·14) | 0·58 (±0·15) | 0·24 (±0·02) | 0·13 (±0·03) | 0·63 (±0·03) | 0·19 (±0·04) | 0·11 | 0·36 |
| AKT1 | 0·60 (±0·16) | 0·03 (±0·01) | 0·59 (±0·17) | 0·60 (±0·17) | 0·06 (±0·02) | 0·06 (±0·06) | 0·62 (±0·11) | 0·06 (±0·04) | 0·02 | 0·67 |
| TBX3 | 0·61 (±0·09) | 0·06 (±0·01) | 0·54 (±0·10) | 0·62 (±0·10) | 0·11 (±0·02) | 0·07 (±0·04) | 0·59 (±0·03) | 0·08 (±0·02) | 0·04 | 0·42 |
| PIK3CA | 0·47 (±0·08) | 0·31 (±0·01) | 0·71 (±0·16) | 0·38 (±0·16) | 0·43 (±0·03) | 0·09 (±0·04) | 0·58 (±0·03) | 0·35 (±0·02) | 0·28 | 0·20 |
| KRAS | 0·53 (±0·03) | 0·48 (±0·04) | 0·52 (±0·19) | 0·55 (±0·20) | 0·48 (±0·10) | 0·07 (±0·05) | 0·56 (±0·02) | 0·52 (±0·03) | 0·44 | 0·10 |

**Tab. S8: Performance metrics of Multi-Target Transformers including MSI as a target (primary model) for external validation on CPTAC.** Results are presented as mean and standard deviation across seven cross-validation folds for selected prediction targets. Metrics include Matthews Correlation Coefficient (MCC), Area Under the Receiver Operating Characteristic Curve (AUROC), and Area Under the Precision-Recall Curve (AUPRC), with mutation rates in the cohort. Binary classification thresholds were pre-defined at 0·5. Data is sorted by AUROC.

| Target | Accuracy | Precision | Sensitivity | Specificity | F1 Score | MCC | AUROC | AUPRC | Mutation Rate | (Target MUT + MSI) / Target MUT |
|---|---|---|---|---|---|---|---|---|---|---|
| MSI | 0·51 (±0·12) | 0·32 (±0·06) | 0·99 (±0·02) | 0·36 (±0·16) | 0·48 (±0·07) | 0·33 (±0·11) | 0·90 (±0·03) | 0·75 (±0·05) | 0·23 | 1.00 |
| CCDC40 | 0·39 (±0·25) | 0·11 (±0·04) | 0·98 (±0·05) | 0·35 (±0·27) | 0·19 (±0·07) | 0·18 (±0·11) | 0·80 (±0·06) | 0·26 (±0·08) | 0·07 | 1.00 |
| RNF43 | 0·32 (±0·08) | 0·18 (±0·02) | 0·98 (±0·05) | 0·20 (±0·10) | 0·30 (±0·03) | 0·17 (±0·07) | 0·80 (±0·03) | 0·47 (±0·06) | 0·15 | 0·88 |
| BRAF | 0·36 (±0·12) | 0·18 (±0·03) | 0·88 (±0·06) | 0·27 (±0·15) | 0·30 (±0·03) | 0·13 (±0·06) | 0·80 (±0·02) | 0·53 (±0·08) | 0·15 | 0·81 |
| TGFBR2 | 0·34 (±0·17) | 0·08 (±0·02) | 0·95 (±0·08) | 0·31 (±0·19) | 0·15 (±0·03) | 0·14 (±0·06) | 0·79 (±0·08) | 0·25 (±0·09) | 0·06 | 1.00 |
| FHOD3 | 0·42 (±0·16) | 0·16 (±0·04) | 0·89 (±0·10) | 0·36 (±0·19) | 0·27 (±0·04) | 0·18 (±0·06) | 0·78 (±0·01) | 0·29 (±0·02) | 0·11 | 0·92 |
| PLEKHA6 | 0·25 (±0·11) | 0·09 (±0·01) | 0·96 (±0·06) | 0·20 (±0·12) | 0·17 (±0·02) | 0·10 (±0·10) | 0·76 (±0·10) | 0·22 (±0·06) | 0·08 | 0·88 |
| BMPR2 | 0·37 (±0·08) | 0·16 (±0·02) | 0·99 (±0·03) | 0·28 (±0·10) | 0·28 (±0·02) | 0·20 (±0·05) | 0·76 (±0·03) | 0·27 (±0·02) | 0·12 | 0·85 |
| ACVR1B | 0·25 (±0·07) | 0·03 (±0·01) | 0·95 (±0·13) | 0·22 (±0·07) | 0·07 (±0·01) | 0·07 (±0·06) | 0·75 (±0·08) | 0·29 (±0·22) | 0·03 | 0·67 |
| RFX5 | 0·33 (±0·22) | 0·07 (±0·04) | 0·94 (±0·10) | 0·30 (±0·23) | 0·13 (±0·06) | 0·12 (±0·08) | 0·75 (±0·05) | 0·17 (±0·06) | 0·05 | 1.00 |
| KIF1A | 0·20 (±0·11) | 0·09 (±0·01) | 0·98 (±0·05) | 0·14 (±0·13) | 0·16 (±0·02) | 0·09 (±0·05) | 0·75 (±0·05) | 0·23 (±0·06) | 0·08 | 0·88 |
| TBX3 | 0·25 (±0·18) | 0·10 (±0·03) | 1·00 (±0·00) | 0·19 (±0·20) | 0·18 (±0·04) | 0·12 (±0·10) | 0·74 (±0·10) | 0·25 (±0·10) | 0·08 | 0·62 |
| DUSP16 | 0·22 (±0·17) | 0·07 (±0·01) | 0·98 (±0·06) | 0·18 (±0·18) | 0·13 (±0·02) | 0·09 (±0·07) | 0·73 (±0·04) | 0·15 (±0·06) | 0·06 | 0·33 |
| WNT16 | 0·35 (±0·15) | 0·12 (±0·02) | 0·89 (±0·09) | 0·29 (±0·18) | 0·21 (±0·04) | 0·12 (±0·07) | 0·73 (±0·03) | 0·23 (±0·02) | 0·09 | 0·80 |
| ALK | 0·23 (±0·19) | 0·09 (±0·02) | 0·91 (±0·16) | 0·17 (±0·22) | 0·16 (±0·02) | 0·06 (±0·06) | 0·72 (±0·09) | 0·23 (±0·11) | 0·08 | 0·75 |
| CDK12 | 0·26 (±0·15) | 0·04 (±0·01) | 1·00 (±0·00) | 0·24 (±0·15) | 0·07 (±0·02) | 0·09 (±0·04) | 0·71 (±0·09) | 0·10 (±0·02) | 0·03 | 0·67 |
| MECOM | 0·32 (±0·11) | 0·10 (±0·01) | 0·96 (±0·06) | 0·27 (±0·12) | 0·18 (±0·02) | 0·14 (±0·03) | 0·71 (±0·09) | 0·18 (±0·08) | 0·08 | 0·88 |
| ZNRF3 | 0·32 (±0·12) | 0·10 (±0·02) | 0·83 (±0·06) | 0·27 (±0·13) | 0·17 (±0·03) | 0·06 (±0·06) | 0·71 (±0·03) | 0·30 (±0·06) | 0·08 | 0·78 |
| KRAS | 0·70 (±0·02) | 0·66 (±0·20) | 0·23 (±0·14) | 0·93 (±0·07) | 0·32 (±0·14) | 0·23 (±0·10) | 0·69 (±0·06) | 0·55 (±0·05) | 0·33 | 0.00 |
| SMG1 | 0·27 (±0·19) | 0·08 (±0·01) | 0·90 (±0·21) | 0·23 (±0·21) | 0·14 (±0·01) | 0·08 (±0·05) | 0·68 (±0·08) | 0·16 (±0·06) | 0·07 | 0·71 |
| CHD1 | 0·33 (±0·12) | 0·07 (±0·00) | 0·88 (±0·16) | 0·30 (±0·14) | 0·13 (±0·01) | 0·10 (±0·03) | 0·68 (±0·04) | 0·14 (±0·04) | 0·06 | 0·83 |
| APC | 0·46 (±0·14) | 0·83 (±0·04) | 0·35 (±0·20) | 0·81 (±0·08) | 0·46 (±0·20) | 0·15 (±0·13) | 0·67 (±0·04) | 0·83 (±0·02) | 0·75 | 0·14 |
| TRPS1 | 0·49 (±0·23) | 0·21 (±0·05) | 0·66 (±0·30) | 0·46 (±0·33) | 0·30 (±0·08) | 0·10 (±0·09) | 0·66 (±0·02) | 0·32 (±0·07) | 0·17 | 0·56 |
| PIK3CA | 0·45 (±0·15) | 0·25 (±0·04) | 0·78 (±0·23) | 0·35 (±0·24) | 0·38 (±0·06) | 0·12 (±0·12) | 0·64 (±0·04) | 0·38 (±0·07) | 0·22 | 0·43 |

| | | | | | | | | | | |
|---|---|---|---|---|---|---|---|---|---|---|
| SMAD2 | 0·15 (±0·04) | 0·10 (±0·00) | 1·00 (±0·00) | 0·06 (±0·04) | 0·18 (±0·01) | 0·07 (±0·04) | 0·62 (±0·05) | 0·15 (±0·03) | 0·09 | 0·40 |
| ATM | 0·28 (±0·11) | 0·08 (±0·01) | 0·88 (±0·22) | 0·23 (±0·14) | 0·15 (±0·02) | 0·08 (±0·08) | 0·59 (±0·08) | 0·12 (±0·03) | 0·08 | 0·50 |
| TP53 | 0·54 (±0·03) | 0·58 (±0·05) | 0·43 (±0·24) | 0·66 (±0·23) | 0·46 (±0·19) | 0·10 (±0·06) | 0·59 (±0·01) | 0·62 (±0·03) | 0·53 | 0·14 |
| AKT1 | 0·10 (±0·09) | 0·01 (±0·00) | 1·00 (±0·00) | 0·09 (±0·09) | 0·02 (±0·00) | 0·03 (±0·02) | 0·50 (±0·22) | 0·02 (±0·01) | 0·01 | 1.00 |
| ELL2 | 0·18 (±0·09) | 0·04 (±0·00) | 0·79 (±0·09) | 0·16 (±0·09) | 0·07 (±0·01) | -0·05 (±0·09) | 0·49 (±0·06) | 0·09 (±0·09) | 0·04 | 0·50 |
| NRAS | 0·81 (±0·16) | 0·03 (±0·05) | 0·07 (±0·13) | 0·85 (±0·17) | 0·03 (±0·06) | -0·05 (±0·07) | 0·48 (±0·13) | 0·07 (±0·03) | 0·06 | 0·17 |
| ZHX2 | 0·22 (±0·13) | 0·04 (±0·01) | 0·75 (±0·14) | 0·20 (±0·13) | 0·07 (±0·01) | -0·03 (±0·07) | 0·46 (±0·05) | 0·05 (±0·02) | 0·04 | 0·50 |
| CTNND1 | 0·38 (±0·18) | 0·01 (±0·01) | 0·43 (±0·45) | 0·38 (±0·19) | 0·02 (±0·02) | -0·05 (±0·08) | 0·39 (±0·07) | 0·02 (±0·00) | 0·02 | 0.00 |

**Tab. S9: Performance metrics of Multi-Target Transformers excluding MSI as a target (secondary model) for external validation on CRA.** Results are presented as mean and standard deviation across seven cross-validation folds for selected prediction targets. Metrics include Matthews Correlation Coefficient (MCC), Area Under the Receiver Operating Characteristic Curve (AUROC), and Area Under the Precision-Recall Curve (AUPRC), with mutation rates in the cohort. Binary classification thresholds were pre-defined at 0·5. DeLong p-values, as detailed in Materials and Methods, compare performance to the primary model (Tab. S5). Data is sorted by AUROC.

| Target | Accuracy | Precision | Sensitivity | Specificity | F1 Score | MCC | AUROC | AUPRC | Mutation Rate | (Target MUT + MSI) / Target MUT | DeLong test p-value |
|---|---|---|---|---|---|---|---|---|---|---|---|
| hypermutated | 0·82 (±0·05) | 0·58 (±0·11) | 0·83 (±0·17) | 0·82 (±0·09) | 0·66 (±0·05) | 0·58 (±0·05) | 0·91 (±0·01) | 0·74 (±0·04) | 0·21 | 0·97 | 0·1534 |
| PLEKHA6 | 0·79 (±0·09) | 0·31 (±0·08) | 0·85 (±0·15) | 0·79 (±0·11) | 0·44 (±0·07) | 0·43 (±0·07) | 0·90 (±0·02) | 0·38 (±0·05) | 0·09 | 0·97 | 0·6779 |
| CHD1 | 0·78 (±0·07) | 0·13 (±0·05) | 0·86 (±0·16) | 0·77 (±0·08) | 0·22 (±0·06) | 0·28 (±0·06) | 0·89 (±0·04) | 0·17 (±0·05) | 0·03 | 0·91 | 0·1341 |
| BMPR2 | 0·83 (±0·03) | 0·45 (±0·06) | 0·75 (±0·13) | 0·84 (±0·06) | 0·55 (±0·03) | 0·49 (±0·04) | 0·88 (±0·01) | 0·47 (±0·03) | 0·14 | 0·98 | 0·0929 |
| RNF43 | 0·81 (±0·02) | 0·46 (±0·03) | 0·65 (±0·14) | 0·85 (±0·05) | 0·53 (±0·05) | 0·44 (±0·05) | 0·85 (±0·01) | 0·48 (±0·03) | 0·17 | 0·81 | 0·0613 |
| FHOD3 | 0·81 (±0·04) | 0·35 (±0·05) | 0·73 (±0·13) | 0·82 (±0·06) | 0·47 (±0·03) | 0·41 (±0·04) | 0·84 (±0·01) | 0·39 (±0·03) | 0·11 | 0·81 | 0·3732 |
| AKT1 | 0·64 (±0·22) | 0·10 (±0·05) | 0·83 (±0·26) | 0·63 (±0·24) | 0·16 (±0·06) | 0·20 (±0·06) | 0·82 (±0·07) | 0·16 (±0·04) | 0·03 | 0·82 | 0·1237 |
| ELL2 | 0·77 (±0·08) | 0·05 (±0·02) | 0·79 (±0·17) | 0·77 (±0·08) | 0·09 (±0·03) | 0·15 (±0·03) | 0·82 (±0·06) | 0·05 (±0·01) | 0·01 | 0·75 | 0·5174 |
| BRAF | 0·79 (±0·01) | 0·57 (±0·02) | 0·64 (±0·08) | 0·84 (±0·03) | 0·60 (±0·03) | 0·47 (±0·03) | 0·82 (±0·03) | 0·61 (±0·04) | 0·25 | 0·65 | 0·1373 |
| WNT16 | 0·79 (±0·07) | 0·13 (±0·02) | 0·65 (±0·23) | 0·80 (±0·09) | 0·21 (±0·04) | 0·22 (±0·06) | 0·81 (±0·03) | 0·20 (±0·05) | 0·04 | 0·93 | 0·9363 |
| ZNRF3 | 0·77 (±0·06) | 0·27 (±0·05) | 0·68 (±0·09) | 0·77 (±0·07) | 0·38 (±0·04) | 0·32 (±0·05) | 0·80 (±0·02) | 0·32 (±0·02) | 0·1 | 0·91 | 0·0462 |
| RFX5 | 0·79 (±0·06) | 0·13 (±0·03) | 0·65 (±0·15) | 0·80 (±0·07) | 0·21 (±0·04) | 0·22 (±0·05) | 0·78 (±0·05) | 0·14 (±0·03) | 0·04 | 0·85 | 0·6139 |
| MECOM | 0·77 (±0·06) | 0·20 (±0·04) | 0·57 (±0·17) | 0·79 (±0·08) | 0·29 (±0·04) | 0·23 (±0·06) | 0·77 (±0·02) | 0·20 (±0·02) | 0·08 | 0·88 | 0·0605 |
| TRPS1 | 0·80 (±0·11) | 0·28 (±0·09) | 0·58 (±0·14) | 0·82 (±0·14) | 0·36 (±0·06) | 0·30 (±0·06) | 0·76 (±0·02) | 0·30 (±0·03) | 0·09 | 0·68 | 0·5339 |
| DUSP16 | 0·61 (±0·15) | 0·06 (±0·02) | 0·79 (±0·23) | 0·60 (±0·16) | 0·11 (±0·03) | 0·14 (±0·07) | 0·74 (±0·11) | 0·10 (±0·06) | 0·03 | 0·78 | 0·8470 |
| TBX3 | 0·62 (±0·23) | 0·12 (±0·03) | 0·70 (±0·21) | 0·62 (±0·26) | 0·20 (±0·04) | 0·17 (±0·06) | 0·74 (±0·04) | 0·15 (±0·04) | 0·06 | 0·68 | 0·8747 |
| TGFBR2 | 0·73 (±0·07) | 0·11 (±0·02) | 0·55 (±0·13) | 0·74 (±0·08) | 0·18 (±0·02) | 0·15 (±0·03) | 0·74 (±0·02) | 0·14 (±0·02) | 0·05 | 0·65 | 0·6833 |
| CDK12 | 0·69 (±0·10) | 0·10 (±0·03) | 0·69 (±0·20) | 0·69 (±0·10) | 0·17 (±0·05) | 0·18 (±0·10) | 0·73 (±0·14) | 0·16 (±0·08) | 0·04 | 0·86 | 0·0486 |
| SMG1 | 0·52 (±0·25) | 0·08 (±0·03) | 0·84 (±0·08) | 0·51 (±0·26) | 0·15 (±0·06) | 0·13 (±0·16) | 0·72 (±0·18) | 0·13 (±0·06) | 0·04 | 0·77 | 0·2523 |
| ALK | 0·69 (±0·14) | 0·17 (±0·03) | 0·64 (±0·28) | 0·70 (±0·17) | 0·25 (±0·04) | 0·20 (±0·06) | 0·72 (±0·07) | 0·20 (±0·02) | 0·08 | 0·73 | 0·6855 |
| ACVR1B | 0·73 (±0·06) | 0·11 (±0·02) | 0·65 (±0·06) | 0·73 (±0·06) | 0·18 (±0·03) | 0·18 (±0·04) | 0·72 (±0·03) | 0·23 (±0·05) | 0·05 | 0·53 | 0·0493 |
| APC | 0·68 (±0·09) | 0·81 (±0·05) | 0·75 (±0·20) | 0·52 (±0·22) | 0·76 (±0·12) | 0·28 (±0·05) | 0·71 (±0·03) | 0·85 (±0·02) | 0·72 | 0·12 | 0·1261 |
| KIF1A | 0·58 (±0·09) | 0·17 (±0·03) | 0·71 (±0·07) | 0·57 (±0·11) | 0·28 (±0·03) | 0·18 (±0·04) | 0·70 (±0·04) | 0·24 (±0·05) | 0·11 | 0·57 | 0·1216 |

| | | | | | | | | | | |
|---|---|---|---|---|---|---|---|---|---|---|
| ZHX2 | 0·63 (±0·15) | 0·04 (±0·01) | 0·62 (±0·12) | 0·63 (±0·16) | 0·08 (±0·02) | 0·09 (±0·03) | 0·68 (±0·03) | 0·06 (±0·02) | 0·02 | 0·62 | 0·6861 |
| TP53 | 0·58 (±0·10) | 0·80 (±0·06) | 0·51 (±0·25) | 0·71 (±0·21) | 0·58 (±0·21) | 0·23 (±0·05) | 0·68 (±0·02) | 0·79 (±0·02) | 0·66 | 0·10 | 0·2982 |
| CCDC40 | 0·67 (±0·09) | 0·05 (±0·01) | 0·57 (±0·22) | 0·68 (±0·10) | 0·09 (±0·03) | 0·09 (±0·06) | 0·67 (±0·08) | 0·06 (±0·01) | 0·03 | 0·67 | 0·1356 |
| CTNND1 | 0·73 (±0·14) | 0·07 (±0·02) | 0·52 (±0·17) | 0·74 (±0·15) | 0·13 (±0·04) | 0·12 (±0·05) | 0·67 (±0·06) | 0·10 (±0·02) | 0·03 | 0·82 | 0·0557 |
| ATM | 0·68 (±0·09) | 0·16 (±0·02) | 0·54 (±0·20) | 0·70 (±0·13) | 0·24 (±0·05) | 0·15 (±0·06) | 0·65 (±0·04) | 0·18 (±0·03) | 0·1 | 0·62 | 0·0495 |
| SMAD2 | 0·64 (±0·10) | 0·08 (±0·02) | 0·57 (±0·17) | 0·65 (±0·11) | 0·13 (±0·04) | 0·10 (±0·06) | 0·64 (±0·07) | 0·11 (±0·06) | 0·05 | 0·33 | 0·0421 |
| KRAS | 0·50 (±0·03) | 0·41 (±0·01) | 0·84 (±0·06) | 0·31 (±0·07) | 0·55 (±0·02) | 0·16 (±0·04) | 0·63 (±0·02) | 0·50 (±0·02) | 0·36 | 0·08 | 0·0804 |
| PIK3CA | 0·60 (±0·10) | 0·24 (±0·03) | 0·55 (±0·24) | 0·61 (±0·17) | 0·32 (±0·07) | 0·13 (±0·07) | 0·60 (±0·03) | 0·23 (±0·02) | 0·18 | 0·39 | 0·0303 |
| NRAS | 0·63 (±0·18) | 0·06 (±0·01) | 0·42 (±0·23) | 0·64 (±0·21) | 0·10 (±0·02) | 0·03 (±0·03) | 0·57 (±0·08) | 0·08 (±0·02) | 0·05 | 0.00 | 0·0003 |

**Tab. S10: Performance metrics of Multi-Target Transformers excluding MSI as a target (secondary model) for external validation on WHI.** Results are presented as mean and standard deviation across seven cross-validation folds for selected prediction targets. Metrics include Matthews Correlation Coefficient (MCC), Area Under the Receiver Operating Characteristic Curve (AUROC), and Area Under the Precision-Recall Curve (AUPRC), with mutation rates in the cohort. Binary classification thresholds were pre-defined at 0·5. DeLong p-values, as detailed in Materials and Methods, compare performance to the primary model (Tab. S6). Data is sorted by AUROC.

| Target | Accuracy | Precision | Sensitivity | Specificity | F1 Score | MCC | AUROC | AUPRC | Mutation Rate | (Target MUT + MSI) / Target MUT | DeLong test p-value |
|---|---|---|---|---|---|---|---|---|---|---|---|
| hypermutated | 0·78 (±0·03) | 0·66 (±0·06) | 0·79 (±0·07) | 0·78 (±0·07) | 0·71 (±0·02) | 0·55 (±0·03) | 0·86 (±0·01) | 0·76 (±0·00) | 0·34 | 0·89 | 0·8738 |
| BMPR2 | 0·74 (±0·03) | 0·41 (±0·02) | 0·84 (±0·08) | 0·72 (±0·05) | 0·55 (±0·01) | 0·45 (±0·02) | 0·84 (±0·01) | 0·45 (±0·03) | 0·19 | 0·97 | 0·5499 |
| RNF43 | 0·77 (±0·04) | 0·57 (±0·06) | 0·80 (±0·07) | 0·76 (±0·07) | 0·66 (±0·02) | 0·51 (±0·04) | 0·84 (±0·01) | 0·63 (±0·03) | 0·28 | 0·89 | 0·7227 |
| ZHX2 | 0·55 (±0·21) | 0·09 (±0·02) | 0·95 (±0·05) | 0·53 (±0·22) | 0·17 (±0·04) | 0·20 (±0·08) | 0·78 (±0·04) | 0·12 (±0·03) | 0·04 | 0·86 | 0·2049 |
| TGFBR2 | 0·67 (±0·05) | 0·17 (±0·01) | 0·75 (±0·14) | 0·66 (±0·07) | 0·27 (±0·02) | 0·24 (±0·05) | 0·78 (±0·02) | 0·20 (±0·02) | 0·08 | 0·78 | 0·3054 |
| TBX3 | 0·53 (±0·22) | 0·11 (±0·03) | 0·90 (±0·13) | 0·51 (±0·24) | 0·19 (±0·04) | 0·20 (±0·08) | 0·76 (±0·05) | 0·18 (±0·05) | 0·06 | 0·83 | 0·6648 |
| ACVR1B | 0·64 (±0·08) | 0·15 (±0·02) | 0·80 (±0·10) | 0·62 (±0·09) | 0·25 (±0·02) | 0·23 (±0·03) | 0·76 (±0·04) | 0·19 (±0·05) | 0·07 | 0·50 | 0·0459 |
| BRAF | 0·69 (±0·05) | 0·33 (±0·03) | 0·79 (±0·07) | 0·66 (±0·07) | 0·47 (±0·02) | 0·35 (±0·03) | 0·76 (±0·01) | 0·42 (±0·01) | 0·17 | 0·80 | 0·1273 |
| CCDC40 | 0·59 (±0·12) | 0·07 (±0·02) | 0·71 (±0·16) | 0·58 (±0·13) | 0·13 (±0·03) | 0·13 (±0·07) | 0·74 (±0·07) | 0·16 (±0·08) | 0·04 | 0·93 | 0·2795 |
| TP53 | 0·68 (±0·02) | 0·62 (±0·04) | 0·57 (±0·15) | 0·76 (±0·10) | 0·58 (±0·07) | 0·34 (±0·04) | 0·73 (±0·01) | 0·63 (±0·01) | 0·4 | 0·18 | 0·5195 |
| CTNND1 | 0·68 (±0·09) | 0·10 (±0·02) | 0·65 (±0·25) | 0·68 (±0·11) | 0·17 (±0·04) | 0·16 (±0·07) | 0·72 (±0·06) | 0·11 (±0·02) | 0·05 | 0·94 | 0·1893 |
| SMG1 | 0·46 (±0·18) | 0·07 (±0·02) | 0·87 (±0·16) | 0·45 (±0·19) | 0·12 (±0·03) | 0·13 (±0·06) | 0·71 (±0·11) | 0·10 (±0·04) | 0·04 | 0·77 | 0·2984 |
| CDK12 | 0·66 (±0·13) | 0·08 (±0·03) | 0·65 (±0·31) | 0·66 (±0·15) | 0·13 (±0·06) | 0·13 (±0·08) | 0·69 (±0·12) | 0·13 (±0·04) | 0·05 | 0·87 | 0·1927 |
| ZNRF3 | 0·63 (±0·06) | 0·20 (±0·02) | 0·74 (±0·06) | 0·62 (±0·07) | 0·32 (±0·02) | 0·23 (±0·03) | 0·69 (±0·02) | 0·22 (±0·02) | 0·11 | 0·84 | 0·8470 |
| ALK | 0·57 (±0·20) | 0·13 (±0·02) | 0·67 (±0·23) | 0·56 (±0·24) | 0·21 (±0·03) | 0·14 (±0·05) | 0·67 (±0·03) | 0·15 (±0·01) | 0·08 | 0·81 | 0·8384 |
| KIF1A | 0·58 (±0·11) | 0·13 (±0·02) | 0·67 (±0·13) | 0·57 (±0·13) | 0·22 (±0·02) | 0·14 (±0·03) | 0·67 (±0·01) | 0·18 (±0·03) | 0·09 | 0·71 | 0·6340 |
| SMAD2 | 0·57 (±0·11) | 0·09 (±0·01) | 0·71 (±0·17) | 0·56 (±0·12) | 0·15 (±0·02) | 0·12 (±0·05) | 0·66 (±0·05) | 0·10 (±0·02) | 0·06 | 0·44 | 0·3794 |
| KRAS | 0·52 (±0·05) | 0·37 (±0·01) | 0·79 (±0·12) | 0·41 (±0·12) | 0·50 (±0·02) | 0·20 (±0·03) | 0·65 (±0·01) | 0·41 (±0·01) | 0·3 | 0·08 | 0·0342 |
| ATM | 0·60 (±0·13) | 0·13 (±0·02) | 0·65 (±0·13) | 0·59 (±0·15) | 0·22 (±0·02) | 0·14 (±0·03) | 0·64 (±0·02) | 0·14 (±0·01) | 0·08 | 0·63 | 0·8906 |
| NRAS | 0·68 (±0·23) | 0·05 (±0·03) | 0·46 (±0·33) | 0·69 (±0·26) | 0·09 (±0·04) | 0·07 (±0·05) | 0·62 (±0·04) | 0·06 (±0·01) | 0·04 | 0·17 | 0·1990 |
| AKT1 | 0·56 (±0·16) | 0·08 (±0·01) | 0·59 (±0·23) | 0·56 (±0·19) | 0·13 (±0·02) | 0·07 (±0·04) | 0·62 (±0·03) | 0·10 (±0·02) | 0·06 | 0·53 | 0·2625 |
| DUSP16 | 0·51 (±0·18) | 0·03 (±0·01) | 0·63 (±0·11) | 0·50 (±0·19) | 0·06 (±0·02) | 0·04 (±0·06) | 0·60 (±0·11) | 0·05 (±0·02) | 0·02 | 0·71 | 0·1360 |
| APC | 0·58 (±0·02) | 0·54 (±0·02) | 0·59 (±0·16) | 0·57 (±0·12) | 0·56 (±0·08) | 0·17 (±0·06) | 0·59 (±0·03) | 0·52 (±0·03) | 0·46 | 0·21 | 0·8926 |
| PIK3CA | 0·55 (±0·13) | 0·13 (±0·03) | 0·40 (±0·21) | 0·58 (±0·19) | 0·18 (±0·06) | -0·02 (±0·03) | 0·51 (±0·04) | 0·15 (±0·02) | 0·14 | 0·46 | 0·6556 |

**Tab. S11: Performance metrics of Multi-Target Transformers excluding MSI as a target (secondary model) for external validation on TCGA.** Results are presented as mean and standard deviation across seven cross-validation folds for selected prediction targets. Metrics include Matthews Correlation Coefficient (MCC), Area Under the Receiver Operating Characteristic Curve (AUROC), and Area Under the Precision-Recall Curve (AUPRC), with mutation rates in the cohort. Binary classification thresholds were pre-defined at 0·5. DeLong p-values, as detailed in Materials and Methods, compare performance to the primary model (Tab. S7). Data is sorted by AUROC.

| **Target** | **Accuracy** | **Precision** | **Sensitivity** | **Specificity** | **F1 Score** | **MCC** | **AUROC** | **AUPRC** | **Mutation Rate** | **(Target MUT + MSI) / Target MUT** | **DeLong test p-value** |
|---|---|---|---|---|---|---|---|---|---|---|---|
| hypermutated | 0·77 (±0·07) | 0·42 (±0·10) | 0·82 (±0·10) | 0·76 (±0·10) | 0·54 (±0·05) | 0·46 (±0·05) | 0·88 (±0·01) | 0·62 (±0·04) | 0·16 | 0·85 | 0·7566 |
| FHOD3 | 0·76 (±0·07) | 0·21 (±0·04) | 0·75 (±0·13) | 0·76 (±0·09) | 0·32 (±0·03) | 0·30 (±0·02) | 0·84 (±0·02) | 0·33 (±0·05) | 0·07 | 0·65 | 0·9096 |
| BMPR2 | 0·77 (±0·06) | 0·21 (±0·04) | 0·72 (±0·09) | 0·77 (±0·07) | 0·32 (±0·04) | 0·30 (±0·03) | 0·83 (±0·01) | 0·36 (±0·04) | 0·07 | 0·84 | 0·2685 |
| WNT16 | 0·76 (±0·10) | 0·05 (±0·02) | 0·69 (±0·12) | 0·76 (±0·10) | 0·08 (±0·03) | 0·13 (±0·05) | 0·80 (±0·06) | 0·08 (±0·07) | 0·01 | 0·67 | 0·0701 |
| PLEKHA6 | 0·73 (±0·08) | 0·15 (±0·03) | 0·75 (±0·08) | 0·72 (±0·09) | 0·24 (±0·04) | 0·25 (±0·03) | 0·80 (±0·02) | 0·21 (±0·03) | 0·06 | 0·96 | 0·8039 |
| ELL2 | 0·75 (±0·08) | 0·05 (±0·01) | 0·60 (±0·26) | 0·76 (±0·09) | 0·10 (±0·03) | 0·12 (±0·06) | 0·80 (±0·01) | 0·11 (±0·04) | 0·02 | 0·40 | 0·2476 |
| MECOM | 0·75 (±0·09) | 0·15 (±0·02) | 0·71 (±0·12) | 0·75 (±0·10) | 0·24 (±0·03) | 0·24 (±0·02) | 0·79 (±0·03) | 0·17 (±0·03) | 0·05 | 0·61 | 0·4972 |
| BRAF | 0·73 (±0·07) | 0·30 (±0·05) | 0·75 (±0·07) | 0·73 (±0·08) | 0·42 (±0·05) | 0·34 (±0·05) | 0·79 (±0·02) | 0·40 (±0·03) | 0·13 | 0·64 | 0·1631 |
| RNF43 | 0·75 (±0·06) | 0·23 (±0·03) | 0·69 (±0·08) | 0·75 (±0·07) | 0·34 (±0·03) | 0·29 (±0·03) | 0·79 (±0·01) | 0·32 (±0·03) | 0·09 | 0·75 | 0·7658 |
| ZHX2 | 0·65 (±0·20) | 0·08 (±0·03) | 0·80 (±0·14) | 0·65 (±0·20) | 0·14 (±0·04) | 0·17 (±0·07) | 0·78 (±0·08) | 0·11 (±0·05) | 0·03 | 0·69 | 0·1292 |
| ZNRF3 | 0·69 (±0·10) | 0·07 (±0·02) | 0·73 (±0·08) | 0·68 (±0·10) | 0·13 (±0·03) | 0·16 (±0·03) | 0·77 (±0·02) | 0·15 (±0·06) | 0·03 | 0·85 | 0·5666 |
| RFX5 | 0·76 (±0·09) | 0·10 (±0·02) | 0·67 (±0·11) | 0·76 (±0·09) | 0·17 (±0·03) | 0·19 (±0·03) | 0·76 (±0·01) | 0·20 (±0·06) | 0·04 | 0·80 | 0·5684 |
| ALK | 0·64 (±0·15) | 0·14 (±0·05) | 0·74 (±0·17) | 0·63 (±0·17) | 0·23 (±0·05) | 0·20 (±0·04) | 0·74 (±0·03) | 0·19 (±0·04) | 0·07 | 0·50 | 0·3049 |
| TGFBR2 | 0·69 (±0·09) | 0·08 (±0·02) | 0·65 (±0·07) | 0·69 (±0·10) | 0·13 (±0·03) | 0·14 (±0·04) | 0·73 (±0·03) | 0·14 (±0·04) | 0·04 | 0·60 | 0·6144 |
| ACVR1B | 0·69 (±0·10) | 0·10 (±0·02) | 0·65 (±0·14) | 0·69 (±0·11) | 0·17 (±0·02) | 0·16 (±0·03) | 0·72 (±0·04) | 0·15 (±0·05) | 0·05 | 0·48 | 0·8886 |
| TP53 | 0·59 (±0·08) | 0·83 (±0·06) | 0·49 (±0·19) | 0·79 (±0·16) | 0·59 (±0·15) | 0·28 (±0·07) | 0·72 (±0·03) | 0·81 (±0·03) | 0·65 | 0·08 | 0·2894 |
| CDK12 | 0·67 (±0·09) | 0·12 (±0·03) | 0·63 (±0·15) | 0·68 (±0·10) | 0·20 (±0·05) | 0·16 (±0·07) | 0·70 (±0·07) | 0·16 (±0·04) | 0·06 | 0·59 | 0·1618 |
| CCDC40 | 0·66 (±0·15) | 0·07 (±0·02) | 0·61 (±0·14) | 0·66 (±0·16) | 0·13 (±0·03) | 0·11 (±0·04) | 0·67 (±0·06) | 0·13 (±0·04) | 0·04 | 0·44 | 0·8089 |
| APC | 0·66 (±0·09) | 0·84 (±0·01) | 0·70 (±0·15) | 0·52 (±0·13) | 0·76 (±0·09) | 0·20 (±0·05) | 0·67 (±0·03) | 0·87 (±0·01) | 0·79 | 0·08 | 0·7125 |
| CTNND1 | 0·68 (±0·13) | 0·08 (±0·02) | 0·54 (±0·17) | 0·69 (±0·15) | 0·14 (±0·03) | 0·11 (±0·04) | 0·66 (±0·05) | 0·12 (±0·05) | 0·05 | 0·40 | 0·0187 |
| CHD1 | 0·72 (±0·08) | 0·09 (±0·01) | 0·51 (±0·17) | 0·73 (±0·09) | 0·16 (±0·02) | 0·12 (±0·04) | 0·66 (±0·01) | 0·10 (±0·03) | 0·05 | 0·55 | 0·7945 |
| TRPS1 | 0·75 (±0·10) | 0·20 (±0·04) | 0·39 (±0·18) | 0·79 (±0·13) | 0·25 (±0·03) | 0·15 (±0·04) | 0·65 (±0·04) | 0·22 (±0·02) | 0·11 | 0·36 | 0·2477 |
| AKT1 | 0·65 (±0·18) | 0·05 (±0·02) | 0·63 (±0·12) | 0·65 (±0·19) | 0·08 (±0·04) | 0·10 (±0·05) | 0·64 (±0·08) | 0·08 (±0·08) | 0·02 | 0·67 | 0·0020 |
| ATM | 0·63 | 0·22 | 0·58 | 0·64 | 0·30 | 0·17 | 0·64 | 0·24 | 0·13 | 0·37 | 0·2454 |

| | | | | | | | | | | |
|---|---|---|---|---|---|---|---|---|---|---|
| | (±0·16) | (±0·06) | (±0·18) | (±0·22) | (±0·03) | (±0·05) | (±0·02) | (±0·02) | | | |
| SMAD2 | 0·60 (±0·11) | 0·07 (±0·02) | 0·57 (±0·25) | 0·60 (±0·13) | 0·13 (±0·03) | 0·08 (±0·06) | 0·62 (±0·07) | 0·08 (±0·02) | 0·05 | 0·09 | 0·2060 |
| KIF1A | 0·64 (±0·08) | 0·08 (±0·01) | 0·48 (±0·09) | 0·65 (±0·09) | 0·14 (±0·01) | 0·06 (±0·02) | 0·62 (±0·02) | 0·11 (±0·02) | 0·06 | 0·41 | 0·6627 |
| DUSP16 | 0·53 (±0·20) | 0·00 (±0·00) | 0·71 (±0·49) | 0·53 (±0·20) | 0·01 (±0·01) | 0·02 (±0·06) | 0·61 (±0·31) | 0·01 (±0·01) | 0.00 | 1.00 | |
| NRAS | 0·65 (±0·19) | 0·09 (±0·03) | 0·43 (±0·33) | 0·67 (±0·22) | 0·13 (±0·06) | 0·05 (±0·08) | 0·61 (±0·07) | 0·11 (±0·03) | 0·07 | 0·10 | 0·3766 |
| SMG1 | 0·47 (±0·24) | 0·06 (±0·02) | 0·73 (±0·15) | 0·46 (±0·25) | 0·11 (±0·03) | 0·08 (±0·05) | 0·60 (±0·07) | 0·08 (±0·03) | 0·04 | 0·50 | 0·8932 |
| TBX3 | 0·60 (±0·23) | 0·07 (±0·02) | 0·59 (±0·20) | 0·60 (±0·24) | 0·12 (±0·03) | 0·08 (±0·05) | 0·59 (±0·04) | 0·09 (±0·05) | 0·04 | 0·42 | 0·5554 |
| PIK3CA | 0·54 (±0·09) | 0·33 (±0·02) | 0·59 (±0·24) | 0·52 (±0·22) | 0·40 (±0·07) | 0·11 (±0·03) | 0·58 (±0·03) | 0·34 (±0·02) | 0·28 | 0·20 | 0·7623 |
| KRAS | 0·50 (±0·02) | 0·46 (±0·01) | 0·72 (±0·10) | 0·32 (±0·11) | 0·56 (±0·03) | 0·04 (±0·02) | 0·55 (±0·02) | 0·50 (±0·03) | 0·44 | 0·10 | 0·1642 |

**Tab. S12: Performance metrics of Multi-Target Transformers excluding MSI as a target (secondary model) for external validation on CPTAC.** Results are presented as mean and standard deviation across seven cross-validation folds for selected prediction targets. Metrics include Matthews Correlation Coefficient (MCC), Area Under the Receiver Operating Characteristic Curve (AUROC), and Area Under the Precision-Recall Curve (AUPRC), with mutation rates in the cohort. Binary classification thresholds were pre-defined at 0·5. DeLong p-values, as detailed in Materials and Methods, compare performance to the primary model (Tab. S8). Data is sorted by AUROC.

| Target | Accuracy | Precision | Sensitivity | Specificity | F1 Score | MCC | AUROC | AUPRC | Mutation Rate | (Target MUT + MSI) / Target MUT | DeLong test p-value |
|---|---|---|---|---|---|---|---|---|---|---|---|
| CCDC40 | 0·36 (±0·17) | 0·10 (±0·03) | 0·96 (±0·07) | 0·32 (±0·19) | 0·17 (±0·05) | 0·15 (±0·09) | 0·81 (±0·06) | 0·29 (±0·08) | 0·07 | 1.00 | 0·3484 |
| TGFBR2 | 0·28 (±0·13) | 0·07 (±0·01) | 0·98 (±0·06) | 0·24 (±0·14) | 0·14 (±0·02) | 0·12 (±0·07) | 0·80 (±0·04) | 0·34 (±0·14) | 0·06 | 1.00 | 0·7400 |
| RNF43 | 0·38 (±0·13) | 0·19 (±0·03) | 0·93 (±0·07) | 0·28 (±0·17) | 0·32 (±0·04) | 0·17 (±0·08) | 0·80 (±0·03) | 0·49 (±0·01) | 0·15 | 0·88 | 0·6513 |
| BRAF | 0·39 (±0·15) | 0·19 (±0·03) | 0·91 (±0·06) | 0·30 (±0·19) | 0·32 (±0·04) | 0·17 (±0·08) | 0·80 (±0·03) | 0·55 (±0·06) | 0·15 | 0·81 | 0·5381 |
| FHOD3 | 0·44 (±0·23) | 0·18 (±0·07) | 0·93 (±0·09) | 0·37 (±0·27) | 0·30 (±0·09) | 0·22 (±0·12) | 0·79 (±0·04) | 0·31 (±0·04) | 0·11 | 0·92 | 0·0786 |
| RFX5 | 0·43 (±0·26) | 0·08 (±0·03) | 0·89 (±0·16) | 0·41 (±0·28) | 0·14 (±0·05) | 0·14 (±0·07) | 0·77 (±0·06) | 0·16 (±0·05) | 0·05 | 1.00 | 0·7903 |
| BMPR2 | 0·43 (±0·14) | 0·18 (±0·04) | 0·95 (±0·09) | 0·35 (±0·17) | 0·30 (±0·05) | 0·22 (±0·07) | 0·77 (±0·03) | 0·29 (±0·02) | 0·12 | 0·85 | 0·3481 |
| PLEKHA6 | 0·39 (±0·11) | 0·10 (±0·02) | 0·89 (±0·13) | 0·35 (±0·13) | 0·18 (±0·03) | 0·14 (±0·08) | 0·75 (±0·08) | 0·21 (±0·05) | 0·08 | 0·88 | 0·0501 |
| ACVR1B | 0·25 (±0·18) | 0·04 (±0·01) | 0·95 (±0·13) | 0·23 (±0·18) | 0·07 (±0·02) | 0·07 (±0·06) | 0·75 (±0·05) | 0·18 (±0·11) | 0·03 | 0·67 | 0·5133 |
| KIF1A | 0·25 (±0·20) | 0·09 (±0·03) | 0·96 (±0·09) | 0·19 (±0·22) | 0·17 (±0·04) | 0·11 (±0·07) | 0·74 (±0·04) | 0·22 (±0·03) | 0·08 | 0·88 | 0·3716 |
| WNT16 | 0·42 (±0·18) | 0·14 (±0·05) | 0·89 (±0·09) | 0·37 (±0·21) | 0·24 (±0·06) | 0·17 (±0·09) | 0·73 (±0·03) | 0·22 (±0·03) | 0·09 | 0·80 | 0·9503 |
| TBX3 | 0·23 (±0·10) | 0·09 (±0·01) | 1·00 (±0·00) | 0·17 (±0·11) | 0·17 (±0·02) | 0·12 (±0·05) | 0·72 (±0·08) | 0·21 (±0·11) | 0·08 | 0·62 | 0·1304 |
| ZNRF3 | 0·31 (±0·15) | 0·10 (±0·02) | 0·84 (±0·13) | 0·26 (±0·17) | 0·18 (±0·03) | 0·07 (±0·07) | 0·72 (±0·03) | 0·31 (±0·02) | 0·08 | 0·78 | 0·2917 |
| KRAS | 0·67 (±0·08) | 0·55 (±0·11) | 0·49 (±0·25) | 0·75 (±0·24) | 0·47 (±0·09) | 0·27 (±0·07) | 0·72 (±0·03) | 0·58 (±0·06) | 0·33 | 0.00 | 0·3861 |
| MECOM | 0·36 (±0·17) | 0·10 (±0·03) | 0·88 (±0·10) | 0·31 (±0·19) | 0·18 (±0·04) | 0·11 (±0·08) | 0·71 (±0·03) | 0·19 (±0·04) | 0·08 | 0·88 | 0·8027 |
| ALK | 0·37 (±0·28) | 0·10 (±0·03) | 0·84 (±0·33) | 0·33 (±0·33) | 0·17 (±0·03) | 0·12 (±0·06) | 0·70 (±0·06) | 0·25 (±0·10) | 0·08 | 0·75 | 0·7380 |
| CHD1 | 0·40 (±0·22) | 0·08 (±0·03) | 0·76 (±0·30) | 0·38 (±0·25) | 0·13 (±0·02) | 0·08 (±0·06) | 0·69 (±0·07) | 0·15 (±0·03) | 0·06 | 0·83 | 0·8215 |
| TRPS1 | 0·55 (±0·17) | 0·23 (±0·05) | 0·60 (±0·23) | 0·54 (±0·25) | 0·31 (±0·03) | 0·12 (±0·05) | 0·66 (±0·03) | 0·29 (±0·03) | 0·17 | 0·56 | 0·9008 |
| AKT1 | 0·19 (±0·18) | 0·01 (±0·00) | 1·00 (±0·00) | 0·19 (±0·18) | 0·02 (±0·01) | 0·04 (±0·03) | 0·65 (±0·27) | 0·07 (±0·08) | 0·01 | 1.00 | - |
| PIK3CA | 0·57 (±0·15) | 0·23 (±0·11) | 0·54 (±0·34) | 0·58 (±0·27) | 0·31 (±0·15) | 0·10 (±0·12) | 0·65 (±0·05) | 0·37 (±0·08) | 0·22 | 0·43 | 0·7644 |
| APC | 0·43 (±0·18) | 0·78 (±0·07) | 0·33 (±0·34) | 0·74 (±0·31) | 0·39 (±0·29) | 0·07 (±0·10) | 0·64 (±0·07) | 0·82 (±0·04) | 0·75 | 0·14 | 0·7638 |
| CDK12 | 0·39 (±0·15) | 0·03 (±0·01) | 0·71 (±0·36) | 0·38 (±0·16) | 0·06 (±0·03) | 0·03 (±0·11) | 0·63 (±0·13) | 0·16 (±0·10) | 0·03 | 0·67 | 0·3362 |
| SMAD2 | 0·29 (±0·13) | 0·11 (±0·02) | 0·91 (±0·19) | 0·23 (±0·16) | 0·20 (±0·03) | 0·11 (±0·08) | 0·63 (±0·09) | 0·15 (±0·03) | 0·09 | 0·40 | 0·0349 |
| DUSP16 | 0·36 | 0·06 | 0·81 | 0·33 | 0·11 | 0·08 | 0·62 | 0·10 | 0·06 | 0·33 | 0·0248 |

| | | | | | | | | | | |
|---|---|---|---|---|---|---|---|---|---|---|
| | (±0·28) | (±0·03) | (±0·38) | (±0·32) | (±0·05) | (±0·07) | (±0·16) | (±0·03) | | | |
| TP53 | 0·52 (±0·05) | 0·57 (±0·27) | 0·20 (±0·18) | 0·89 (±0·10) | 0·27 (±0·22) | 0·09 (±0·13) | 0·61 (±0·02) | 0·64 (±0·04) | 0·53 | 0·14 | 0·3280 |
| ATM | 0·36 (±0·21) | 0·09 (±0·02) | 0·79 (±0·25) | 0·32 (±0·25) | 0·16 (±0·03) | 0·08 (±0·06) | 0·59 (±0·09) | 0·13 (±0·03) | 0·08 | 0·50 | 0·8098 |
| SMG1 | 0·21 (±0·18) | 0·08 (±0·02) | 0·94 (±0·08) | 0·16 (±0·19) | 0·14 (±0·03) | 0·02 (±0·18) | 0·58 (±0·18) | 0·13 (±0·08) | 0·07 | 0·71 | 0·2335 |
| ZHX2 | 0·26 (±0·19) | 0·04 (±0·01) | 0·75 (±0·20) | 0·24 (±0·20) | 0·07 (±0·01) | -0·00 (±0·06) | 0·51 (±0·08) | 0·08 (±0·03) | 0·04 | 0·50 | 0·6480 |
| NRAS | 0·77 (±0·28) | 0·02 (±0·02) | 0·14 (±0·31) | 0·80 (±0·32) | 0·02 (±0·04) | -0·04 (±0·03) | 0·48 (±0·13) | 0·07 (±0·02) | 0·06 | 0·17 | 0·3146 |
| ELL2 | 0·46 (±0·32) | 0·03 (±0·03) | 0·50 (±0·38) | 0·46 (±0·34) | 0·06 (±0·05) | -0·03 (±0·12) | 0·45 (±0·08) | 0·05 (±0·01) | 0·04 | 0·50 | 0·5055 |
| CTNND1 | 0·54 (±0·30) | 0·01 (±0·01) | 0·36 (±0·38) | 0·54 (±0·31) | 0·02 (±0·02) | -0·03 (±0·04) | 0·34 (±0·11) | 0·02 (±0·00) | 0·02 | 0.00 | 0·7825 |

**Tab. S13: Association Rule Mining Results.** Comprehensive overview of the relationships between various genetic alterations. Each relationship is characterized by an initiating genetic alteration, the 'Antecedent', and a resulting genetic alteration, the 'Consequent'. The relationships are evaluated based on several metrics. 'Support' refers to the frequency of occurrence of the relationship. 'Confidence' measures the predictive power of the relationship. 'Lift' assesses the degree of association between the Antecedent and Consequent. 'Leverage' calculates the difference between observed and expected Support. 'Conviction' gauges the dependability of the relationship. 'Zhang's Metric' is a measure of the relationship's interestingness. The relationships are sorted in descending order of Confidence. The selection encompasses a variety of relevant and representative prediction targets from all prediction targets investigated in this study.

| Antecedent | Consequent | Antecedent support | Consequent support | Support | Confidence | Lift | Leverage | Conviction | Zhang's metric |
|---|---|---|---|---|---|---|---|---|---|
| BMPR2 | MSI | 0·15 | 0·25 | 0·15 | 0·95 | 3·88 | 0·11 | 16·07 | 0·88 |
| WNT16 | hypermutated | 0·03 | 0·26 | 0·03 | 0·95 | 3·69 | 0·02 | 15·58 | 0·75 |
| WNT16 | MSI | 0·03 | 0·25 | 0·03 | 0·95 | 3·88 | 0·02 | 15·84 | 0·77 |
| MSI | hypermutated | 0·25 | 0·26 | 0·23 | 0·94 | 3·66 | 0·17 | 13·4 | 0·96 |
| BMPR2 | hypermutated | 0·15 | 0·26 | 0·14 | 0·94 | 3·64 | 0·1 | 12·16 | 0·86 |
| NRAS | MSS | 0·04 | 0·75 | 0·04 | 0·93 | 1·24 | 0·01 | 3·75 | 0·2 |
| RFX5 | MSI | 0·03 | 0·25 | 0·03 | 0·93 | 3·8 | 0·02 | 11·31 | 0·76 |
| KRAS | MSS | 0·33 | 0·75 | 0·3 | 0·93 | 1·23 | 0·06 | 3·32 | 0·28 |
| ZNRF3 | hypermutated | 0·11 | 0·26 | 0·1 | 0·92 | 3·58 | 0·07 | 9·52 | 0·81 |
| ELL2 | hypermutated | 0·02 | 0·26 | 0·02 | 0·92 | 3·55 | 0·01 | 8·91 | 0·73 |
| CDK12 | hypermutated | 0·04 | 0·26 | 0·03 | 0·91 | 3·51 | 0·02 | 7·87 | 0·74 |
| hypermutated | MSI | 0·26 | 0·25 | 0·23 | 0·9 | 3·66 | 0·17 | 7·54 | 0·98 |
| MECOM | hypermutated | 0·06 | 0·26 | 0·05 | 0·9 | 3·49 | 0·04 | 7·42 | 0·76 |
| PLEKHA6 | MSI | 0·06 | 0·25 | 0·06 | 0·9 | 3·65 | 0·04 | 7·38 | 0·78 |
| CCDC40 | hypermutated | 0·04 | 0·26 | 0·04 | 0·89 | 3·45 | 0·02 | 6·8 | 0·74 |
| ZNRF3 | MSI | 0·11 | 0·25 | 0·1 | 0·89 | 3·62 | 0·07 | 6·83 | 0·81 |
| RFX5 | hypermutated | 0·03 | 0·26 | 0·03 | 0·89 | 3·45 | 0·02 | 6·68 | 0·73 |
| FHOD3 | hypermutated | 0·08 | 0·26 | 0·07 | 0·89 | 3·44 | 0·05 | 6·56 | 0·77 |
| PLEKHA6 | hypermutated | 0·06 | 0·26 | 0·06 | 0·89 | 3·44 | 0·04 | 6·53 | 0·76 |
| FHOD3 | MSI | 0·08 | 0·25 | 0·07 | 0·88 | 3·57 | 0·05 | 6·2 | 0·78 |
| CTNND1 | hypermutated | 0·04 | 0·26 | 0·04 | 0·88 | 3·4 | 0·03 | 6·04 | 0·74 |
| MECOM | MSI | 0·06 | 0·25 | 0·05 | 0·87 | 3·56 | 0·04 | 6·03 | 0·76 |
| TP53 | MSS | 0·58 | 0·75 | 0·5 | 0·87 | 1·16 | 0·07 | 1·96 | 0·32 |
| CHD1 | hypermutated | 0·04 | 0·26 | 0·04 | 0·87 | 3·36 | 0·03 | 5·57 | 0·73 |
| APC | MSS | 0·59 | 0·75 | 0·51 | 0·86 | 1·14 | 0·06 | 1·76 | 0·3 |
| CTNND1 | MSI | 0·04 | 0·25 | 0·04 | 0·86 | 3·5 | 0·03 | 5·37 | 0·74 |
| RNF43 | MSI | 0·21 | 0·25 | 0·17 | 0·84 | 3·41 | 0·12 | 4·67 | 0·89 |
| CHD1 | MSI | 0·04 | 0·25 | 0·04 | 0·83 | 3·39 | 0·03 | 4·53 | 0·74 |
| ELL2 | MSI | 0·02 | 0·25 | 0·01 | 0·83 | 3·39 | 0·01 | 4·53 | 0·72 |
| CDK12 | MSI | 0·04 | 0·25 | 0·03 | 0·81 | 3·3 | 0·02 | 4 | 0·72 |
| RNF43 | hypermutated | 0·21 | 0·26 | 0·17 | 0·81 | 3·13 | 0·11 | 3·86 | 0·86 |
| ZHX2 | hypermutated | 0·03 | 0·26 | 0·03 | 0·8 | 3·12 | 0·02 | 3·79 | 0·7 |
| RFX5 | BRAF | 0·03 | 0·23 | 0·03 | 0·8 | 3·49 | 0·02 | 3·85 | 0·74 |
| AKT1 | hypermutated | 0·04 | 0·26 | 0·03 | 0·79 | 3·07 | 0·02 | 3·58 | 0·7 |
| ELL2 | BRAF | 0·02 | 0·23 | 0·01 | 0·79 | 3·45 | 0·01 | 3·7 | 0·72 |
| TGFBR2 | hypermutated | 0·06 | 0·26 | 0·05 | 0·78 | 3·04 | 0·03 | 3·42 | 0·71 |
| CCDC40 | MSI | 0·04 | 0·25 | 0·03 | 0·78 | 3·18 | 0·02 | 3·46 | 0·71 |
| SMG1 | hypermutated | 0·04 | 0·26 | 0·03 | 0·78 | 3·02 | 0·02 | 3·37 | 0·7 |
| MECOM | BRAF | 0·06 | 0·23 | 0·04 | 0·78 | 3·38 | 0·03 | 3·43 | 0·75 |
| TBX3 | hypermutated | 0·05 | 0·26 | 0·04 | 0·77 | 2·98 | 0·03 | 3·2 | 0·7 |
| ZNRF3 | RNF43 | 0·11 | 0·21 | 0·08 | 0·77 | 3·68 | 0·06 | 3·39 | 0·82 |
| CCDC40 | RNF43 | 0·04 | 0·21 | 0·03 | 0·76 | 3·66 | 0·02 | 3·35 | 0·76 |
| NRAS | APC | 0·04 | 0·59 | 0·03 | 0·75 | 1·28 | 0·01 | 1·67 | 0·23 |
| ALK | hypermutated | 0·08 | 0·26 | 0·06 | 0·75 | 2·9 | 0·04 | 2·94 | 0·71 |
| TGFBR2 | MSI | 0·06 | 0·25 | 0·04 | 0·75 | 3·04 | 0·03 | 2·98 | 0·71 |
| FHOD3 | BRAF | 0·08 | 0·23 | 0·06 | 0·74 | 3·22 | 0·04 | 2·95 | 0·75 |
| PLEKHA6 | BRAF | 0·06 | 0·23 | 0·05 | 0·74 | 3·22 | 0·03 | 2·95 | 0·74 |
| CDK12 | RNF43 | 0·04 | 0·21 | 0·03 | 0·74 | 3·53 | 0·02 | 3 | 0·75 |
| CHD1 | RNF43 | 0·04 | 0·21 | 0·03 | 0·73 | 3·52 | 0·02 | 2·97 | 0·75 |
| CHD1 | BRAF | 0·04 | 0·23 | 0·03 | 0·73 | 3·2 | 0·02 | 2·89 | 0·72 |
| KRAS | APC | 0·33 | 0·59 | 0·24 | 0·73 | 1·24 | 0·05 | 1·52 | 0·29 |
| KIF1A | hypermutated | 0·1 | 0·26 | 0·08 | 0·73 | 2·82 | 0·05 | 2·71 | 0·72 |
| FHOD3 | RNF43 | 0·08 | 0·21 | 0·06 | 0·72 | 3·46 | 0·04 | 2·84 | 0·78 |
| WNT16 | BRAF | 0·03 | 0·23 | 0·02 | 0·71 | 3·12 | 0·01 | 2·7 | 0·7 |
| WNT16 | RNF43 | 0·03 | 0·21 | 0·02 | 0·71 | 3·43 | 0·02 | 2·77 | 0·73 |
| MSI | RNF43 | 0·25 | 0·21 | 0·17 | 0·71 | 3·41 | 0·12 | 2·74 | 0·94 |
| BMPR2 | RNF43 | 0·15 | 0·21 | 0·11 | 0·7 | 3·38 | 0·08 | 2·68 | 0·83 |
| MECOM | RNF43 | 0·06 | 0·21 | 0·04 | 0·7 | 3·36 | 0·03 | 2·64 | 0·74 |
| ZHX2 | MSI | 0·03 | 0·25 | 0·02 | 0·7 | 2·83 | 0·01 | 2·48 | 0·67 |
| TBX3 | MSI | 0·05 | 0·25 | 0·03 | 0·7 | 2·83 | 0·02 | 2·48 | 0·68 |
| SMG1 | MSI | 0·04 | 0·25 | 0·03 | 0·69 | 2·83 | 0·02 | 2·47 | 0·67 |
| ACVR1B | hypermutated | 0·05 | 0·26 | 0·04 | 0·69 | 2·69 | 0·02 | 2·42 | 0·66 |
| RFX5 | RNF43 | 0·03 | 0·21 | 0·02 | 0·69 | 3·3 | 0·02 | 2·54 | 0·72 |
| BRAF | hypermutated | 0·23 | 0·26 | 0·16 | 0·68 | 2·65 | 0·1 | 2·35 | 0·81 |
| BRAF | MSI | 0·23 | 0·25 | 0·16 | 0·68 | 2·76 | 0·1 | 2·34 | 0·83 |
| DUSP16 | hypermutated | 0·03 | 0·26 | 0·02 | 0·68 | 2·62 | 0·01 | 2·29 | 0·64 |

| | | | | | | | | | |
|---|---|---|---|---|---|---|---|---|---|
| ALK | MSI | 0·08 | 0·25 | 0·05 | 0·67 | 2·74 | 0·03 | 2·31 | 0·69 |
| CCDC40 | BRAF | 0·04 | 0·23 | 0·03 | 0·67 | 2·93 | 0·02 | 2·36 | 0·69 |
| MSS | APC | 0·75 | 0·59 | 0·51 | 0·67 | 1·14 | 0·06 | 1·25 | 0·5 |
| RNF43 | BRAF | 0·21 | 0·23 | 0·14 | 0·67 | 2·92 | 0·09 | 2·34 | 0·83 |
| MSS | TP53 | 0·75 | 0·58 | 0·5 | 0·67 | 1·16 | 0·07 | 1·28 | 0·56 |
| TRPS1 | hypermutated | 0·06 | 0·26 | 0·04 | 0·67 | 2·59 | 0·02 | 2·23 | 0·65 |
| ELL2 | RNF43 | 0·02 | 0·21 | 0·01 | 0·67 | 3·2 | 0·01 | 2·37 | 0·7 |
| NRAS | TP53 | 0·04 | 0·58 | 0·03 | 0·66 | 1·14 | 0 | 1·23 | 0·13 |
| hypermutated | RNF43 | 0·26 | 0·21 | 0·17 | 0·65 | 3·13 | 0·11 | 2·28 | 0·92 |
| TGFBR2 | BRAF | 0·06 | 0·23 | 0·04 | 0·65 | 2·84 | 0·03 | 2·21 | 0·69 |
| DUSP16 | BRAF | 0·03 | 0·23 | 0·02 | 0·65 | 2·83 | 0·01 | 2·19 | 0·66 |
| DUSP16 | MSI | 0·03 | 0·25 | 0·02 | 0·65 | 2·64 | 0·01 | 2·15 | 0·64 |
| TP53 | APC | 0·58 | 0·59 | 0·37 | 0·65 | 1·1 | 0·03 | 1·17 | 0·22 |
| WNT16 | BMPR2 | 0·03 | 0·15 | 0·02 | 0·64 | 4·21 | 0·01 | 2·37 | 0·79 |
| BMPR2 | BRAF | 0·15 | 0·23 | 0·1 | 0·64 | 2·79 | 0·06 | 2·13 | 0·76 |
| PLEKHA6 | RNF43 | 0·06 | 0·21 | 0·04 | 0·64 | 3·05 | 0·03 | 2·18 | 0·72 |
| APC | TP53 | 0·59 | 0·58 | 0·37 | 0·64 | 1·1 | 0·03 | 1·16 | 0·22 |
| MSI | BRAF | 0·25 | 0·23 | 0·16 | 0·63 | 2·76 | 0·1 | 2·1 | 0·85 |
| CTNND1 | BRAF | 0·04 | 0·23 | 0·03 | 0·63 | 2·76 | 0·02 | 2·09 | 0·66 |
| SMAD2 | APC | 0·04 | 0·59 | 0·03 | 0·63 | 1·07 | 0 | 1·11 | 0·07 |
| TRPS1 | MSI | 0·06 | 0·25 | 0·04 | 0·63 | 2·56 | 0·02 | 2·03 | 0·64 |
| ZNRF3 | BRAF | 0·11 | 0·23 | 0·07 | 0·62 | 2·72 | 0·04 | 2·05 | 0·71 |
| CDK12 | BRAF | 0·04 | 0·23 | 0·02 | 0·62 | 2·72 | 0·01 | 2·04 | 0·66 |
| CHD1 | BMPR2 | 0·04 | 0·15 | 0·03 | 0·62 | 4·04 | 0·02 | 2·21 | 0·79 |
| KIF1A | MSI | 0·1 | 0·25 | 0·06 | 0·62 | 2·51 | 0·04 | 1·97 | 0·67 |
| PIK3CA | APC | 0·18 | 0·59 | 0·11 | 0·62 | 1·05 | 0·01 | 1·07 | 0·05 |
| ATM | hypermutated | 0·11 | 0·26 | 0·07 | 0·61 | 2·38 | 0·04 | 1·92 | 0·65 |
| ACVR1B | MSI | 0·05 | 0·25 | 0·03 | 0·61 | 2·5 | 0·02 | 1·95 | 0·63 |
| BRAF | RNF43 | 0·23 | 0·21 | 0·14 | 0·61 | 2·92 | 0·09 | 2·03 | 0·85 |
| ZHX2 | RNF43 | 0·03 | 0·21 | 0·02 | 0·61 | 2·92 | 0·01 | 2·02 | 0·68 |
| hypermutated | BRAF | 0·26 | 0·23 | 0·16 | 0·61 | 2·65 | 0·1 | 1·97 | 0·84 |
| ZNRF3 | BMPR2 | 0·11 | 0·15 | 0·07 | 0·6 | 3·96 | 0·05 | 2·14 | 0·84 |
| AKT1 | MSI | 0·04 | 0·25 | 0·02 | 0·6 | 2·46 | 0·01 | 1·9 | 0·62 |
| RFX5 | BMPR2 | 0·03 | 0·15 | 0·02 | 0·6 | 3·93 | 0·01 | 2·12 | 0·77 |
| CTNND1 | RNF43 | 0·04 | 0·21 | 0·02 | 0·6 | 2·86 | 0·02 | 1·96 | 0·68 |
| DUSP16 | APC | 0·03 | 0·59 | 0·02 | 0·59 | 1·01 | 0 | 1·01 | 0·01 |
| DUSP16 | RNF43 | 0·03 | 0·21 | 0·02 | 0·59 | 2·85 | 0·01 | 1·95 | 0·67 |
| SMG1 | BRAF | 0·04 | 0·23 | 0·03 | 0·59 | 2·59 | 0·02 | 1·89 | 0·64 |
| MSI | BMPR2 | 0·25 | 0·15 | 0·15 | 0·59 | 3·88 | 0·11 | 2·08 | 0·98 |
| PIK3CA | MSS | 0·18 | 0·75 | 0·11 | 0·58 | 0·77 | -0·03 | 0·58 | -0·27 |
| TRPS1 | BRAF | 0·06 | 0·23 | 0·03 | 0·58 | 2·52 | 0·02 | 1·82 | 0·64 |
| SMG1 | RNF43 | 0·04 | 0·21 | 0·02 | 0·58 | 2·76 | 0·02 | 1·87 | 0·67 |
| ATM | APC | 0·11 | 0·59 | 0·06 | 0·58 | 0·98 | 0 | 0·97 | -0·03 |
| ATM | MSI | 0·11 | 0·25 | 0·06 | 0·57 | 2·31 | 0·04 | 1·75 | 0·64 |
| PLEKHA6 | BMPR2 | 0·06 | 0·15 | 0·04 | 0·57 | 3·72 | 0·03 | 1·96 | 0·78 |
| ZHX2 | BRAF | 0·03 | 0·23 | 0·02 | 0·57 | 2·47 | 0·01 | 1·77 | 0·61 |
| SMAD2 | MSS | 0·04 | 0·75 | 0·03 | 0·56 | 0·75 | -0·01 | 0·56 | -0·26 |
| ALK | RNF43 | 0·08 | 0·21 | 0·04 | 0·56 | 2·69 | 0·03 | 1·8 | 0·68 |
| KRAS | TP53 | 0·33 | 0·58 | 0·18 | 0·56 | 0·96 | -0·01 | 0·95 | -0·05 |
| hypermutated | BMPR2 | 0·26 | 0·15 | 0·14 | 0·56 | 3·64 | 0·1 | 1·91 | 0·98 |
| TGFBR2 | RNF43 | 0·06 | 0·21 | 0·03 | 0·55 | 2·66 | 0·02 | 1·78 | 0·66 |
| TBX3 | RNF43 | 0·05 | 0·21 | 0·03 | 0·55 | 2·64 | 0·02 | 1·76 | 0·65 |
| MECOM | BMPR2 | 0·06 | 0·15 | 0·03 | 0·55 | 3·6 | 0·02 | 1·88 | 0·77 |
| SMAD2 | hypermutated | 0·04 | 0·26 | 0·02 | 0·55 | 2·13 | 0·01 | 1·64 | 0·55 |
| CCDC40 | APC | 0·04 | 0·59 | 0·02 | 0·55 | 0·93 | 0 | 0·9 | -0·08 |
| FHOD3 | BMPR2 | 0·08 | 0·15 | 0·04 | 0·54 | 3·53 | 0·03 | 1·84 | 0·78 |
| MECOM | ZNRF3 | 0·06 | 0·11 | 0·03 | 0·54 | 4·87 | 0·02 | 1·92 | 0·84 |
| ACVR1B | APC | 0·05 | 0·59 | 0·03 | 0·53 | 0·91 | 0 | 0·88 | -0·1 |
| RFX5 | ZNRF3 | 0·03 | 0·11 | 0·02 | 0·53 | 4·83 | 0·01 | 1·91 | 0·82 |
| RFX5 | FHOD3 | 0·03 | 0·08 | 0·02 | 0·53 | 6·47 | 0·01 | 1·97 | 0·87 |
| AKT1 | APC | 0·04 | 0·59 | 0·02 | 0·53 | 0·9 | 0 | 0·87 | -0·11 |
| CTNND1 | BMPR2 | 0·04 | 0·15 | 0·02 | 0·53 | 3·45 | 0·02 | 1·79 | 0·74 |
| SMG1 | APC | 0·04 | 0·59 | 0·02 | 0·53 | 0·89 | 0 | 0·87 | -0·11 |
| WNT16 | FHOD3 | 0·03 | 0·08 | 0·02 | 0·52 | 6·36 | 0·01 | 1·93 | 0·87 |
| CHD1 | FHOD3 | 0·04 | 0·08 | 0·02 | 0·52 | 6·27 | 0·02 | 1·9 | 0·88 |
| RNF43 | BMPR2 | 0·21 | 0·15 | 0·11 | 0·52 | 3·38 | 0·08 | 1·75 | 0·89 |
| CDK12 | TP53 | 0·04 | 0·58 | 0·02 | 0·51 | 0·88 | 0 | 0·86 | -0·12 |
| CCDC40 | BMPR2 | 0·04 | 0·15 | 0·02 | 0·51 | 3·34 | 0·01 | 1·73 | 0·73 |
| TBX3 | BMPR2 | 0·05 | 0·15 | 0·03 | 0·51 | 3·32 | 0·02 | 1·72 | 0·74 |
| KIF1A | RNF43 | 0·1 | 0·21 | 0·05 | 0·51 | 2·43 | 0·03 | 1·61 | 0·66 |
| TRPS1 | RNF43 | 0·06 | 0·21 | 0·03 | 0·5 | 2·4 | 0·02 | 1·58 | 0·62 |
| MECOM | FHOD3 | 0·06 | 0·08 | 0·03 | 0·5 | 6·07 | 0·02 | 1·84 | 0·89 |
| CHD1 | ZNRF3 | 0·04 | 0·11 | 0·02 | 0·5 | 4·53 | 0·02 | 1·78 | 0·81 |
| ZHX2 | APC | 0·03 | 0·59 | 0·02 | 0·5 | 0·85 | 0 | 0·82 | -0·16 |

**Tab. S14: Performance metrics of Multi-Target Transformers for external validation on the GECCO test cohort.** Mean and standard deviation from the 7 folds of the cross-validation for relevant selected prediction targets. The threshold for binary classification is pre-defined as 0·5. The evaluation metrics include the Matthews Correlation Coefficient (MCC), the Area Under the Receiver Operating Characteristic Curve (AUROC), and the Area Under the Precision-Recall Curve (AUPRC), along with the corresponding mutation rates in external cohorts. Mutation rate contains the ratio of mutations of the respective target within the external data set. The data is sorted for AUROC, as shown in Fig. S3B.

| **Target** | **Accuracy** | **Precision** | **Sensitivity** | **Specificity** | **F1 Score** | **MCC** | **AUROC** | **AUPRC** | **Mutation Rate** | **(Target MUT + MSI) / Target MUT** |
|---|---|---|---|---|---|---|---|---|---|---|
| MSI | 0·85 (±0·04) | 0·68 (±0·09) | 0·88 (±0·05) | 0·83 (±0·07) | 0·76 (±0·04) | 0·67 (±0·06) | 0·93 (±0·01) | 0·86 (±0·03) | 0·27 | 1·00 |
| PLEKHA6 | 0·77 (±0·05) | 0·28 (±0·04) | 0·91 (±0·08) | 0·76 (±0·06) | 0·42 (±0·04) | 0·42 (±0·03) | 0·90 (±0·01) | 0·38 (±0·06) | 0·04 | 0·97 |
| hypermutated | 0·78 (±0·05) | 0·58 (±0·07) | 0·85 (±0·04) | 0·76 (±0·08) | 0·69 (±0·04) | 0·56 (±0·05) | 0·88 (±0·01) | 0·74 (±0·02) | 0·28 | 0·92 |
| CHD1 | 0·76 (±0·07) | 0·12 (±0·03) | 0·84 (±0·15) | 0·76 (±0·08) | 0·2 (±0·03) | 0·26 (±0·02) | 0·87 (±0·02) | 0·17 (±0·06) | 0·02 | 0·91 |
| BMPR2 | 0·75 (±0·05) | 0·4 (±0·05) | 0·89 (±0·08) | 0·73 (±0·07) | 0·55 (±0·04) | 0·48 (±0·03) | 0·87 (±0·01) | 0·45 (±0·02) | 0·16 | 0·97 |
| RNF43 | 0·77 (±0·04) | 0·5 (±0·05) | 0·82 (±0·06) | 0·76 (±0·06) | 0·62 (±0·02) | 0·5 (±0·03) | 0·86 (±0·01) | 0·59 (±0·02) | 0·22 | 0·86 |
| FHOD3 | 0·79 (±0·06) | 0·33 (±0·06) | 0·77 (±0·14) | 0·79 (±0·08) | 0·45 (±0·04) | 0·4 (±0·04) | 0·84 (±0·02) | 0·38 (±0·03) | 0·06 | 0·81 |
| WNT16 | 0·77 (±0·08) | 0·14 (±0·04) | 0·76 (±0·14) | 0·77 (±0·09) | 0·24 (±0·05) | 0·26 (±0·04) | 0·82 (±0·02) | 0·18 (±0·02) | 0·02 | 0·93 |
| ELL2 | 0·71 (±0·08) | 0·03 (±0·0) | 0·75 (±0·14) | 0·71 (±0·09) | 0·06 (±0·01) | 0·11 (±0·01) | 0·79 (±0·05) | 0·06 (±0·01) | 0·01 | 0·75 |
| MECOM | 0·75 (±0·05) | 0·20 (±0·04) | 0·70 (±0·09) | 0·75 (±0·06) | 0·31 (±0·05) | 0·28 (±0·07) | 0·78 (±0·04) | 0·21 (±0·04) | 0·04 | 0·88 |
| BRAF | 0·73 (±0·03) | 0·42 (±0·03) | 0·73 (±0·08) | 0·73 (±0·05) | 0·53 (±0·01) | 0·39 (±0·01) | 0·78 (±0·01) | 0·45 (±0·02) | 0·21 | 0·71 |
| CDK12 | 0·60 (±0·07) | 0·09 (±0·01) | 0·86 (±0·07) | 0·58 (±0·08) | 0·16 (±0·01) | 0·19 (±0·01) | 0·78 (±0·02) | 0·13 (±0·02) | 0·04 | 0·86 |
| TGFBR2 | 0·64 (±0·07) | 0·14 (±0·01) | 0·81 (±0·11) | 0·63 (±0·08) | 0·24 (±0·02) | 0·23 (±0·02) | 0·77 (±0·02) | 0·17 (±0·01) | 0·07 | 0·73 |
| RFX5 | 0·71 (±0·09) | 0·11 (±0·02) | 0·77 (±0·11) | 0·71 (±0·1) | 0·18 (±0·03) | 0·21 (±0·04) | 0·77 (±0·02) | 0·12 (±0·01) | 0·02 | 0·85 |
| CCDC40 | 0·63 (±0·1) | 0·07 (±0·01) | 0·76 (±0·22) | 0·62 (±0·12) | 0·13 (±0·01) | 0·15 (±0·04) | 0·76 (±0·04) | 0·1 (±0·03) | 0·04 | 0·83 |
| SMG1 | 0·58 (±0·13) | 0·08 (±0·02) | 0·82 (±0·16) | 0·57 (±0·14) | 0·14 (±0·04) | 0·16 (±0·06) | 0·76 (±0·07) | 0·1 (±0·02) | 0·04 | 0·77 |

| | | | | | | | | | | |
|---|---|---|---|---|---|---|---|---|---|---|
| ACVR1B | 0·61 (±0·08) | 0·11 (±0·01) | 0·78 (±0·11) | 0·60 (±0·09) | 0·2 (±0·02) | 0·18 (±0·03) | 0·75 (±0·03) | 0·17 (±0·01) | 0·06 | 0·51 |
| ZNRF3 | 0·70 (±0·04) | 0·23 (±0·02) | 0·73 (±0·05) | 0·69 (±0·05) | 0·34 (±0·02) | 0·28 (±0·02) | 0·75 (±0·01) | 0·25 (±0·01) | 0·11 | 0·87 |
| CTNND1 | 0·67 (±0·07) | 0·09 (±0·02) | 0·74 (±0·06) | 0·66 (±0·07) | 0·17 (±0·03) | 0·18 (±0·04) | 0·75 (±0·03) | 0·1 (±0·01) | 0·04 | 0·89 |
| ZHX2 | 0·6 (±0·06) | 0·07 (±0·0) | 0·81 (±0·13) | 0·59 (±0·06) | 0·12 (±0·01) | 0·15 (±0·03) | 0·74 (±0·02) | 0·08 (±0·01) | 0·03 | 0·77 |
| TBX3 | 0·53 (±0·13) | 0·1 (±0·02) | 0·83 (±0·11) | 0·51 (±0·14) | 0·17 (±0·02) | 0·16 (±0·03) | 0·74 (±0·03) | 0·14 (±0·03) | 0·06 | 0·76 |
| DUSP16 | 0·46 (±0·18) | 0·04 (±0·01) | 0·83 (±0·11) | 0·45 (±0·18) | 0·07 (±0·02) | 0·09 (±0·04) | 0·74 (±0·04) | 0·08 (±0·03) | 0·02 | 0·75 |
| TRPS1 | 0·48 (±0·17) | 0·14 (±0·05) | 0·8 (±0·07) | 0·45 (±0·2) | 0·23 (±0·07) | 0·15 (±0·09) | 0·73 (±0·03) | 0·24 (±0·05) | 0·04 | 0·68 |
| TP53 | 0·65 (±0·01) | 0·7 (±0·05) | 0·61 (±0·15) | 0·68 (±0·17) | 0·64 (±0·05) | 0·31 (±0·03) | 0·72 (±0·02) | 0·72 (±0·03) | 0·53 | 0·13 |
| AKT1 | 0·48 (±0·17) | 0·07 (±0·02) | 0·81 (±0·12) | 0·47 (±0·18) | 0·13 (±0·03) | 0·12 (±0·04) | 0·7 (±0·03) | 0·1 (±0·02) | 0·05 | 0·63 |
| ALK | 0·43 (±0·23) | 0·12 (±0·03) | 0·84 (±0·14) | 0·39 (±0·26) | 0·21 (±0·04) | 0·14 (±0·06) | 0·7 (±0·02) | 0·17 (±0·02) | 0·08 | 0·77 |
| KIF1A | 0·46 (±0·19) | 0·14 (±0·02) | 0·8 (±0·14) | 0·42 (±0·23) | 0·23 (±0·03) | 0·14 (±0·05) | 0·67 (±0·03) | 0·19 (±0·03) | 0·10 | 0·63 |
| APC | 0·63 (±0·06) | 0·7 (±0·05) | 0·68 (±0·23) | 0·55 (±0·2) | 0·66 (±0·15) | 0·25 (±0·04) | 0·66 (±0·02) | 0·71 (±0·04) | 0·59 | 0·15 |
| SMAD2 | 0·41 (±0·16) | 0·07 (±0·01) | 0·8 (±0·11) | 0·39 (±0·17) | 0·13 (±0·02) | 0·09 (±0·03) | 0·65 (±0·03) | 0·11 (±0·03) | 0·05 | 0·39 |
| KRAS | 0·61 (±0·04) | 0·44 (±0·03) | 0·57 (±0·19) | 0·63 (±0·15) | 0·48 (±0·07) | 0·19 (±0·04) | 0·65 (±0·03) | 0·46 (±0·03) | 0·33 | 0·08 |
| ATM | 0·61 (±0·08) | 0·14 (±0·02) | 0·62 (±0·07) | 0·61 (±0·1) | 0·23 (±0·02) | 0·14 (±0·03) | 0·63 (±0·01) | 0·15 (±0·01) | 0·09 | 0·63 |
| PIK3CA | 0·49 (±0·09) | 0·19 (±0·02) | 0·66 (±0·14) | 0·46 (±0·14) | 0·29 (±0·02) | 0·09 (±0·04) | 0·56 (±0·03) | 0·19 (±0·02) | 0·16 | 0·42 |
| NRAS | 0·58 (±0·14) | 0·05 (±0·01) | 0·49 (±0·21) | 0·59 (±0·15) | 0·09 (±0·02) | 0·03 (±0·03) | 0·56 (±0·04) | 0·06 (±0·01) | 0·04 | 0·07 |

**Tab. S15: Comparison of the Microsatellite Instability (MSI) prediction scores with the target prediction scores within subgroups.** The data is organized by subgroups (Fig. 1D), with each subgroup representing a specific category such as MSI, Microsatellite Stability (MSS), Prediction Target Wild Type (WT), and Prediction Target Mutated (MUT). For each subgroup, the table provides the count of samples (N) and the median values for the prediction scores for both the train set (internal, i) and test set (external, e). The p-value from the Mann-Whitney U test is also included, which indicates the statistical significance of the comparison between the MSI and prediction target scores within each subgroup. This data is visualized within Fig. 4 and Figs. S4-S5.

| Target | i/e | MSS/WT_N | MSS/WT_p | MSS/WT_MSI_median | MSS/WT_target_median | MSI/WT_N | MSI/WT_p | MSI/WT_MSI_median | MSI/WT_target_median | MSS/MUT_N | MSS/MUT_p | MSS/MUT_MSI_median | MSS/MUT_target_median | MSI/MUT_N | MSI/MUT_p | MSI/MUT_MSI_median | MSI/MUT_target_median |
|---|---|---|---|---|---|---|---|---|---|---|---|---|---|---|---|---|---|
| ACVR1B | i | 563 | 6·01e-40 | 0·01 | 0·08 | 133 | 1·04e-7 | 0·95 | 0·46 | 10 | 4·92e-1 | 0·02 | 0·13 | 25 | 2·26e-3 | 0·92 | 0·60 |
| | e | 449 | 1·30e-63 | 0·12 | 0·29 | 157 | 3·06e-13 | 0·89 | 0·78 | 19 | 2·93e-1 | 0·69 | 0·62 | 20 | 1·99e-3 | 0·93 | 0·81 |
| AKT1 | i | 563 | 2·37e-39 | 0·01 | 0·10 | 146 | 1·24e-8 | 0·91 | 0·36 | 10 | 1·37e-2 | 0·00 | 0·51 | 12 | 1·61e-2 | 0·99 | 0·50 |
| | e | 457 | 5·90e-66 | 0·12 | 0·44 | 158 | 3·74e-15 | 0·89 | 0·67 | 11 | 9·77e-4 | 0·12 | 0·53 | 19 | 1·69e-3 | 0·93 | 0·69 |
| ALK | i | 551 | 2·76e-48 | 0·01 | 0·19 | 128 | 2·11e-8 | 0·95 | 0·47 | 22 | 4·28e-4 | 0·00 | 0·22 | 30 | 6·85e-1 | 0·77 | 0·56 |
| | e | 456 | 3·03e-72 | 0·12 | 0·49 | 136 | 2·35e-10 | 0·90 | 0·76 | 12 | 2·44e-3 | 0·25 | 0·53 | 41 | 1·66e-2 | 0·83 | 0·74 |
| APC | i | 196 | 2·80e-22 | 0·01 | 0·54 | 103 | 1·09e-9 | 0·95 | 0·21 | 377 | 1·59e-43 | 0·01 | 0·66 | 55 | 3·70e-2 | 0·94 | 0·31 |
| | e | 146 | 7·75e-17 | 0·14 | 0·62 | 118 | 8·35e-19 | 0·91 | 0·33 | 322 | 4·96e-43 | 0·11 | 0·66 | 59 | 1·10e-8 | 0·88 | 0·39 |
| ATM | i | 530 | 2·57e-45 | 0·01 | 0·16 | 111 | 1·60e-4 | 0·89 | 0·53 | 43 | 1·83e-4 | 0·02 | 0·20 | 47 | 1·60e-3 | 0·98 | 0·54 |
| | e | 446 | 7·23e-62 | 0·12 | 0·37 | 140 | 1·18e-17 | 0·90 | 0·72 | 22 | 2·62e-5 | 0·09 | 0·32 | 37 | 4·55e-6 | 0·89 | 0·72 |
| BMPR2 | i | 566 | 2·16e-8 | 0·01 | 0·02 | 60 | 1·55e-5 | 0·74 | 0·40 | 7 | 9·38e-1 | 0·01 | 0·04 | 98 | 6·51e-6 | 0·97 | 0·83 |
| | e | 465 | 2·60e-36 | 0·12 | 0·16 | 74 | 2·61e-5 | 0·92 | 0·88 | 3 | 5·00e-1 | 0·54 | 0·58 | 103 | 9·24e-9 | 0·89 | 0·82 |
| BRAF | i | 510 | 2·30e-37 | 0·00 | 0·06 | 41 | 2·91e-1 | 0·53 | 0·72 | 63 | 5·89e-8 | 0·03 | 0·44 | 117 | 2·05e-1 | 0·97 | 0·91 |
| | e | 429 | 5·02e-49 | 0·11 | 0·22 | 81 | 1·22e-7 | 0·90 | 0·85 | 39 | 5·59e-4 | 0·23 | 0·34 | 96 | 9·23e-11 | 0·89 | 0·83 |
| CCDC40 | i | 565 | 8·85e-38 | 0·01 | 0·06 | 136 | 2·03e-9 | 0·94 | 0·49 | 8 | 2·50e-1 | 0·01 | 0·38 | 22 | 1·09e-3 | 0·98 | 0·53 |
| | e | 464 | 4·18e-56 | 0·12 | 0·33 | 158 | 7·49e-21 | 0·90 | 0·70 | 4 | 1·25e-1 | 0·34 | 0·52 | 19 | 6·63e-2 | 0·83 | 0·66 |
| CDK12 | i | 567 | 6·88e-34 | 0·01 | 0·07 | 141 | 9·54e-12 | 0·95 | 0·45 | 6 | 3·13e-1 | 0·01 | 0·26 | 17 | 5·05e-2 | 0·90 | 0·51 |
| | e | 464 | 5·30e-68 | 0·12 | 0·36 | 152 | 8·54e-18 | 0·89 | 0·74 | 4 | 1·25e-1 | 0·39 | 0·53 | 25 | 2·03e-3 | 0·90 | 0·72 |
| CHD1 | i | 417 | 1·46e-19 | 0·01 | 0·07 | 108 | 9·83e-7 | 0·94 | 0·49 | 9 | 1·00e0 | 0·13 | 0·23 | 39 | 1·58e-4 | 0·99 | 0·68 |
| | e | 255 | 5·36e-16 | 0·13 | 0·20 | 55 | 6·27e-10 | 0·86 | 0·65 | 1 | 1·00e0 | 0·30 | 0·31 | 10 | 1·95e-3 | 0·87 | 0·67 |
| CTNND1 | i | 568 | 3·09e-27 | 0·01 | 0·05 | 134 | 6·32e-12 | 0·94 | 0·42 | 5 | 3·13e-1 | 0·23 | 0·03 | 24 | 3·66e-5 | 0·97 | 0·42 |
| | e | 465 | 9·42e-44 | 0·12 | 0·27 | 152 | 8·00e-24 | 0·90 | 0·74 | 3 | 1·00e0 | 0·38 | 0·38 | 25 | 1·49e-6 | 0·88 | 0·72 |
| DUSP16 | i | 564 | 2·71e-34 | 0·01 | 0·12 | 147 | 5·14e-12 | 0·94 | 0·47 | 9 | 3·91e-3 | 0·00 | 0·17 | 11 | 1·86e-2 | 0·99 | 0·38 |
| | e | 464 | 7·34e-72 | 0·12 | 0·47 | 165 | 2·66e-16 | 0·89 | 0·73 | 4 | 1·25e-1 | 0·21 | 0·51 | 12 | 9·77e-4 | 0·91 | 0·74 |
| ELL2 | i | 423 | 8·96e-14 | 0·01 | 0·07 | 131 | 9·37e-12 | 0·96 | 0·53 | 3 | 7·50e-1 | 0·03 | 0·39 | 16 | 7·63e-3 | 0·99 | 0·34 |
| | e | 255 | 9·70e-26 | 0·13 | 0·27 | 62 | 1·13e-8 | 0·86 | 0·68 | 1 | 1·00e0 | 0·30 | 0·28 | 3 | 2·50e-1 | 0·86 | 0·74 |
| FHOD3 | i | 420 | 3·87e-15 | 0·01 | 0·05 | 79 | 1·36e-8 | 0·95 | 0·54 | 6 | 1·00e0 | 0·42 | 0·15 | 68 | 1·21e-5 | 0·97 | 0·65 |
| | e | 249 | 2·34e-18 | 0·13 | 0·19 | 36 | 3·11e-8 | 0·84 | 0·73 | 7 | 1·00e0 | 0·30 | 0·39 | 29 | 3·73e-9 | 0·87 | 0·75 |
| hypermutated | i | 551 | 1·73e-28 | 0·01 | 0·02 | 5 | 3·13e-1 | 1·00 | 0·98 | 22 | 2·38e-6 | 0·00 | 0·04 | 153 | 2·66e-2 | 0·94 | 0·96 |
| | e | 454 | 3·14e-68 | 0·12 | 0·22 | 13 | 1·71e-3 | 0·96 | 0·93 | 14 | 1·22e-4 | 0·10 | 0·16 | 164 | 8·27e-1 | 0·89 | 0·88 |
| KIF1A | i | 540 | 8·89e-41 | 0·01 | 0·14 | 111 | 1·25e-4 | 0·94 | 0·50 | 33 | 5·63e-6 | 0·00 | 0·31 | 47 | 8·50e-3 | 0·97 | 0·48 |
| | e | 445 | 3·38e-70 | 0·12 | 0·45 | 137 | 1·63e-10 | 0·90 | 0·77 | 23 | 1·67e-5 | 0·25 | 0·56 | 40 | 5·54e-1 | 0·84 | 0·78 |
| KRAS | i | 349 | 3·12e-34 | 0·00 | 0·35 | 142 | 6·33e-12 | 0·97 | 0·10 | 224 | 5·54e-25 | 0·01 | 0·51 | 16 | 7·82e-1 | 0·16 | 0·50 |
| | e | 271 | 3·56e-30 | 0·11 | 0·51 | 160 | 5·78e-25 | 0·89 | 0·32 | 197 | 8·63e-22 | 0·13 | 0·54 | 17 | 4·58e-5 | 0·91 | 0·36 |
| MECOM | i | 420 | 1·52e-10 | 0·01 | 0·04 | 103 | 1·96e-9 | 0·95 | 0·58 | 6 | 1·00e0 | 0·07 | 0·08 | 44 | 2·01e-5 | 0·97 | 0·64 |
| | e | 253 | 3·81e- | 0·13 | 0·22 | 42 | 3·95e-9 | 0·87 | 0·74 | 3 | 2·50e-1 | 0·17 | 0·24 | 23 | 2·15e-4 | 0·82 | 0·70 |

| | | | | | | | | | | | | | | | | | |
|---|---|---|---|---|---|---|---|---|---|---|---|---|---|---|---|---|---|
| | | | 27 | | | | | | | | | | | | | | |
| NRAS | i | 542 | 1·83e-35 | 0·01 | 0·20 | 156 | 1·30e-16 | 0·94 | 0·12 | 31 | 5·41e-4 | 0·00 | 0·23 | 2 | 1·00e0 | 0·50 | 0·03 |
| | e | 442 | 1·96e-44 | 0·12 | 0·50 | 175 | 4·27e-28 | 0·90 | 0·34 | 26 | 8·35e-4 | 0·12 | 0·49 | 2 | 1·00e0 | 0·30 | 0·51 |
| PIK3CA | i | 486 | 1·18e-32 | 0·01 | 0·09 | 96 | 1·09e-4 | 0·93 | 0·45 | 87 | 1·68e-7 | 0·01 | 0·16 | 62 | 2·89e-5 | 0·97 | 0·48 |
| | e | 407 | 1·82e-59 | 0·12 | 0·45 | 133 | 8·83e-17 | 0·91 | 0·71 | 61 | 4·85e-10 | 0·12 | 0·53 | 44 | 4·81e-5 | 0·83 | 0·66 |
| PLEKHA6 | i | 418 | 7·19e-19 | 0·01 | 0·07 | 97 | 1·85e-8 | 0·94 | 0·54 | 8 | 7·81e-3 | 0·03 | 0·19 | 50 | 2·00e-7 | 0·99 | 0·65 |
| | e | 255 | 1·74e-33 | 0·13 | 0·24 | 37 | 1·63e-4 | 0·82 | 0·72 | 1 | 1·00e0 | 0·95 | 0·83 | 28 | 1·49e-8 | 0·89 | 0·75 |
| RFX5 | i | 425 | 1·49e-8 | 0·01 | 0·03 | 120 | 1·93e-12 | 0·95 | 0·50 | 1 | 1·00e0 | 0·13 | 0·01 | 27 | 2·52e-6 | 0·99 | 0·71 |
| | e | 254 | 1·84e-31 | 0·13 | 0·24 | 54 | 9·01e-7 | 0·86 | 0·74 | 2 | 5·00e-1 | 0·07 | 0·16 | 11 | 9·77e-4 | 0·86 | 0·72 |
| RNF43 | i | 546 | 1·16e-28 | 0·01 | 0·03 | 45 | 2·62e-1 | 0·79 | 0·69 | 27 | 1·51e-2 | 0·02 | 0·16 | 113 | 1·04e-2 | 0·97 | 0·86 |
| | e | 448 | 2·19e-41 | 0·12 | 0·17 | 54 | 2·53e-6 | 0·86 | 0·83 | 20 | 4·41e-2 | 0·29 | 0·36 | 123 | 1·74e-13 | 0·90 | 0·83 |
| SMAD2 | i | 558 | 1·13e-53 | 0·01 | 0·17 | 145 | 1·90e-11 | 0·97 | 0·47 | 15 | 4·21e-1 | 0·04 | 0·37 | 13 | 5·42e-1 | 0·04 | 0·14 |
| | e | 448 | 7·36e-72 | 0·12 | 0·48 | 164 | 3·06e-7 | 0·89 | 0·77 | 20 | 1·34e-4 | 0·34 | 0·65 | 13 | 4·64e-3 | 0·90 | 0·78 |
| SMG1 | i | 561 | 3·41e-39 | 0·01 | 0·11 | 138 | 4·95e-11 | 0·95 | 0·51 | 12 | 3·01e-1 | 0·01 | 0·16 | 20 | 8·26e-2 | 0·41 | 0·35 |
| | e | 462 | 3·20e-65 | 0·12 | 0·38 | 157 | 7·87e-19 | 0·89 | 0·69 | 6 | 3·13e-1 | 0·43 | 0·57 | 20 | 2·67e-5 | 0·92 | 0·69 |
| TBX3 | i | 561 | 9·45e-48 | 0·01 | 0·11 | 139 | 4·82e-9 | 0·95 | 0·42 | 12 | 6·77e-1 | 0·03 | 0·23 | 19 | 9·45e-3 | 0·89 | 0·24 |
| | e | 459 | 1·69e-69 | 0·12 | 0·43 | 149 | 5·45e-10 | 0·89 | 0·74 | 9 | 1·95e-2 | 0·37 | 0·55 | 28 | 7·20e-5 | 0·92 | 0·74 |
| TGFBR2 | i | 564 | 2·92e-44 | 0·01 | 0·07 | 128 | 9·85e-10 | 0·95 | 0·50 | 9 | 3·91e-3 | 0·01 | 0·27 | 30 | 5·01e-3 | 0·91 | 0·54 |
| | e | 456 | 7·01e-62 | 0·12 | 0·27 | 145 | 2·79e-14 | 0·89 | 0·77 | 12 | 3·42e-3 | 0·40 | 0·52 | 32 | 5·43e-5 | 0·90 | 0·76 |
| TP53 | i | 174 | 1·01e-16 | 0·01 | 0·50 | 105 | 1·23e-5 | 0·89 | 0·21 | 399 | 4·77e-56 | 0·00 | 0·76 | 53 | 2·48e-2 | 0·97 | 0·43 |
| | e | 170 | 1·54e-8 | 0·16 | 0·48 | 133 | 6·73e-22 | 0·90 | 0·27 | 298 | 5·23e-41 | 0·10 | 0·58 | 44 | 2·80e-9 | 0·89 | 0·31 |
| TRPS1 | i | 406 | 2·38e-38 | 0·01 | 0·22 | 118 | 1·82e-7 | 0·97 | 0·52 | 20 | 2·96e-2 | 0·02 | 0·38 | 29 | 6·23e-2 | 0·78 | 0·47 |
| | e | 247 | 3·52e-40 | 0·13 | 0·49 | 46 | 6·84e-3 | 0·82 | 0·71 | 9 | 7·81e-3 | 0·12 | 0·54 | 19 | 9·54e-5 | 0·90 | 0·73 |
| WNT16 | i | 425 | 3·11e-4 | 0·01 | 0·03 | 121 | 2·75e-13 | 0·95 | 0·49 | 1 | 1·00e0 | 0·27 | 0·12 | 26 | 5·66e-7 | 0·97 | 0·48 |
| | e | 255 | 9·02e-12 | 0·13 | 0·19 | 52 | 1·39e-9 | 0·86 | 0·68 | 1 | 1·00e0 | 0·02 | 0·08 | 13 | 2·44e-4 | 0·87 | 0·67 |
| ZHX2 | i | 564 | 2·09e-32 | 0·01 | 0·07 | 143 | 4·03e-12 | 0·95 | 0·41 | 9 | 4·26e-1 | 0·00 | 0·17 | 15 | 2·29e-1 | 0·40 | 0·43 |
| | e | 463 | 1·18e-53 | 0·12 | 0·33 | 160 | 2·43e-19 | 0·89 | 0·69 | 5 | 6·25e-1 | 0·48 | 0·55 | 17 | 6·56e-4 | 0·92 | 0·68 |
| ZNRF3 | i | 565 | 3·83e-26 | 0·01 | 0·05 | 87 | 2·47e-4 | 0·87 | 0·52 | 8 | 3·83e-1 | 0·01 | 0·03 | 71 | 6·68e-6 | 0·99 | 0·65 |
| | e | 459 | 1·66e-43 | 0·12 | 0·21 | 116 | 3·27e-16 | 0·90 | 0·80 | 9 | 3·91e-3 | 0·07 | 0·17 | 61 | 4·04e-10 | 0·89 | 0·76 |

**Tab. S16: Comparison of microsatellite instability (MSI) scores and prediction target scores among subgroups.** The results are arranged by subgroups (Fig. 1D), where each subgroup represents a particular combination of microsatellite and prediction target mutation status, such as MSI, Microsatellite Stability (MSS), Prediction Target Wild Type (WT) and Prediction Target Mutated (MUT). For each subgroup within the train and test dataset, the table provides the p-value obtained from the Wilcoxon test for comparing MSI and prediction target scores, respectively, which is shown in the second column. These results are illustrated in Fig. 4 and Figs. S4-S5.

| | | Train set | | Test set | |
|---|---|---|---|---|---|
| **Target** | **comparison** | **p_value_MSI** | **p_value_target** | **p_value_MSI** | **p_value_target** |
| ACVR1B | MSS/WT vs MSI/WT | 0·00e0 | 0·00e0 | 0·00e0 | 0·00e0 |
| | MSS/WT vs MSS/MUT | 1·05e-1 | 1·25e-1 | 5·00e-5 | 1·00e-5 |
| | MSS/WT vs MSI/MUT | 0·00e0 | 0·00e0 | 0·00e0 | 0·00e0 |
| | MSI/WT vs MSS/MUT | 9·10e-3 | 7·69e-2 | 9·00e-5 | 5·60e-4 |
| | MSI/WT vs MSI/MUT | 6·00e-1 | 8·19e-1 | 1·29e-1 | 1·52e-1 |
| | MSS/MUT vs MSI/MUT | 1·85e-2 | 1·21e-1 | 4·20e-4 | 8·70e-4 |
| AKT1 | MSS/WT vs MSI/WT | 0·00e0 | 0·00e0 | 0·00e0 | 0·00e0 |
| | MSS/WT vs MSS/MUT | 5·51e-1 | 7·50e-3 | 3·86e-1 | 1·94e-2 |
| | MSS/WT vs MSI/MUT | 0·00e0 | 7·00e-5 | 0·00e0 | 0·00e0 |
| | MSI/WT vs MSS/MUT | 1·60e-4 | 7·92e-1 | 0·00e0 | 4·80e-4 |
| | MSI/WT vs MSI/MUT | 1·61e-1 | 3·17e-1 | 8·48e-1 | 3·49e-1 |
| | MSS/MUT vs MSI/MUT | 5·40e-4 | 1·00e0 | 5·00e-5 | 1·45e-3 |
| ALK | MSS/WT vs MSI/WT | 0·00e0 | 0·00e0 | 0·00e0 | 0·00e0 |
| | MSS/WT vs MSS/MUT | 7·79e-1 | 3·19e-1 | 1·31e-1 | 2·62e-1 |
| | MSS/WT vs MSI/MUT | 0·00e0 | 0·00e0 | 0·00e0 | 0·00e0 |
| | MSI/WT vs MSS/MUT | 0·00e0 | 8·22e-3 | 0·00e0 | 0·00e0 |
| | MSI/WT vs MSI/MUT | 3·31e-1 | 5·51e-1 | 2·67e-2 | 7·31e-2 |
| | MSS/MUT vs MSI/MUT | 6·00e-5 | 7·44e-3 | 1·00e-5 | 2·00e-5 |
| APC | MSS/WT vs MSI/WT | 0·00e0 | 0·00e0 | 0·00e0 | 0·00e0 |
| | MSS/WT vs MSS/MUT | 3·98e-1 | 1·24e-2 | 2·26e-1 | 3·76e-2 |
| | MSS/WT vs MSI/MUT | 0·00e0 | 5·23e-2 | 0·00e0 | 0·00e0 |
| | MSI/WT vs MSS/MUT | 0·00e0 | 0·00e0 | 0·00e0 | 0·00e0 |
| | MSI/WT vs MSI/MUT | 3·48e-1 | 1·60e-1 | 6·80e-2 | 1·09e-2 |
| | MSS/MUT vs MSI/MUT | 0·00e0 | 8·40e-4 | 0·00e0 | 0·00e0 |
| ATM | MSS/WT vs MSI/WT | 0·00e0 | 0·00e0 | 0·00e0 | 0·00e0 |
| | MSS/WT vs MSS/MUT | 3·30e-2 | 7·90e-1 | 2·45e-1 | 1·30e-1 |
| | MSS/WT vs MSI/MUT | 0·00e0 | 0·00e0 | 0·00e0 | 0·00e0 |
| | MSI/WT vs MSS/MUT | 0·00e0 | 0·00e0 | 0·00e0 | 0·00e0 |
| | MSI/WT vs MSI/MUT | 1·65e-1 | 4·84e-1 | 3·27e-1 | 5·41e-1 |
| | MSS/MUT vs MSI/MUT | 0·00e0 | 2·00e-5 | 0·00e0 | 0·00e0 |
| BMPR2 | MSS/WT vs MSI/WT | 0·00e0 | 0·00e0 | 0·00e0 | 0·00e0 |
| | MSS/WT vs MSS/MUT | 4·07e-1 | 2·62e-1 | 8·88e-3 | 8·14e-3 |
| | MSS/WT vs MSI/MUT | 0·00e0 | 0·00e0 | 0·00e0 | 0·00e0 |
| | MSI/WT vs MSS/MUT | 3·00e-2 | 7·95e-2 | 2·85e-2 | 2·85e-2 |
| | MSI/WT vs MSI/MUT | 4·06e-2 | 2·93e-2 | 1·03e-1 | 6·90e-2 |
| | MSS/MUT vs MSI/MUT | 9·90e-4 | 1·05e-3 | 3·93e-2 | 3·49e-2 |
| BRAF | MSS/WT vs MSI/WT | 0·00e0 | 0·00e0 | 0·00e0 | 0·00e0 |
| | MSS/WT vs MSS/MUT | 2·50e-4 | 0·00e0 | 2·70e-4 | 5·02e-3 |
| | MSS/WT vs MSI/MUT | 0·00e0 | 0·00e0 | 0·00e0 | 0·00e0 |
| | MSI/WT vs MSS/MUT | 2·12e-3 | 1·60e-1 | 0·00e0 | 0·00e0 |
| | MSI/WT vs MSI/MUT | 3·88e-2 | 3·80e-2 | 9·80e-1 | 8·40e-1 |
| | MSS/MUT vs MSI/MUT | 0·00e0 | 1·00e-5 | 0·00e0 | 0·00e0 |
| CCDC40 | MSS/WT vs MSI/WT | 0·00e0 | 0·00e0 | 0·00e0 | 0·00e0 |
| | MSS/WT vs MSS/MUT | 6·97e-1 | 6·19e-2 | 3·93e-2 | 2·60e-2 |
| | MSS/WT vs MSI/MUT | 0·00e0 | 1·60e-4 | 0·00e0 | 0·00e0 |
| | MSI/WT vs MSS/MUT | 5·82e-3 | 3·07e-1 | 1·80e-4 | 8·10e-4 |
| | MSI/WT vs MSI/MUT | 7·35e-1 | 9·86e-1 | 3·14e-1 | 5·91e-1 |
| | MSS/MUT vs MSI/MUT | 3·49e-2 | 4·75e-1 | 8·58e-3 | 8·58e-3 |
| CDK12 | MSS/WT vs MSI/WT | 0·00e0 | 0·00e0 | 0·00e0 | 0·00e0 |
| | MSS/WT vs MSS/MUT | 9·24e-1 | 2·61e-1 | 3·61e-2 | 3·61e-2 |
| | MSS/WT vs MSI/MUT | 0·00e0 | 5·00e-5 | 0·00e0 | 0·00e0 |
| | MSI/WT vs MSS/MUT | 3·70e-2 | 1·83e-1 | 4·70e-4 | 1·48e-3 |
| | MSI/WT vs MSI/MUT | 9·64e-1 | 9·28e-1 | 8·94e-1 | 9·92e-1 |
| | MSS/MUT vs MSI/MUT | 1·58e-2 | 1·77e-1 | 2·27e-3 | 2·27e-3 |
| CHD1 | MSS/WT vs MSI/WT | 0·00e0 | 0·00e0 | 0·00e0 | 0·00e0 |
| | MSS/WT vs MSS/MUT | 1·83e-2 | 1·11e-1 | 4·92e-1 | 5·70e-1 |
| | MSS/WT vs MSI/MUT | 0·00e0 | 0·00e0 | 0·00e0 | 0·00e0 |
| | MSI/WT vs MSS/MUT | 7·10e-2 | 1·48e-1 | 1·43e-1 | 1·43e-1 |
| | MSI/WT vs MSI/MUT | 3·50e-2 | 7·10e-2 | 4·95e-1 | 2·79e-1 |
| | MSS/MUT vs MSI/MUT | 6·01e-3 | 2·65e-2 | 1·82e-1 | 1·82e-1 |
| CTNND1 | MSS/WT vs MSI/WT | 0·00e0 | 0·00e0 | 0·00e0 | 0·00e0 |
| | MSS/WT vs MSS/MUT | 1·94e-1 | 8·61e-1 | 2·68e-1 | 3·79e-1 |
| | MSS/WT vs MSI/MUT | 0·00e0 | 0·00e0 | 0·00e0 | 0·00e0 |
| | MSI/WT vs MSS/MUT | 3·38e-1 | 1·01e-1 | 7·46e-2 | 5·70e-2 |
| | MSI/WT vs MSI/MUT | 4·60e-1 | 4·63e-1 | 3·34e-1 | 1·64e-1 |
| | MSS/MUT vs MSI/MUT | 2·01e-1 | 7·79e-2 | 1·25e-1 | 1·67e-1 |
| DUSP16 | MSS/WT vs MSI/WT | 0·00e0 | 0·00e0 | 0·00e0 | 0·00e0 |
| | MSS/WT vs MSS/MUT | 9·73e-1 | 6·27e-1 | 6·30e-1 | 6·98e-1 |
| | MSS/WT vs MSI/MUT | 1·00e-5 | 3·04e-3 | 0·00e0 | 0·00e0 |
| | MSI/WT vs MSS/MUT | 5·30e-4 | 1·70e-2 | 1·20e-4 | 1·30e-4 |
| | MSI/WT vs MSI/MUT | 3·86e-1 | 7·59e-1 | 4·75e-1 | 3·14e-1 |

| | | | | | |
|---|---|---|---|---|---|
| | MSS/MUT vs MSI/MUT | 1·84e-3 | 9·46e-2 | 1·10e-3 | 1·10e-3 |
| ELL2 | MSS/WT vs MSI/WT | 0·00e0 | 0·00e0 | 0·00e0 | 0·00e0 |
| | MSS/WT vs MSS/MUT | 2·12e-1 | 6·50e-2 | 4·92e-1 | 9·14e-1 |
| | MSS/WT vs MSI/MUT | 0·00e0 | 0·00e0 | 1·70e-4 | 1·40e-4 |
| | MSI/WT vs MSS/MUT | 1·96e-1 | 4·99e-1 | 1·27e-1 | 6·35e-2 |
| | MSI/WT vs MSI/MUT | 6·39e-1 | 7·96e-1 | 5·41e-1 | 2·81e-1 |
| | MSS/MUT vs MSI/MUT | 1·09e-1 | 6·34e-1 | 5·00e-1 | 5·00e-1 |
| FHOD3 | MSS/WT vs MSI/WT | 0·00e0 | 0·00e0 | 0·00e0 | 0·00e0 |
| | MSS/WT vs MSS/MUT | 4·85e-1 | 3·54e-1 | 5·09e-2 | 4·77e-2 |
| | MSS/WT vs MSI/MUT | 0·00e0 | 0·00e0 | 0·00e0 | 0·00e0 |
| | MSI/WT vs MSS/MUT | 6·96e-2 | 1·66e-1 | 2·40e-2 | 2·40e-2 |
| | MSI/WT vs MSI/MUT | 3·79e-1 | 1·88e-1 | 4·25e-1 | 6·30e-1 |
| | MSS/MUT vs MSI/MUT | 4·69e-2 | 6·31e-2 | 3·37e-3 | 5·34e-3 |
| hypermutated | MSS/WT vs MSI/WT | 8·48e-3 | 2·38e-2 | 0·00e0 | 0·00e0 |
| | MSS/WT vs MSS/MUT | 9·29e-2 | 3·41e-2 | 8·56e-1 | 7·66e-1 |
| | MSS/WT vs MSI/MUT | 0·00e0 | 0·00e0 | 0·00e0 | 0·00e0 |
| | MSI/WT vs MSS/MUT | 1·26e-2 | 1·29e-1 | 1·00e-5 | 1·00e-5 |
| | MSI/WT vs MSI/MUT | 7·29e-1 | 1·00e0 | 1·55e-2 | 2·23e-2 |
| | MSS/MUT vs MSI/MUT | 0·00e0 | 6·00e-5 | 0·00e0 | 0·00e0 |
| KIF1A | MSS/WT vs MSI/WT | 0·00e0 | 0·00e0 | 0·00e0 | 0·00e0 |
| | MSS/WT vs MSS/MUT | 2·45e-1 | 6·07e-3 | 8·65e-2 | 1·43e-1 |
| | MSS/WT vs MSI/MUT | 0·00e0 | 0·00e0 | 0·00e0 | 0·00e0 |
| | MSI/WT vs MSS/MUT | 0·00e0 | 1·07e-1 | 0·00e0 | 0·00e0 |
| | MSI/WT vs MSI/MUT | 8·22e-1 | 9·58e-1 | 1·31e-1 | 7·30e-1 |
| | MSS/MUT vs MSI/MUT | 0·00e0 | 1·20e-1 | 0·00e0 | 0·00e0 |
| KRAS | MSS/WT vs MSI/WT | 0·00e0 | 0·00e0 | 0·00e0 | 0·00e0 |
| | MSS/WT vs MSS/MUT | 7·88e-3 | 1·80e-4 | 1·08e-1 | 1·14e-3 |
| | MSS/WT vs MSI/MUT | 8·10e-4 | 3·28e-1 | 0·00e0 | 1·00e-5 |
| | MSI/WT vs MSS/MUT | 0·00e0 | 0·00e0 | 0·00e0 | 0·00e0 |
| | MSI/WT vs MSI/MUT | 3·32e-2 | 1·54e-2 | 9·19e-1 | 7·27e-2 |
| | MSS/MUT vs MSI/MUT | 5·15e-3 | 8·98e-1 | 0·00e0 | 0·00e0 |
| MECOM | MSS/WT vs MSI/WT | 0·00e0 | 0·00e0 | 0·00e0 | 0·00e0 |
| | MSS/WT vs MSS/MUT | 5·54e-2 | 2·53e-1 | 7·16e-1 | 6·78e-1 |
| | MSS/WT vs MSI/MUT | 0·00e0 | 0·00e0 | 0·00e0 | 0·00e0 |
| | MSI/WT vs MSS/MUT | 4·21e-2 | 5·52e-2 | 1·55e-3 | 1·55e-3 |
| | MSI/WT vs MSI/MUT | 9·98e-1 | 3·32e-1 | 1·86e-1 | 1·37e-1 |
| | MSS/MUT vs MSI/MUT | 1·10e-2 | 2·13e-2 | 3·08e-3 | 5·38e-3 |
| NRAS | MSS/WT vs MSI/WT | 0·00e0 | 0·00e0 | 0·00e0 | 0·00e0 |
| | MSS/WT vs MSS/MUT | 2·96e-1 | 8·12e-1 | 5·81e-1 | 2·52e-1 |
| | MSS/WT vs MSI/MUT | 2·65e-1 | 3·06e-2 | 9·31e-1 | 9·53e-1 |
| | MSI/WT vs MSS/MUT | 0·00e0 | 9·14e-3 | 0·00e0 | 0·00e0 |
| | MSI/WT vs MSI/MUT | 9·94e-1 | 1·09e-1 | 1·55e-2 | 2·17e-2 |
| | MSS/MUT vs MSI/MUT | 1·86e-1 | 7·58e-2 | 8·25e-1 | 6·98e-1 |
| PIK3CA | MSS/WT vs MSI/WT | 0·00e0 | 0·00e0 | 0·00e0 | 0·00e0 |
| | MSS/WT vs MSS/MUT | 4·36e-2 | 7·07e-3 | 5·82e-1 | 5·41e-3 |
| | MSS/WT vs MSI/MUT | 0·00e0 | 0·00e0 | 0·00e0 | 0·00e0 |
| | MSI/WT vs MSS/MUT | 0·00e0 | 5·41e-3 | 0·00e0 | 0·00e0 |
| | MSI/WT vs MSI/MUT | 2·21e-1 | 8·16e-1 | 8·92e-3 | 6·45e-3 |
| | MSS/MUT vs MSI/MUT | 0·00e0 | 7·06e-3 | 0·00e0 | 1·00e-5 |
| PLEKHA6 | MSS/WT vs MSI/WT | 0·00e0 | 0·00e0 | 0·00e0 | 0·00e0 |
| | MSS/WT vs MSS/MUT | 2·47e-1 | 8·99e-2 | 7·81e-3 | 7·81e-3 |
| | MSS/WT vs MSI/MUT | 0·00e0 | 0·00e0 | 0·00e0 | 0·00e0 |
| | MSI/WT vs MSS/MUT | 1·81e-3 | 6·66e-2 | 2·63e-1 | 3·16e-1 |
| | MSI/WT vs MSI/MUT | 2·36e-2 | 2·06e-1 | 9·12e-2 | 2·63e-1 |
| | MSS/MUT vs MSI/MUT | 7·70e-4 | 3·74e-2 | 3·45e-1 | 1·38e-1 |
| RFX5 | MSS/WT vs MSI/WT | 0·00e0 | 0·00e0 | 0·00e0 | 0·00e0 |
| | MSS/WT vs MSS/MUT | 3·80e-1 | 5·63e-1 | 3·82e-1 | 2·43e-1 |
| | MSS/WT vs MSI/MUT | 0·00e0 | 0·00e0 | 0·00e0 | 0·00e0 |
| | MSI/WT vs MSS/MUT | 5·12e-1 | 1·98e-1 | 5·19e-3 | 2·60e-3 |
| | MSI/WT vs MSI/MUT | 1·09e-1 | 1·41e-1 | 6·81e-1 | 9·09e-1 |
| | MSS/MUT vs MSI/MUT | 3·57e-1 | 7·14e-2 | 2·56e-2 | 2·56e-2 |
| RNF43 | MSS/WT vs MSI/WT | 0·00e0 | 0·00e0 | 0·00e0 | 0·00e0 |
| | MSS/WT vs MSS/MUT | 1·75e-2 | 3·77e-3 | 4·26e-3 | 1·30e-3 |
| | MSS/WT vs MSI/MUT | 0·00e0 | 0·00e0 | 0·00e0 | 0·00e0 |
| | MSI/WT vs MSS/MUT | 1·44e-3 | 4·93e-2 | 0·00e0 | 0·00e0 |
| | MSI/WT vs MSI/MUT | 9·92e-2 | 1·41e-2 | 5·57e-1 | 5·67e-1 |
| | MSS/MUT vs MSI/MUT | 0·00e0 | 0·00e0 | 0·00e0 | 0·00e0 |
| SMAD2 | MSS/WT vs MSI/WT | 0·00e0 | 0·00e0 | 0·00e0 | 0·00e0 |
| | MSS/WT vs MSS/MUT | 1·74e-2 | 1·70e-1 | 1·72e-3 | 1·21e-3 |
| | MSS/WT vs MSI/MUT | 2·06e-1 | 7·18e-1 | 0·00e0 | 0·00e0 |
| | MSI/WT vs MSS/MUT | 5·60e-4 | 1·73e-1 | 0·00e0 | 1·00e-5 |
| | MSI/WT vs MSI/MUT | 2·80e-4 | 3·46e-2 | 7·17e-1 | 9·44e-1 |
| | MSS/MUT vs MSI/MUT | 6·45e-1 | 5·49e-1 | 4·00e-5 | 2·68e-3 |
| SMG1 | MSS/WT vs MSI/WT | 0·00e0 | 0·00e0 | 0·00e0 | 0·00e0 |
| | MSS/WT vs MSS/MUT | 6·54e-1 | 2·95e-1 | 1·50e-3 | 6·50e-4 |
| | MSS/WT vs MSI/MUT | 5·00e-5 | 6·06e-3 | 0·00e0 | 0·00e0 |
| | MSI/WT vs MSS/MUT | 4·20e-4 | 1·41e-2 | 1·84e-3 | 1·01e-2 |
| | MSI/WT vs MSI/MUT | 8·39e-2 | 3·27e-2 | 5·73e-1 | 8·58e-1 |
| | MSS/MUT vs MSI/MUT | 3·73e-2 | 3·60e-1 | 1·97e-3 | 1·31e-2 |
| TBX3 | MSS/WT vs MSI/WT | 0·00e0 | 0·00e0 | 0·00e0 | 0·00e0 |
| | MSS/WT vs MSS/MUT | 1·48e-2 | 1·13e-1 | 1·92e-1 | 1·52e-1 |
| | MSS/WT vs MSI/MUT | 1·00e-5 | 5·00e-2 | 0·00e0 | 0·00e0 |
| | MSI/WT vs MSS/MUT | 6·65e-3 | 9·12e-2 | 2·00e-5 | 1·00e-5 |
| | MSI/WT vs MSI/MUT | 8·56e-1 | 9·86e-2 | 2·72e-1 | 4·98e-1 |
| | MSS/MUT vs MSI/MUT | 4·92e-2 | 8·87e-1 | 3·00e-5 | 2·00e-5 |

| | | | | | |
|---|---|---|---|---|---|
| TGFBR2 | MSS/WT vs MSI/WT | 0·00e0 | 0·00e0 | 0·00e0 | 0·00e0 |
| | MSS/WT vs MSS/MUT | 6·69e-1 | 2·51e-1 | 9·00e-4 | 3·00e-4 |
| | MSS/WT vs MSI/MUT | 0·00e0 | 0·00e0 | 0·00e0 | 0·00e0 |
| | MSI/WT vs MSS/MUT | 3·87e-3 | 2·53e-1 | 0·00e0 | 5·00e-5 |
| | MSI/WT vs MSI/MUT | 2·96e-1 | 8·99e-1 | 6·54e-1 | 7·13e-1 |
| | MSS/MUT vs MSI/MUT | 2·67e-2 | 4·74e-1 | 2·00e-5 | 1·70e-4 |
| TP53 | MSS/WT vs MSI/WT | 0·00e0 | 2·70e-4 | 0·00e0 | 0·00e0 |
| | MSS/WT vs MSS/MUT | 3·12e-3 | 0·00e0 | 7·00e-5 | 0·00e0 |
| | MSS/WT vs MSI/MUT | 0·00e0 | 3·28e-1 | 0·00e0 | 0·00e0 |
| | MSI/WT vs MSS/MUT | 0·00e0 | 0·00e0 | 0·00e0 | 0·00e0 |
| | MSI/WT vs MSI/MUT | 7·54e-1 | 4·44e-2 | 4·26e-1 | 7·67e-2 |
| | MSS/MUT vs MSI/MUT | 0·00e0 | 1·00e-5 | 0·00e0 | 0·00e0 |
| TRPS1 | MSS/WT vs MSI/WT | 0·00e0 | 0·00e0 | 0·00e0 | 0·00e0 |
| | MSS/WT vs MSS/MUT | 4·41e-1 | 3·69e-2 | 1·00e0 | 6·80e-1 |
| | MSS/WT vs MSI/MUT | 0·00e0 | 1·10e-4 | 0·00e0 | 0·00e0 |
| | MSI/WT vs MSS/MUT | 1·00e-5 | 4·90e-2 | 4·00e-5 | 1·20e-4 |
| | MSI/WT vs MSI/MUT | 6·67e-1 | 5·97e-1 | 3·72e-2 | 6·18e-2 |
| | MSS/MUT vs MSI/MUT | 1·26e-3 | 2·26e-1 | 5·00e-5 | 2·20e-4 |
| WNT16 | MSS/WT vs MSI/WT | 0·00e0 | 0·00e0 | 0·00e0 | 0·00e0 |
| | MSS/WT vs MSS/MUT | 2·77e-1 | 4·55e-1 | 9·38e-2 | 1·25e-1 |
| | MSS/WT vs MSI/MUT | 0·00e0 | 0·00e0 | 0·00e0 | 0·00e0 |
| | MSI/WT vs MSS/MUT | 6·23e-1 | 5·41e-1 | 3·77e-2 | 3·77e-2 |
| | MSI/WT vs MSI/MUT | 5·31e-1 | 8·69e-1 | 5·72e-1 | 7·24e-1 |
| | MSS/MUT vs MSI/MUT | 4·44e-1 | 4·44e-1 | 1·43e-1 | 1·43e-1 |
| ZHX2 | MSS/WT vs MSI/WT | 0·00e0 | 0·00e0 | 0·00e0 | 0·00e0 |
| | MSS/WT vs MSS/MUT | 7·25e-1 | 1·51e-1 | 2·67e-2 | 1·83e-2 |
| | MSS/WT vs MSI/MUT | 2·90e-4 | 4·90e-4 | 0·00e0 | 0·00e0 |
| | MSI/WT vs MSS/MUT | 1·79e-3 | 1·11e-1 | 2·02e-3 | 9·00e-4 |
| | MSI/WT vs MSI/MUT | 1·69e-1 | 6·65e-1 | 8·09e-1 | 7·59e-1 |
| | MSS/MUT vs MSI/MUT | 3·69e-2 | 2·57e-1 | 8·58e-3 | 3·19e-3 |
| ZNRF3 | MSS/WT vs MSI/WT | 0·00e0 | 0·00e0 | 0·00e0 | 0·00e0 |
| | MSS/WT vs MSS/MUT | 9·27e-1 | 3·96e-1 | 9·89e-2 | 1·91e-1 |
| | MSS/WT vs MSI/MUT | 0·00e0 | 0·00e0 | 0·00e0 | 0·00e0 |
| | MSI/WT vs MSS/MUT | 1·50e-3 | 1·30e-4 | 0·00e0 | 0·00e0 |
| | MSI/WT vs MSI/MUT | 1·83e-1 | 3·52e-1 | 1·52e-1 | 1·27e-1 |
| | MSS/MUT vs MSI/MUT | 1·80e-4 | 1·00e-5 | 0·00e0 | 1·00e-5 |

**Tab. S17: Performance metrics of Multi-Target Transformers for the MSI subgroup in external validation.** Mean and standard deviation from the 7 folds of the cross-validation for relevant selected prediction targets in the MSI subgroup. The threshold for binary classification is pre-defined as 0·5· The evaluation metrics include the Matthews Correlation Coefficient (MCC), the Area Under the Receiver Operating Characteristic Curve (AUROC), and the Area Under the Precision-Recall Curve (AUPRC), along with the corresponding mutation rates in external cohorts. Mutation rate contains the ratio of mutations of the respective target within the external data set. The data is sorted for Target and visualized in Fig. S3B.

| Target | Accuracy | Precision | Sensitivity | Specificity | F1 Score | MCC | AUROC | AUPRC | Mutation Rate |
|---|---|---|---|---|---|---|---|---|---|
| ACVR1B | 0·2 (±0·05) | 0·11 (±0·01) | 0·91 (±0·05) | 0·11 (±0·05) | 0·2 (±0·01) | 0·01 (±0·06) | 0·6 (±0·04) | 0·19 (±0·04) | 0·11 |
| AKT1 | 0·2 (±0·07) | 0·11 (±0·0) | 0·91 (±0·08) | 0·12 (±0·08) | 0·2 (±0·01) | 0·03 (±0·04) | 0·55 (±0·04) | 0·15 (±0·03) | 0·11 |
| ALK | 0·25 (±0·02) | 0·22 (±0·01) | 0·92 (±0·08) | 0·05 (±0·05) | 0·36 (±0·02) | -0·06 (±0·06) | 0·44 (±0·03) | 0·24 (±0·05) | 0·23 |
| APC | 0·64 (±0·03) | 0·53 (±0·22) | 0·26 (±0·19) | 0·84 (±0·14) | 0·28 (±0·16) | 0·12 (±0·05) | 0·6 (±0·02) | 0·43 (±0·03) | 0·33 |
| ATM | 0·28 (±0·05) | 0·21 (±0·01) | 0·9 (±0·07) | 0·11 (±0·09) | 0·34 (±0·01) | 0·02 (±0·05) | 0·47 (±0·03) | 0·2 (±0·02) | 0·21 |
| BMPR2 | 0·56 (±0·02) | 0·58 (±0·01) | 0·89 (±0·08) | 0·1 (±0·05) | 0·7 (±0·03) | -0·01 (±0·04) | 0·44 (±0·03) | 0·54 (±0·01) | 0·58 |
| BRAF | 0·57 (±0·02) | 0·56 (±0·01) | 0·91 (±0·07) | 0·17 (±0·07) | 0·7 (±0·02) | 0·12 (±0·04) | 0·49 (±0·02) | 0·55 (±0·02) | 0·54 |
| CCDC40 | 0·2 (±0·09) | 0·1 (±0·01) | 0·81 (±0·21) | 0·13 (±0·13) | 0·18 (±0·03) | -0·04 (±0·06) | 0·46 (±0·04) | 0·13 (±0·04) | 0·11 |
| CDK12 | 0·21 (±0·04) | 0·14 (±0·01) | 0·91 (±0·06) | 0·1 (±0·05) | 0·25 (±0·01) | 0·01 (±0·05) | 0·5 (±0·04) | 0·16 (±0·02) | 0·14 |
| CHD1 | 0·35 (±0·12) | 0·18 (±0·01) | 0·91 (±0·15) | 0·25 (±0·17) | 0·3 (±0·01) | 0·16 (±0·03) | 0·6 (±0·06) | 0·22 (±0·07) | 0·06 |
| CTNND1 | 0·24 (±0·04) | 0·13 (±0·01) | 0·78 (±0·06) | 0·15 (±0·05) | 0·22 (±0·02) | -0·07 (±0·09) | 0·42 (±0·04) | 0·13 (±0·01) | 0·14 |
| DUSP16 | 0·14 (±0·07) | 0·07 (±0·0) | 0·95 (±0·07) | 0·08 (±0·07) | 0·13 (±0·0) | 0·03 (±0·03) | 0·58 (±0·05) | 0·11 (±0·04) | 0·07 |
| ELL2 | 0·22 (±0·1) | 0·05 (±0·0) | 0·95 (±0·13) | 0·18 (±0·11) | 0·1 (±0·0) | 0·08 (±0·03) | 0·65 (±0·06) | 0·1 (±0·02) | 0·02 |
| FHOD3 | 0·52 (±0·04) | 0·48 (±0·02) | 0·84 (±0·13) | 0·25 (±0·15) | 0·61 (±0·04) | 0·13 (±0·08) | 0·54 (±0·03) | 0·49 (±0·03) | 0·16 |
| hypermutated | 0·85 (±0·04) | 0·92 (±0·0) | 0·91 (±0·04) | 0·0 (±0·0) | 0·92 (±0·02) | -0·08 (±0·02) | 0·34 (±0·05) | 0·9 (±0·01) | 0·93 |
| KIF1A | 0·26 (±0·03) | 0·22 (±0·01) | 0·91 (±0·1) | 0·07 (±0·06) | 0·36 (±0·02) | -0·02 (±0·06) | 0·51 (±0·04) | 0·28 (±0·03) | 0·23 |
| KRAS | 0·82 (±0·06) | 0·14 (±0·11) | 0·17 (±0·11) | 0·89 (±0·08) | 0·14 (±0·1) | 0·05 (±0·1) | 0·6 (±0·05) | 0·17 (±0·06) | 0·1 |
| MECOM | 0·39 (±0·04) | 0·34 (±0·02) | 0·78 (±0·09) | 0·17 (±0·07) | 0·47 (±0·04) | -0·06 (±0·12) | 0·42 (±0·04) | 0·32 (±0·03) | 0·13 |
| NRAS | 0·86 (±0·09) | 0·06 (±0·06) | 0·64 (±0·38) | 0·87 (±0·09) | 0·11 (±0·1) | 0·17 (±0·15) | 0·81 (±0·16) | 0·11 (±0·07) | 0·01 |
| PIK3CA | 0·26 (±0·02) | 0·23 (±0·01) | 0·81 (±0·08) | 0·08 (±0·05) | 0·35 (±0·02) | -0·15 (±0·06) | 0·38 (±0·04) | 0·22 (±0·04) | 0·25 |
| PLEKHA6 | 0·49 (±0·04) | 0·46 (±0·02) | 0·9 (±0·08) | 0·19 (±0·07) | 0·61 (±0·03) | 0·13 (±0·11) | 0·57 (±0·05) | 0·47 (±0·04) | 0·16 |
| RFX5 | 0·3 (±0·07) | 0·18 (±0·01) | 0·91 (±0·13) | 0·17 (±0·11) | 0·3 (±0·02) | 0·09 (±0·06) | 0·51 (±0·06) | 0·19 (±0·02) | 0·06 |
| RNF43 | 0·68 (±0·02) | 0·71 (±0·01) | 0·9 (±0·05) | 0·17 (±0·07) | 0·79 (±0·02) | 0·1 (±0·07) | 0·53 (±0·01) | 0·71 (±0·02) | 0·69 |
| SMAD2 | 0·11 (±0·03) | 0·07 (±0·0) | 0·97 (±0·06) | 0·05 (±0·04) | 0·14 (±0·01) | 0·02 (±0·05) | 0·51 (±0·07) | 0·11 (±0·04) | 0·07 |
| SMG1 | 0·22 (±0·09) | 0·11 (±0·01) | 0·87 (±0·16) | 0·14 (±0·12) | 0·2 (±0·02) | 0·03 (±0·08) | 0·48 (±0·06) | 0·12 (±0·02) | 0·11 |

| | | | | | | | | | |
|---|---|---|---|---|---|---|---|---|---|
| TBX3 | 0·21 (±0·05) | 0·16 (±0·0) | 0·96 (±0·07) | 0·07 (±0·07) | 0·28 (±0·0) | 0·06 (±0·02) | 0·53 (±0·05) | 0·2 (±0·03) | 0·16 |
| TGFBR2 | 0·25 (±0·04) | 0·18 (±0·0) | 0·89 (±0·08) | 0·11 (±0·06) | 0·3 (±0·01) | 0·0 (±0·03) | 0·51 (±0·03) | 0·21 (±0·01) | 0·18 |
| TP53 | 0·71 (±0·08) | 0·4 (±0·16) | 0·17 (±0·2) | 0·88 (±0·16) | 0·19 (±0·1) | 0·09 (±0·07) | 0·58 (±0·03) | 0·32 (±0·02) | 0·25 |
| TRPS1 | 0·37 (±0·08) | 0·31 (±0·03) | 0·94 (±0·05) | 0·13 (±0·12) | 0·47 (±0·02) | 0·09 (±0·1) | 0·58 (±0·07) | 0·38 (±0·07) | 0·11 |
| WNT16 | 0·38 (±0·11) | 0·22 (±0·02) | 0·81 (±0·15) | 0·27 (±0·17) | 0·34 (±0·03) | 0·08 (±0·08) | 0·53 (±0·03) | 0·24 (±0·02) | 0·07 |
| ZHX2 | 0·19 (±0·04) | 0·09 (±0·01) | 0·87 (±0·09) | 0·12 (±0·06) | 0·17 (±0·01) | -0·01 (±0·05) | 0·48 (±0·06) | 0·11 (±0·02) | 0·1 |
| ZNRF3 | 0·36 (±0·01) | 0·33 (±0·01) | 0·84 (±0·06) | 0·11 (±0·05) | 0·47 (±0·01) | -0·08 (±0·03) | 0·44 (±0·02) | 0·32 (±0·02) | 0·34 |

**Tab. S18: Performance metrics of Multi-Target Transformers for the MSS subgroup in external validation.** Mean and standard deviation from the 7 folds of the cross-validation for relevant selected prediction targets in the MSS subgroup. The threshold for binary classification is pre-defined as 0·5· The evaluation metrics include the Matthews Correlation Coefficient (MCC), the Area Under the Receiver Operating Characteristic Curve (AUROC), and the Area Under the Precision-Recall Curve (AUPRC), along with the corresponding mutation rates in external cohorts. Mutation rate contains the ratio of mutations of the respective target within the external data set. The data is sorted for Target and visualized in Fig. S3B.

| Target | Accuracy | Precision | Sensitivity | Specificity | F1 Score | MCC | AUROC | AUPRC | Mutation Rate |
|---|---|---|---|---|---|---|---|---|---|
| ACVR1B | 0·77 (±0·1) | 0·11 (±0·03) | 0·64 (±0·21) | 0·77 (±0·11) | 0·19 (±0·05) | 0·2 (±0·06) | 0·77 (±0·05) | 0·21 (±0·08) | 0·04 |
| AKT1 | 0·59 (±0·22) | 0·04 (±0·01) | 0·64 (±0·26) | 0·59 (±0·23) | 0·07 (±0·01) | 0·08 (±0·02) | 0·67 (±0·04) | 0·05 (±0·01) | 0·02 |
| ALK | 0·49 (±0·31) | 0·03 (±0·01) | 0·58 (±0·34) | 0·49 (±0·32) | 0·06 (±0·01) | 0·03 (±0·03) | 0·58 (±0·03) | 0·04 (±0·01) | 0·03 |
| APC | 0·62 (±0·09) | 0·72 (±0·03) | 0·76 (±0·24) | 0·32 (±0·27) | 0·71 (±0·15) | 0·09 (±0·06) | 0·56 (±0·05) | 0·73 (±0·04) | 0·69 |
| ATM | 0·73 (±0·1) | 0·03 (±0·01) | 0·16 (±0·09) | 0·76 (±0·11) | 0·05 (±0·02) | -0·04 (±0·02) | 0·42 (±0·02) | 0·05 (±0·02) | 0·05 |
| BMPR2 | 0·83 (±0·08) | 0·03 (±0·01) | 0·71 (±0·3) | 0·83 (±0·08) | 0·06 (±0·03) | 0·12 (±0·06) | 0·87 (±0·08) | 0·05 (±0·02) | 0·01 |
| BRAF | 0·79 (±0·04) | 0·13 (±0·03) | 0·28 (±0·11) | 0·83 (±0·05) | 0·17 (±0·05) | 0·08 (±0·04) | 0·63 (±0·03) | 0·15 (±0·02) | 0·08 |
| CCDC40 | 0·78 (±0·12) | 0·02 (±0·01) | 0·54 (±0·34) | 0·79 (±0·12) | 0·04 (±0·02) | 0·07 (±0·05) | 0·77 (±0·07) | 0·03 (±0·01) | 0·01 |
| CDK12 | 0·74 (±0·09) | 0·02 (±0·01) | 0·57 (±0·28) | 0·74 (±0·09) | 0·04 (±0·01) | 0·07 (±0·04) | 0·75 (±0·08) | 0·03 (±0·01) | 0·01 |
| CHD1 | 0·87 (±0·06) | 0·0 (±0·01) | 0·14 (±0·38) | 0·87 (±0·06) | 0·01 (±0·02) | 0·0 (±0·06) | 0·71 (±0·15) | 0·02 (±0·01) | 0·0 |
| CTNND1 | 0·83 (±0·08) | 0·02 (±0·01) | 0·43 (±0·16) | 0·83 (±0·08) | 0·03 (±0·01) | 0·06 (±0·02) | 0·63 (±0·11) | 0·04 (±0·03) | 0·01 |
| DUSP16 | 0·57 (±0·23) | 0·01 (±0·01) | 0·46 (±0·34) | 0·58 (±0·23) | 0·02 (±0·01) | 0·01 (±0·05) | 0·55 (±0·11) | 0·01 (±0·0) | 0·01 |
| ELL2 | 0·84 (±0·08) | 0·0 (±0·0) | 0·14 (±0·38) | 0·84 (±0·09) | 0·0 (±0·01) | -0·01 (±0·04) | 0·53 (±0·24) | 0·01 (±0·0) | 0·0 |
| FHOD3 | 0·85 (±0·06) | 0·09 (±0·02) | 0·45 (±0·19) | 0·87 (±0·07) | 0·14 (±0·02) | 0·15 (±0·04) | 0·68 (±0·07) | 0·12 (±0·03) | 0·01 |
| hypermutated | 0·76 (±0·08) | 0·03 (±0·01) | 0·18 (±0·07) | 0·78 (±0·08) | 0·05 (±0·02) | -0·01 (±0·03) | 0·49 (±0·05) | 0·03 (±0·0) | 0·03 |
| KIF1A | 0·54 (±0·26) | 0·07 (±0·01) | 0·6 (±0·23) | 0·53 (±0·28) | 0·12 (±0·02) | 0·07 (±0·03) | 0·58 (±0·04) | 0·07 (±0·01) | 0·05 |
| KRAS | 0·53 (±0·05) | 0·46 (±0·03) | 0·6 (±0·2) | 0·47 (±0·21) | 0·51 (±0·07) | 0·08 (±0·06) | 0·57 (±0·04) | 0·5 (±0·04) | 0·42 |
| MECOM | 0·84 (±0·06) | 0·01 (±0·01) | 0·1 (±0·16) | 0·85 (±0·06) | 0·01 (±0·02) | -0·02 (±0·04) | 0·55 (±0·09) | 0·02 (±0·01) | 0·01 |
| NRAS | 0·48 (±0·17) | 0·05 (±0·01) | 0·48 (±0·21) | 0·48 (±0·19) | 0·09 (±0·02) | -0·02 (±0·03) | 0·47 (±0·05) | 0·06 (±0·02) | 0·06 |
| PIK3CA | 0·57 (±0·13) | 0·16 (±0·02) | 0·55 (±0·2) | 0·58 (±0·18) | 0·25 (±0·04) | 0·09 (±0·05) | 0·59 (±0·03) | 0·16 (±0·01) | 0·13 |
| PLEKHA6 | 0·84 (±0·06) | 0·03 (±0·01) | 1·0 (±0·0) | 0·84 (±0·06) | 0·06 (±0·02) | 0·15 (±0·04) | 0·99 (±0·01) | 0·48 (±0·38) | 0·0 |
| RFX5 | 0·81 (±0·09) | 0·0 (±0·0) | 0·0 (±0·0) | 0·82 (±0·1) | 0·0 (±0·0) | -0·04 (±0·01) | 0·3 (±0·13) | 0·01 (±0·0) | 0·0 |
| RNF43 | 0·81 (±0·06) | 0·07 (±0·01) | 0·31 (±0·12) | 0·83 (±0·06) | 0·12 (±0·02) | 0·07 (±0·03) | 0·69 (±0·02) | 0·08 (±0·01) | 0·04 |
| SMAD2 | 0·52 (±0·21) | 0·07 (±0·02) | 0·69 (±0·15) | 0·51 (±0·23) | 0·12 (±0·03) | 0·09 (±0·04) | 0·68 (±0·03) | 0·13 (±0·05) | 0·04 |
| SMG1 | 0·71 (±0·17) | 0·04 (±0·02) | 0·67 (±0·24) | 0·71 (±0·17) | 0·07 (±0·03) | 0·1 (±0·03) | 0·79 (±0·08) | 0·07 (±0·03) | 0·01 |

| | | | | | | | | | |
|---|---|---|---|---|---|---|---|---|---|
| TBX3 | 0·65 (±0·16) | 0·02 (±0·01) | 0·41 (±0·28) | 0·65 (±0·16) | 0·04 (±0·02) | 0·02 (±0·05) | 0·6 (±0·07) | 0·04 (±0·01) | 0·02 |
| TGFBR2 | 0·79 (±0·09) | 0·07 (±0·01) | 0·6 (±0·21) | 0·79 (±0·09) | 0·13 (±0·02) | 0·15 (±0·04) | 0·76 (±0·05) | 0·08 (±0·01) | 0·03 |
| TP53 | 0·62 (±0·03) | 0·72 (±0·04) | 0·68 (±0·14) | 0·52 (±0·18) | 0·69 (±0·05) | 0·21 (±0·06) | 0·66 (±0·03) | 0·75 (±0·03) | 0·64 |
| TRPS1 | 0·51 (±0·2) | 0·04 (±0·02) | 0·51 (±0·16) | 0·51 (±0·21) | 0·08 (±0·03) | 0·01 (±0·06) | 0·52 (±0·09) | 0·05 (±0·01) | 0·02 |
| WNT16 | 0·87 (±0·08) | 0·0 (±0·0) | 0·0 (±0·0) | 0·88 (±0·08) | 0·0 (±0·0) | -0·02 (±0·01) | 0·12 (±0·11) | 0·0 (±0·0) | 0·0 |
| ZHX2 | 0·76 (±0·06) | 0·02 (±0·01) | 0·57 (±0·29) | 0·76 (±0·06) | 0·05 (±0·02) | 0·08 (±0·05) | 0·76 (±0·05) | 0·03 (±0·01) | 0·01 |
| ZNRF3 | 0·82 (±0·05) | 0·0 (±0·0) | 0·02 (±0·04) | 0·84 (±0·05) | 0·0 (±0·01) | -0·05 (±0·02) | 0·4 (±0·07) | 0·02 (±0·0) | 0·02 |

**Tab. S19: Pathological review of top tiles for selected slides.** The slides correspond to those used for creating the heatmaps. We selected the top and bottom slides based on the prediction score (highest and lowest) for the genetic alteration. All of these tiles were assigned high attention by the model and are therefore considered to be of high relevance for the resulting slide-based prediction score. Sorted by Slide Name. Abbreviations: HM: Hypermutation; MSI: Microsatellite instability; MSS: Microsatellite stability; MUT: Mutated; NOS: Not otherwise specified; TILs: Tumor infiltrating lymphocytes; WT: Wild type

| Slide Name | Alteration | Ground Truth | Prediction Score | Tiles shown | Histopathological Assessment |
|---|---|---|---|---|---|
| CRA, 5661 | MSS/MSI | MSI | 0·99 | top | medullary carcinoma (solid growth, sheets of tumor cells, high number of TILs) |
| CRA, 5661 | TP53 | MUT | 0·18 | bottom | some medullary carcinoma, some connective tissue with prominent vessels, some lymphoid aggregates, some inconspicuous glands |
| CRA, 5694 | MSS/MSI | MSI | 0·06 | bottom | gland-forming adenocarcinoma (NOS) with dirty necrosis |
| CRA, 5694 | TP53 | MUT | 0·69 | top | gland-forming adenocarcinoma (NOS) with dirty necrosis |
| CRA, 5733 | MSS/MSI | MSS | 0·02 | bottom | gland-forming adenocarcinoma (NOS) |
| CRA, 5733 | TP53 | WT | 0·7 | top | gland-forming adenocarcinoma (NOS), connective tissue and smooth muscles |
| WHI, 1031546 | BRAF | WT | 0·96 | top | medullary carcinoma (solid growth, sheets of tumor cells, high number of TILs) |
| WHI, 1031546 | MSS/MSI | MSI | 0·98 | top | medullary carcinoma (solid growth, sheets of tumor cells, high number of TILs) |
| WHI, 1031550 | KRAS | WT | 0·09 | bottom | medullary and mucinous differentiation |
| WHI, 1031550 | MSS/MSI | MSI | 0·99 | top | medullary carcinoma (solid growth, sheets of tumor cells, high number of TILs) |
| WHI, 1031553 | HM | WT | 0·89 | top | mucinous differentiation |
| WHI, 1031553 | MSS/MSI | MSI | 0·91 | top | mucinous differentiation |
| WHI, 1031557 | BRAF | WT | 0·95 | top | medullary and mucinous differentiation |
| WHI, 1031557 | HM | WT | 0·95 | top | medullary and mucinous differentiation |
| WHI, 1031557 | MSS/MSI | MSI | 0·99 | top | mucinous and more solid differentiation, high number of TILs |
| WHI, 1031567 | HM | WT | 0·75 | top | medullary and mucinous differentiation, TILs, connective tissue with prominent vessels |
| WHI, 1031567 | MSS/MSI | MSI | 0·88 | top | medullary and mucinous differentiation, TILs |
| WHI, 1031576 | HM | WT | 0·63 | top | lymphoid aggregates, normal mucosa, adipocytes |
| WHI, 1031576 | MSS/MSI | MSI | 0·85 | top | lymphoid aggregates, normal mucosa, adipocytes |
| WHI, 1031609 | HM | MUT | 0·98 | top | medullary carcinoma (solid growth, sheets of tumor cells, high number of TILs) |
| WHI, 1031609 | MSS/MSI | MSI | 0·99 | top | medullary carcinoma (solid growth, sheets of tumor cells, high number of TILs) |
| WHI, 1031622 | TP53 | WT | 0·11 | bottom | mucinous differentiation |
| WHI, | MSS/MSI | MSI | 0·98 | top | mucinous differentiation |

| Slide Name | Alteration | Ground Truth | Prediction Score | Tiles shown | Histopathological Assessment |
|---|---|---|---|---|---|
| 1031622 | | | | | |
| WHI, 1031632 | MSS/MSI | MSI | 0·98 | top | mucinous and medullary differentiation |
| WHI, 1031632 | RNF43 | WT | 0·93 | top | mucinous and medullary differentiation |
| WHI, 1031662 | APC | MUT | 0·17 | bottom | adipocytes, mucin (without relevant tumor content), lymphoid aggregates |
| WHI, 1031662 | MSS/MSI | MSS | 0·93 | top | mucinous differentiation, TILs |
| WHI, 1031666 | APC | MUT | 0·35 | bottom | mucinous differentiation, lymphoid aggregates, connective tissue |
| WHI, 1031666 | MSS/MSI | MSS | 0·96 | top | partly mucinous differentiation, TILs, also conventional gland-forming adenocarcinoma and few normal glands |
| WHI, 1031666 | RNF43 | WT | 0·91 | top | mucinous differentiation, normal glands with lymphocytic background, connective tissue |
| WHI, 1031672 | KRAS | MUT | 0·82 | top | dysplastic epithelium, superficial tumor parts |
| WHI, 1031672 | MSS/MSI | MSS | 0·27 | bottom | dysplastic epithelium |
| WHI, 1031705 | BRAF | WT | 0·06 | bottom | dysplastic epithelium, superficial tumor parts |
| WHI, 1031705 | MSS/MSI | MSS | 0·04 | bottom | dysplastic epithelium, superficial tumor parts |
| WHI, 1031733 | BRAF | MUT | 0·97 | top | medullary carcinoma (solid growth, sheets of tumor cells, high number of TILs) |
| WHI, 1031733 | MSS/MSI | MSI | 0·97 | top | medullary carcinoma (solid growth, sheets of tumor cells, high number of TILs) |
| WHI, 1031786 | KRAS | MUT | 0·14 | bottom | lymphoid aggregates, few tumor glands, few normal glands, connective tissue, adipocytes |
| WHI, 1031786 | MSS/MSI | MSI | 0·95 | top | medullary carcinoma (solid growth, sheets of tumor cells, high number of TILs), mucus |
| WHI, 1031792 | KRAS | WT | 0·08 | bottom | sheets of tumor cells, TILs |
| WHI, 1031792 | MSS/MSI | MSI | 0·98 | top | sheets of tumor cells, TILs |
| WHI, 1031829 | KRAS | WT | 0·75 | top | dysplastic epithelium |
| WHI, 1031829 | MSS/MSI | MSS | 0·05 | bottom | dysplastic epithelium |
| WHI, 1031846 | KRAS | WT | 0·65 | top | gland-forming adenocarcinoma (NOS) |
| WHI, 1031846 | MSS/MSI | MSI | 0·55 | top | mucinous differentiation |
| WHI, 1031849 | BRAF | MUT | 0·09 | bottom | dysplastic epithelium, tumor glands with dirty necrosis (adenocarcinoma NOS) |
| WHI, 1031849 | MSS/MSI | MSS | 0·08 | bottom | dysplastic epithelium, tumor glands |

**Tab. S20: Shapiro-Wilk test for normal distribution of mean prediction scores presented in Fig. 4.** Abbreviations: MSI: Microsatellite instability; MSS: Microsatellite stability; MUT: Mutated; WT: Wild type.

| Target | WT/MUT | MSS/MSI | Score Distribution | Statistic | p-value |
|---|---|---|---|---|---|
| hypermutated | WT | MSS | MSI | 0·78 | <0·0001 |
| hypermutated | WT | MSS | hypermutated | 0·87 | <0·0001 |
| hypermutated | WT | MSI | MSI | 0·88 | 0·0741 |
| hypermutated | WT | MSI | hypermutated | 0·93 | 0·3123 |
| hypermutated | MUT | MSS | MSI | 0·81 | 0·0071 |
| hypermutated | MUT | MSS | hypermutated | 0·84 | 0·0184 |
| hypermutated | MUT | MSI | MSI | 0·78 | <0·0001 |
| hypermutated | MUT | MSI | hypermutated | 0·77 | <0·0001 |
| BRAF | WT | MSS | MSI | 0·77 | <0·0001 |
| BRAF | WT | MSS | BRAF | 0·83 | <0·0001 |
| BRAF | WT | MSI | MSI | 0·74 | <0·0001 |
| BRAF | WT | MSI | BRAF | 0·78 | <0·0001 |
| BRAF | MUT | MSS | MSI | 0·87 | 0·0004 |
| BRAF | MUT | MSS | BRAF | 0·93 | 0·0126 |
| BRAF | MUT | MSI | MSI | 0·82 | <0·0001 |
| BRAF | MUT | MSI | BRAF | 0·89 | <0·0001 |
| TP53 | WT | MSS | MSI | 0·83 | <0·0001 |
| TP53 | WT | MSS | TP53 | 0·98 | 0·0037 |
| TP53 | WT | MSI | MSI | 0·78 | <0·0001 |
| TP53 | WT | MSI | TP53 | 0·93 | <0·0001 |
| TP53 | MUT | MSS | MSI | 0·76 | <0·0001 |
| TP53 | MUT | MSS | TP53 | 0·98 | <0·0001 |
| TP53 | MUT | MSI | MSI | 0·71 | <0·0001 |
| TP53 | MUT | MSI | TP53 | 0·92 | 0·0064 |
| APC | WT | MSS | MSI | 0·81 | <0·0001 |
| APC | WT | MSS | APC | 0·95 | <0·0001 |
| APC | WT | MSI | MSI | 0·75 | <0·0001 |
| APC | WT | MSI | APC | 0·94 | <0·0001 |
| APC | MUT | MSS | MSI | 0·77 | <0·0001 |
| APC | MUT | MSS | APC | 0·94 | <0·0001 |
| APC | MUT | MSI | MSI | 0·78 | <0·0001 |
| APC | MUT | MSI | APC | 0·95 | 0·0102 |
| BMPR2 | WT | MSS | MSI | 0·78 | <0·0001 |
| BMPR2 | WT | MSS | BMPR2 | 0·83 | <0·0001 |
| BMPR2 | WT | MSI | MSI | 0·71 | <0·0001 |
| BMPR2 | WT | MSI | BMPR2 | 0·72 | <0·0001 |
| BMPR2 | MUT | MSS | MSI | Insufficient data | Insufficient data |
| BMPR2 | MUT | MSS | BMPR2 | Insufficient data | Insufficient data |
| BMPR2 | MUT | MSI | MSI | 0·80 | <0·0001 |
| BMPR2 | MUT | MSI | BMPR2 | 0·83 | <0·0001 |
| RNF43 | WT | MSS | MSI | 0·77 | <0·0001 |
| RNF43 | WT | MSS | RNF43 | 0·81 | <0·0001 |
| RNF43 | WT | MSI | MSI | 0·78 | <0·0001 |
| RNF43 | WT | MSI | RNF43 | 0·84 | <0·0001 |
| RNF43 | MUT | MSS | MSI | 0·92 | 0·1023 |
| RNF43 | MUT | MSS | RNF43 | 0·94 | 0·2874 |
| RNF43 | MUT | MSI | MSI | 0·76 | <0·0001 |
| RNF43 | MUT | MSI | RNF43 | 0·82 | <0·0001 |
| ZNRF3 | WT | MSS | MSI | 0·79 | <0·0001 |
| ZNRF3 | WT | MSS | ZNRF43 | 0·85 | <0·0001 |
| ZNRF3 | WT | MSI | MSI | 0·76 | <0·0001 |
| ZNRF3 | WT | MSI | ZNRF43 | 0·86 | <0·0001 |
| ZNRF3 | MUT | MSS | MSI | 0·87 | 0·129 |
| ZNRF3 | MUT | MSS | ZNRF43 | 0·95 | 0·6615 |
| ZNRF3 | MUT | MSI | MSI | 0·78 | <0·0001 |
| ZNRF3 | MUT | MSI | ZNRF43 | 0·85 | <0·0001 |

| | | | | | |
|---|---|---|---|---|---|
| KRAS | WT | MSS | MSI | 0·76 | <0·0001 |
| KRAS | WT | MSS | KRAS | 0·98 | 0·0031 |
| KRAS | WT | MSI | MSI | 0·76 | <0·0001 |
| KRAS | WT | MSI | KRAS | 0·95 | <0·0001 |
| KRAS | MUT | MSS | MSI | 0·81 | <0·0001 |
| KRAS | MUT | MSS | KRAS | 0·99 | 0·0422 |
| KRAS | MUT | MSI | MSI | 0·80 | 0·0022 |
| KRAS | MUT | MSI | KRAS | 0·98 | 0·95 |

### Supplementary Figures

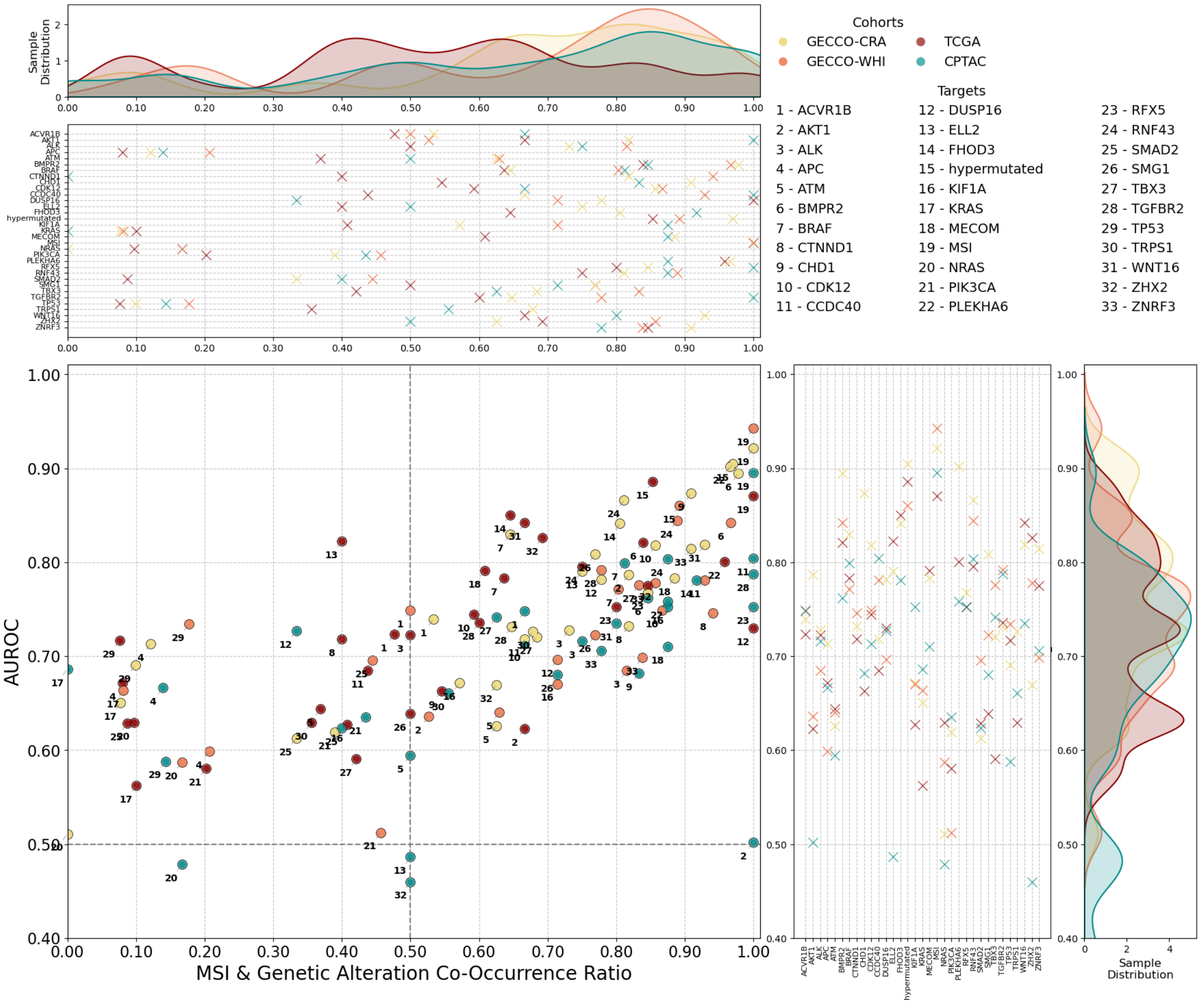

Fig. S1: **Performance evaluation of the primary multi-target transformer on all four external cohorts.** The relationship between the MSI & Genetic Alteration Co-Occurrence Ratio, representing the fraction of cases harboring MSI among those with a specific mutation, and the mean AUROCs from seven folds, reflecting predictive performance, is shown. Each dot corresponds to a prediction target, with IDs mapped to genetic alterations in the legend. Density plots on the top and right illustrate the distribution of co-occurrence ratios and AUROC values between cohorts, respectively. The horizontal and vertical cross distributions, aligned with the target names on the axes, improve clarity of target positions. Further metrics provided in Tab. S5-S8.

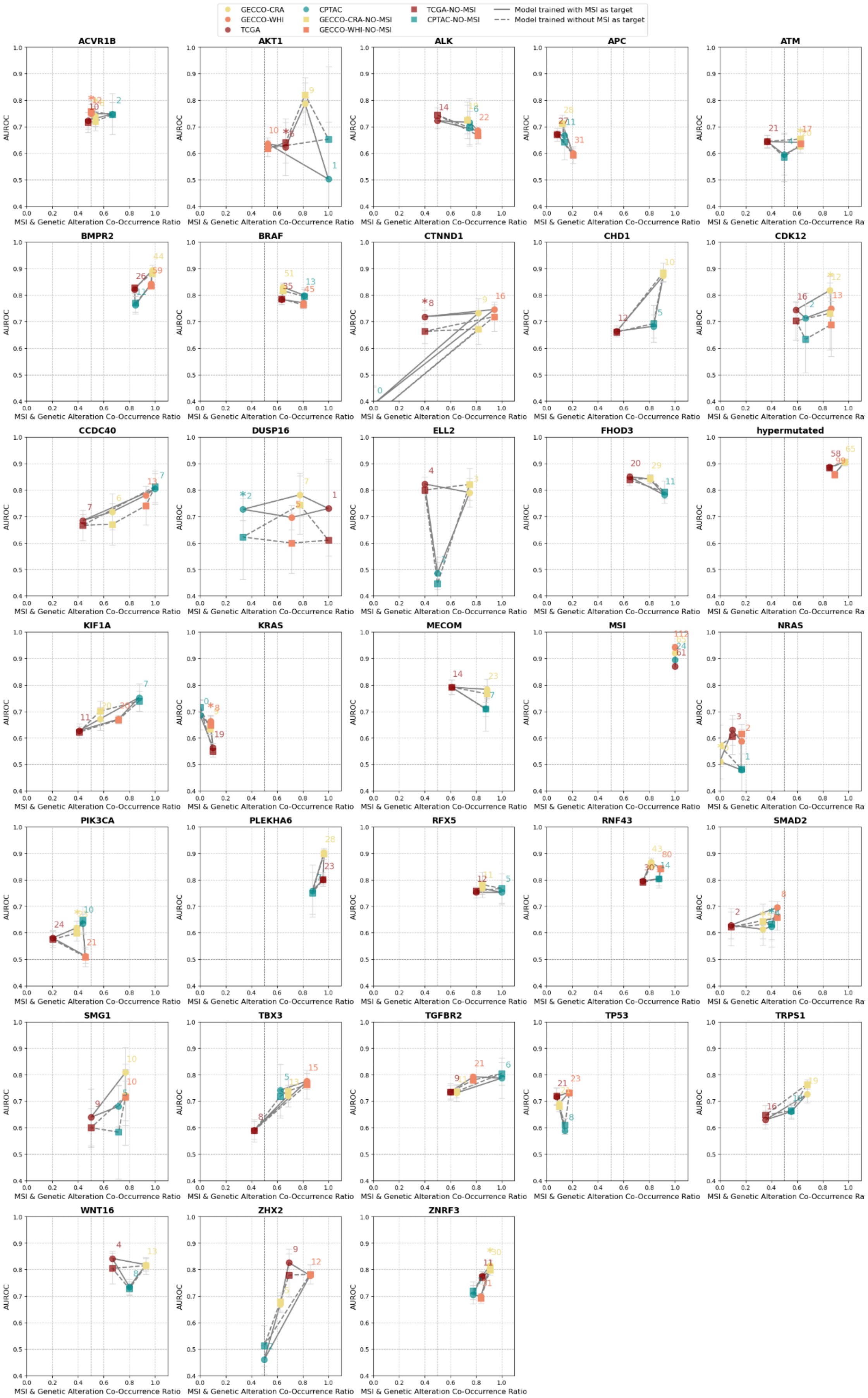

Fig. S2: **Performance comparison of the primary multi-target transformer including MSI as a target with the secondary multi-target transformer excluding MSI as a target on all four external cohorts**. The association between MSI and Genetic Alteration Co-Occurrence Ratio—defined as the proportion of cases exhibiting MSI among those with a given mutation—and the mean AUROCs with standard deviations across seven folds, are presented on a target basis. Each cohort is represented by a distinct color. Dots with continuous lines denote results for the primary model, while squares with dashed lines correspond to the secondary model. Absolute case numbers with concurrent MSI and target mutations are provided for context. Statistical significance, determined via the DeLong test ($p<0{\cdot}05$), is indicated by asterisks. Further metrics provided in Tab. S5-S14.

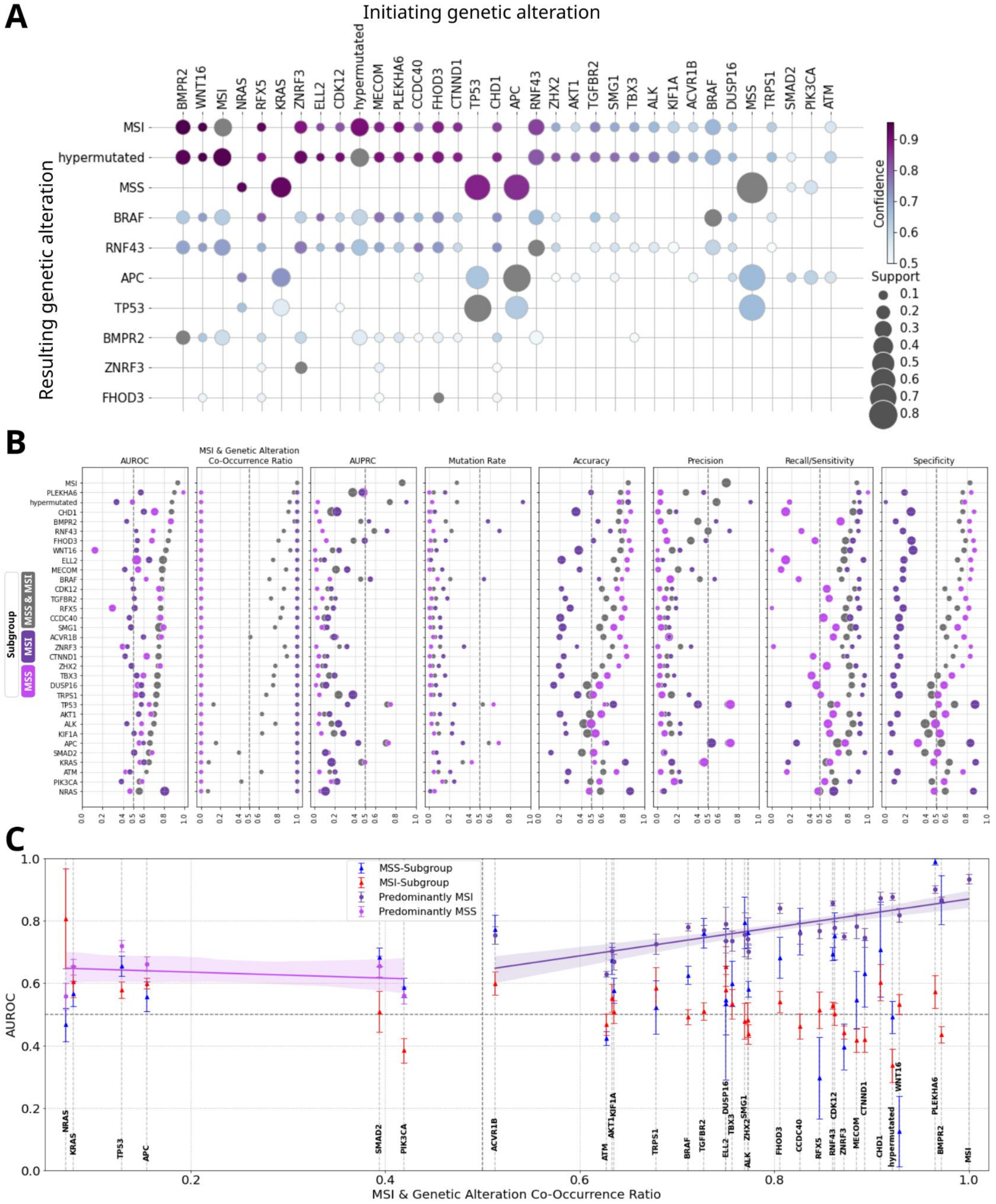

Fig. S3: **Co-Occurrence of genetic alterations and the performance of Multi-Target Transformers on selected alterations. A**. Association rule mining was used to investigate genetic alterations (Tab. S13). The analysis includes an initiating genetic alteration (‘antecedent’) and resulting genetic alteration (‘consequent’). The presence of an initiating alteration statistically increases the probability of observing a resulting alteration, without implying a cause-effect biological sequence. Confidence, calculated as the ratio of co-occurrences to instances of the antecedent alone, indicates the predictive power of the initiating alteration for the resulting alteration. Support measures the frequency of an alteration or combination thereof in the dataset. Gray circles represent the antecedent support, with a support value of 1 indicating that every sample in the dataset has the respective genetic alteration. Microsatellite stability (MSS) and MSI were included to demonstrate relationships with targets commonly associated with MSS. **B.**

Performance metrics of Multi-Target Transformers for external validation. The mean and standard deviation for relevant selected prediction targets for the whole external set, as well as the MSI and MSS subgroups, are displayed based on the 7 folds of cross-validation. The threshold for binary classification is pre-defined as 0·50· The evaluation metrics include the Area Under the Receiver Operating Characteristic Curve (AUROC), and the Area Under the Precision-Recall Curve (AUPRC), along with the corresponding mutation rates in external cohorts. The Mutation Rate refers to the fraction of instances with a specific mutation in the subgroup. The MSI & Genetic Alteration Co-Occurrence Ratio is the fraction of cases harboring MSI among all cases with a particular genetic mutation. The data is sorted for AUROC and shown in Tab. S14 and Tab. S17-S18. **C.** Distribution of AUROCs (mean ± standard deviation) for selected prediction targets and their co-occurrence with MSI in external validation. For comparability, the MSS/MSI subgroup-specific AUROCs are reported for the MSI & Genetic Alteration Co-Occurrence Ratio of the respective alteration in the entire external dataset. The corresponding values and further metrics shown in Tab. S14 and Tab. S17-S18.

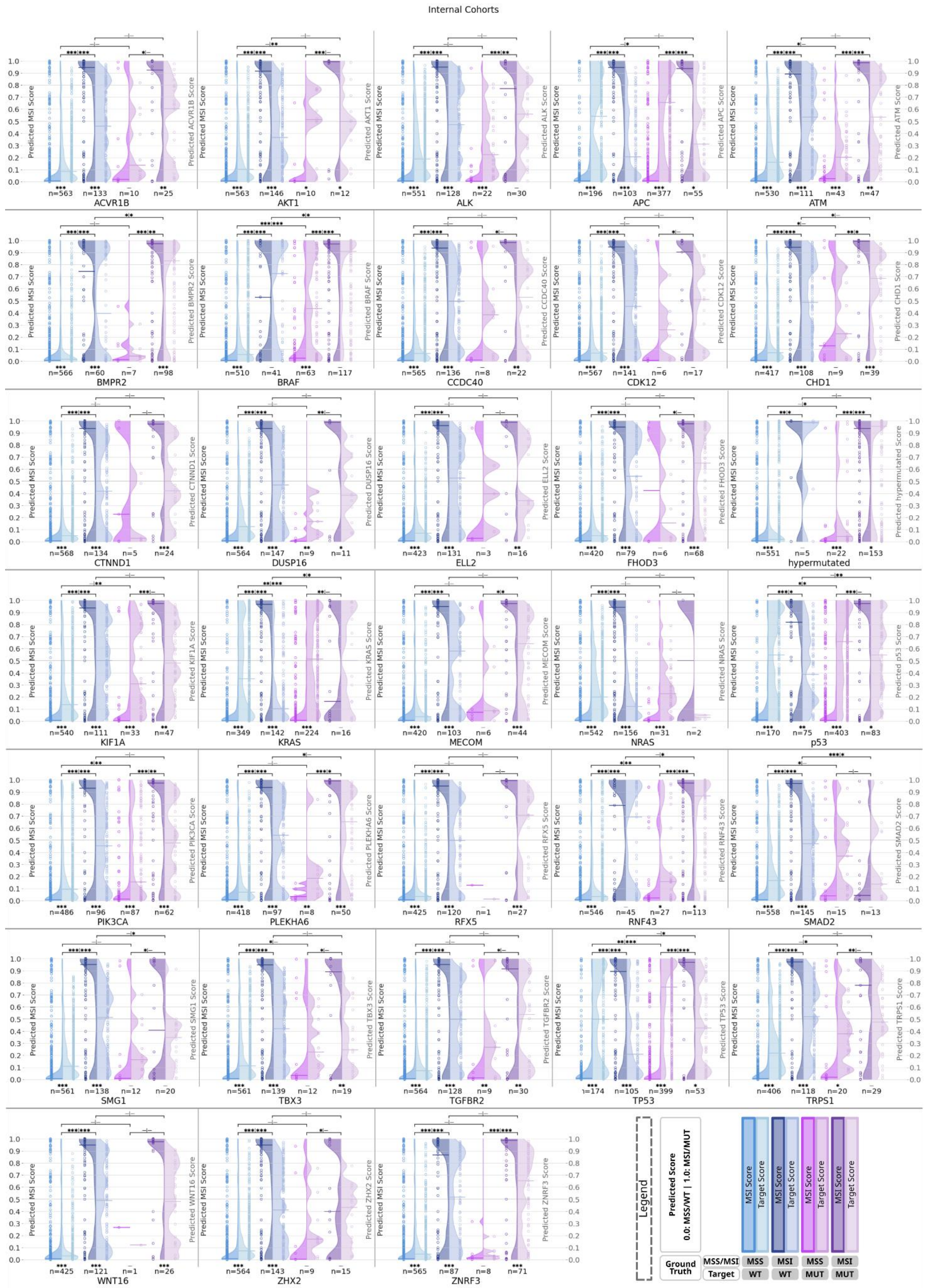

Fig. S4: **Violin plots representing individual patient scores from the train set cohorts for MSI and respective genetic alterations in four subgroups based on microsatellite and alteration mutational status.** The left y-axis represents the MSI score scale (left violins) and the right y-axis corresponds to the prediction target scores (right violins). Representative genetic alterations from genetic Cluster 1-3 were selected, as per Fig. 2. The data encompasses the train set cohorts

(EPIC, CORSA, IWHS) (Fig. 1C). Each dot represents the mean value of individual patient prediction scores calculated from 7 folds, with the horizontal line within each half violin indicating the median of all individual mean patient scores. A horizontal line at 0·50 denotes the line of model uncertainty. The sample count for each subgroup is indicated below the violins. Statistical significance is denoted in the figures as follows: * for $p < 0{\cdot}05$, ** for $p < 0{\cdot}01$, *** for $p < 0{\cdot}001$, with more details provided in Fig. 1D. The Mann-Whitney U test was used for within-group comparisons, and the Wilcoxon test was used for between-group comparisons.

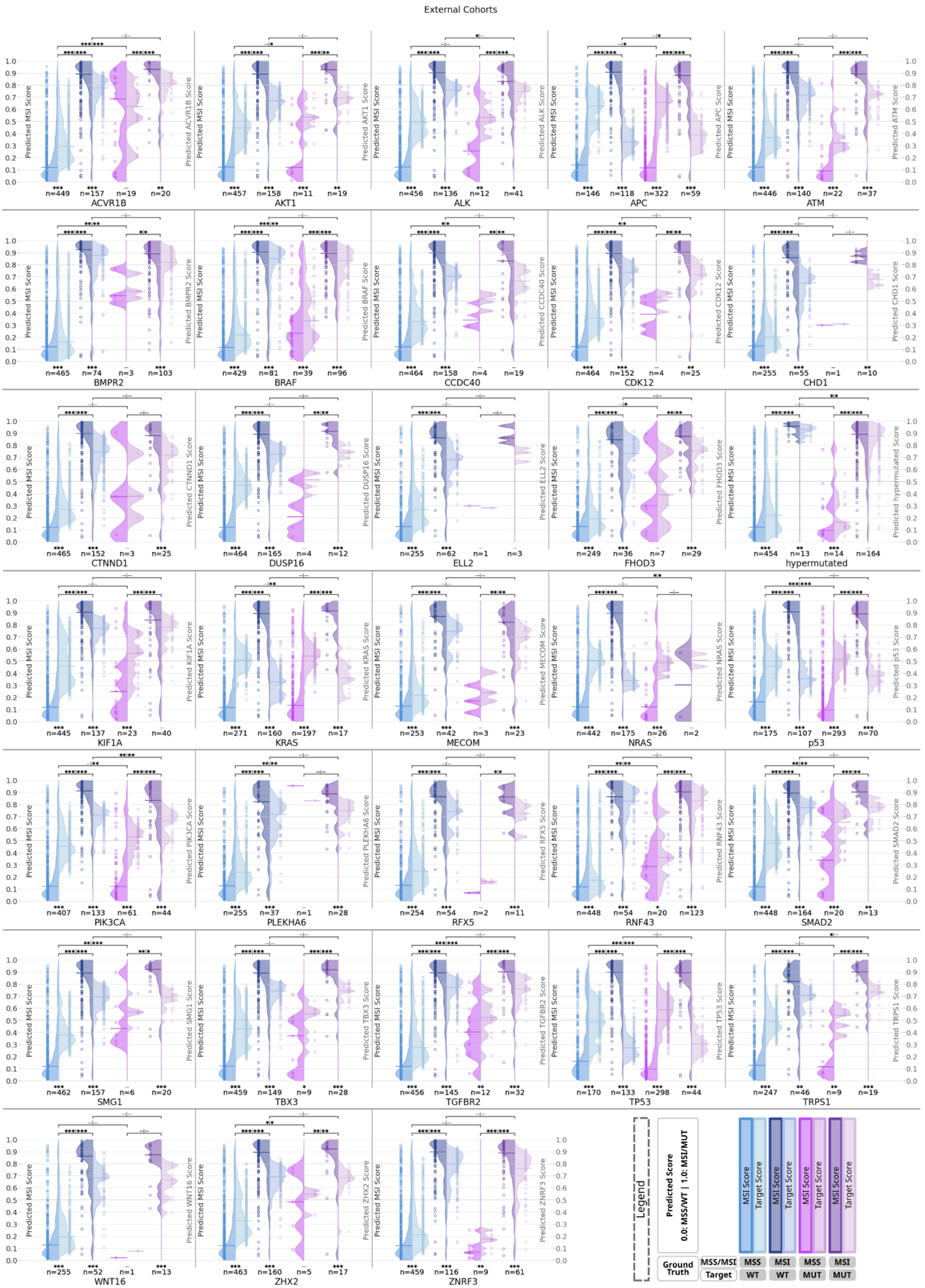

Fig. S5: **Violin plots representing individual patient scores from the test set cohorts for MSI and respective genetic alterations in four subgroups based on microsatellite and alteration mutational status.** The left y-axis represents the MSI score scale (left violins) and the right y-axis corresponds to the prediction target scores (right violins). Representative genetic alterations from genetic Cluster 1-3 were selected, as per Fig. 2. The data encompasses the test set cohorts (CRA,

WHI) (Fig. 1C). Each dot represents the mean value of individual patient prediction scores calculated from 7 folds, with the horizontal line within each half violin indicating the median of all individual mean patient scores. A horizontal line at 0·50 denotes the line of model uncertainty. The sample count for each subgroup is indicated below the violins. Statistical significance is denoted in the figures as follows: * for $p < 0.05$, ** for $p < 0.01$, *** for $p < 0.001$, with more details provided in Fig. 1D. The Mann-Whitney U test was used for within-group comparisons, and the Wilcoxon test was used for between-group comparisons.

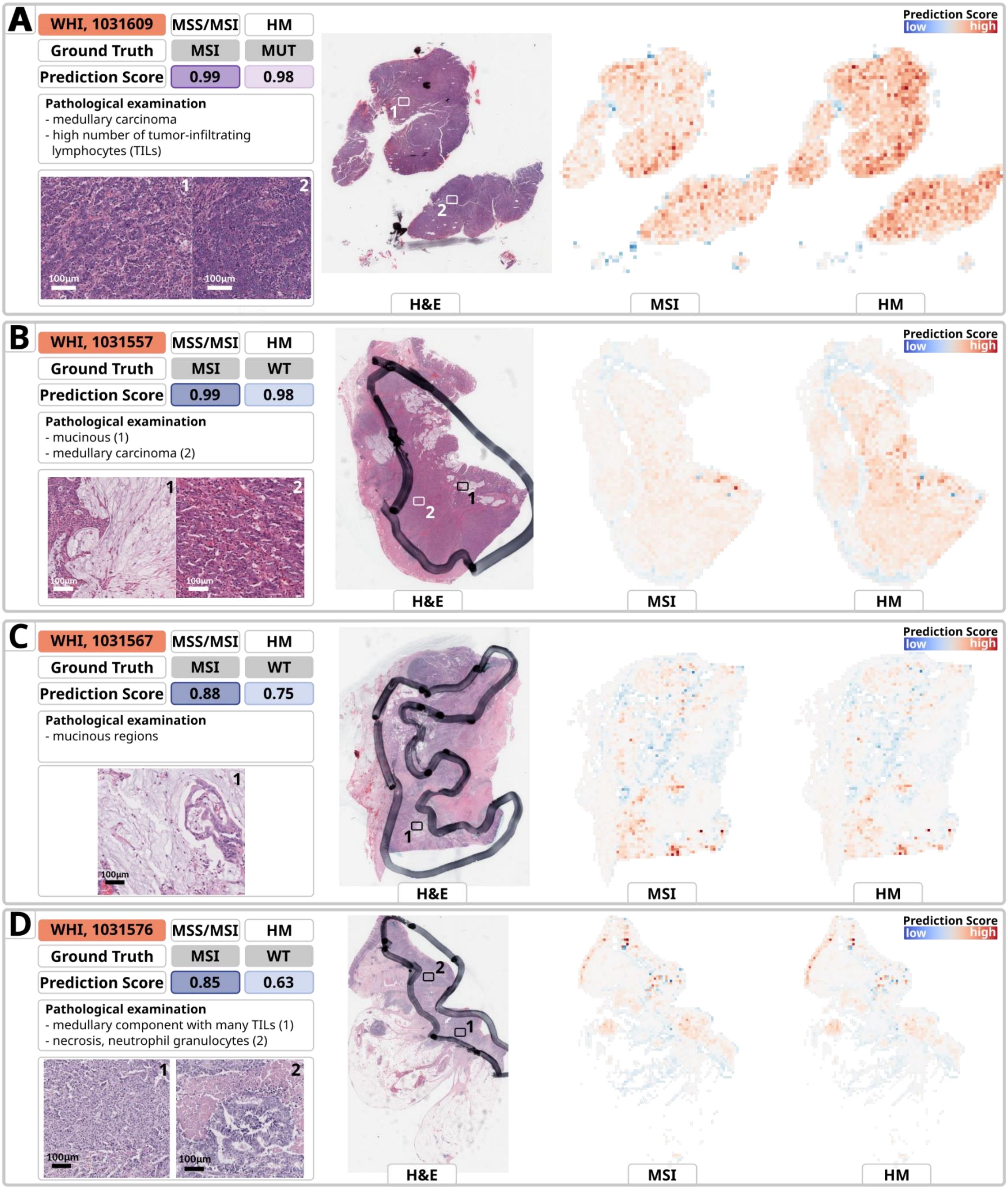

Fig. S6: **Heatmaps of representative samples for prediction of MSI and hypermutation (HM) from the external validation dataset**. The heatmaps are derived from the model with the median AUROC for MSI detection and the majority of prediction targets evaluated by sevenfold cross-validation. The cohort, Sample-ID, ground truth and prediction scores for MSI, along with the HM status, a brief pathological evaluation and magnified views of specific areas are provided for in-depth analysis. The heatmaps show critical tumor areas for predicting MSI (middle) and HM (right). Red signifies high importance and indicates MSI or HM (MUT), while blue signifies low importance and indicates MSS and HM wild type (WT). The color intensity represents the model's attention to that particular area. The sections highlighted in the heatmaps represent tumor tissue and show a high degree of similarity for MSI and HM, indicating that most of the relevant information for both prediction targets is concentrated in the same areas leading to a similar score. Minor variations in the highlighted areas of the heatmaps do not reveal noteworthy differences in pathological assessment, as the tissue in these areas shows similar morphological patterns. **A.** This relatively homogeneous tumor shows sheets of tumor cells and a high number of tumor-infiltrating lymphocytes (TILs), consistent with the diagnosis of a medullary carcinoma. This observation is consistent with the correct prediction of MSI and HM mutations, as these are commonly seen in this type of cancer. **B.-D.** Cases of MSI and wild-type HM displaying elevated scores of both MSI and HM. **B.** MSI-like morphology with mucinous (zoom 1) and solid medullary parts (zoom 2). **C.** Tumor with a mucinous component is shown where the MSI prediction is correct and plausible but the HM WT is predicted with a high score. **D.** Higher degree of intratumoral heterogeneity, which comprises different distinct components, including a medullary component with numerous TILs (zoom 1) and gland-forming areas, necrosis, and neutrophil granulocytes (zoom 2).

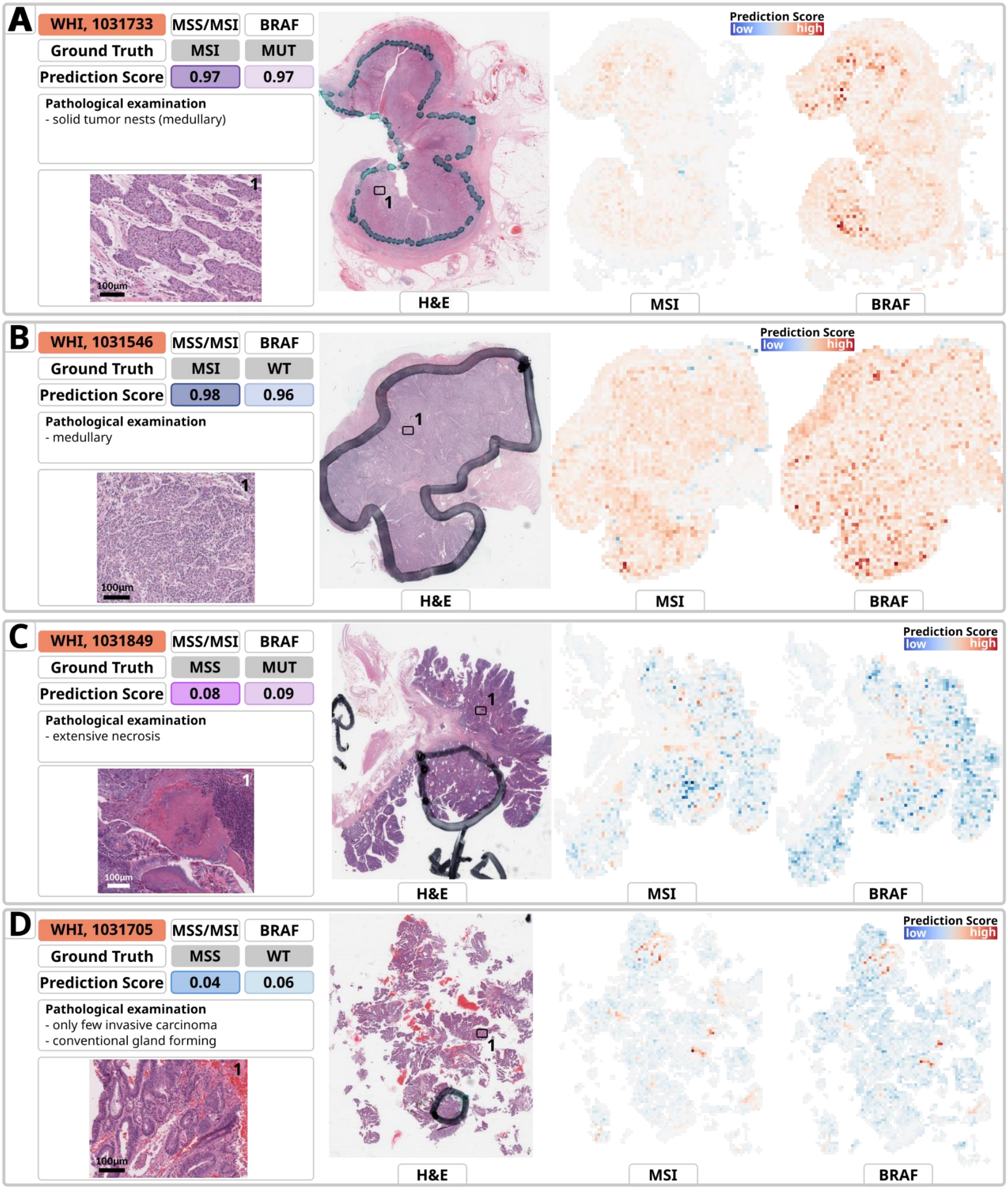

Fig. S7: **Heatmaps of representative samples for prediction of MSI and *BRAF* from the external validation dataset**. The heatmaps are derived from the model with the median AUROC for MSI detection and the majority of prediction targets evaluated by sevenfold cross-validation. The cohort, Sample-ID, ground truth and prediction scores for MSI, along with the *BRAF* mutational status, a brief pathological evaluation and magnified views of specific areas are provided for in-depth analysis. The heatmaps highlight relevant tumor areas for MSI (middle) and *BRAF* mutation (right) prediction. Red signifies high importance and indicates MSI or *BRAF* mutation (MUT), while blue signifies low importance and indicates MSS and *BRAF* wild type (WT). The color intensity represents the model's attention to that particular area. The sections highlighted in the heatmaps represent tumor tissue and show a high degree of similarity for MSI and *BRAF*, indicating that most of the relevant information for both prediction targets is concentrated in the same areas leading to a similar score. Slight deviations in the heatmaps do not indicate any significant variations in a pathological evaluation. **A**. The solid tumor nests are typical of a medullary carcinoma. This observation is consistent with the correct prediction of MSI and *BRAF* mutations, as these are commonly seen in this type of cancer. **B**. The typical morphology indicative of medullary microsatellite insatiable tumor is apparent, providing further support for the prediction of MSI. However, despite a high MSI prediction score, a high score for *BRAF* MUT was obtained even though ground truth is *BRAF* WT. **C**. The extensive necrosis and MSS-like morphology in a MSS, *BRAF* MUT case is potentially responsible for the low prediction score for *BRAF* indicating *BRAF* WT. **D**. The heatmap indicates the existence of numerous superficial components (adenoma) and only a small number of invasive carcinoma components. It is worth noting that the invasive carcinoma features a typical gland-forming morphology resembling that of MSS, which plausibly leads to low MSI and low *BRAF* scores, correctly indicating MSS but falsely indicating *BRAF* WT.

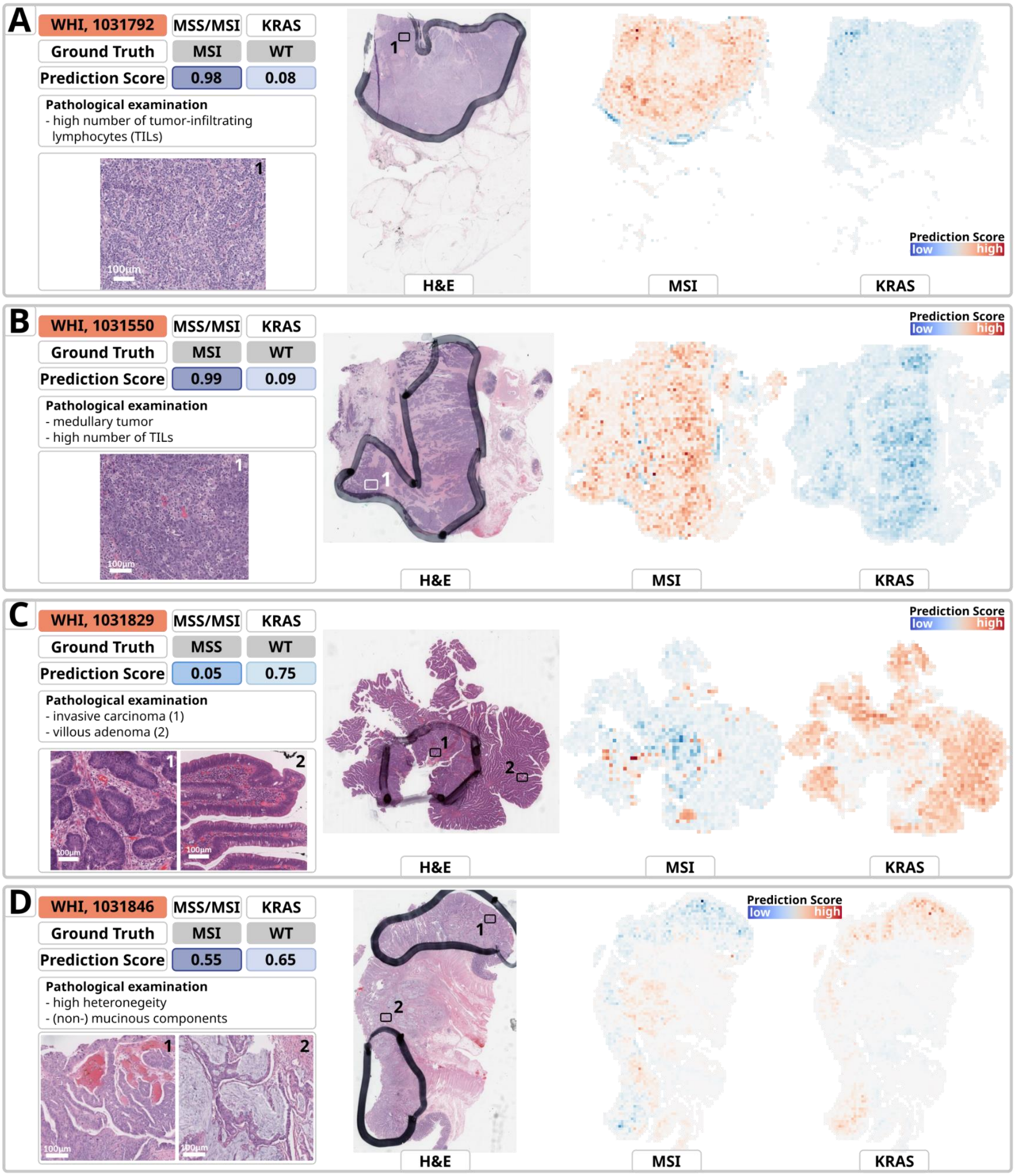

Fig. S8: **Heatmaps of representative samples for prediction of MSI and *KRAS* from the external validation dataset**. The heatmaps are derived from the model with the median AUROC for MSI detection and the majority of prediction targets evaluated by sevenfold cross-validation. The cohort, Sample-ID, ground truth and prediction scores for MSI, along with the *KRAS* mutational status, a brief pathological evaluation and magnified views of specific areas are provided for in-depth analysis. The heatmaps highlight relevant tumor areas for MSI (middle) and *KRAS* mutation (right) prediction. Red signifies high importance and indicates MSI or *KRAS* mutation (MUT), while blue signifies low importance and indicates MSS and *KRAS* wild type (WT). The color intensity represents the model's attention to that particular area. The heatmaps highlight sections of tumor tissue highly similar for MSI and *KRAS*. Nevertheless, corresponding regions in the heatmaps result in divergent scores for MSI (red) and *KRAS* (blue). In certain cases, different regions seem to be crucial for the forecasts of MSI and *KRAS*, respectively. **A.-B.** The tumor has a solid medullary morphology with high number intratumoral lymphocytes, which is quite typical for MSI. The prediction for MSI appears to be driven by the presence of tumor sheets and a high number of TILs, and KRAS is correctly predicted with a low score indicating *KRAS* WT. Identical tumor areas are taken into consideration for predicting each target. **C.** The extensive villous adenoma frequently contains *KRAS* mutations. It is noteworthy that the *KRAS* prediction heatmap focuses on the adenoma parts rather than the invasive carcinoma, which seems more relevant for MSS prediction. While the tissue exhibits MSS-like morphology, resulting in a correspondingly low MSI score, the model yields a relatively high *KRAS* score, falsely indicating *KRAS* MUT. **D.** For predicting MSI, the mucinous component is highlighted whereas for predicting *KRAS*, the superficial tumor parts (without mucin) are highlighted. Due to the heterogeneous nature of the tumor, the predictions seem difficult, resulting in intermediate MSI and *KRAS* scores, indicating model uncertainty.

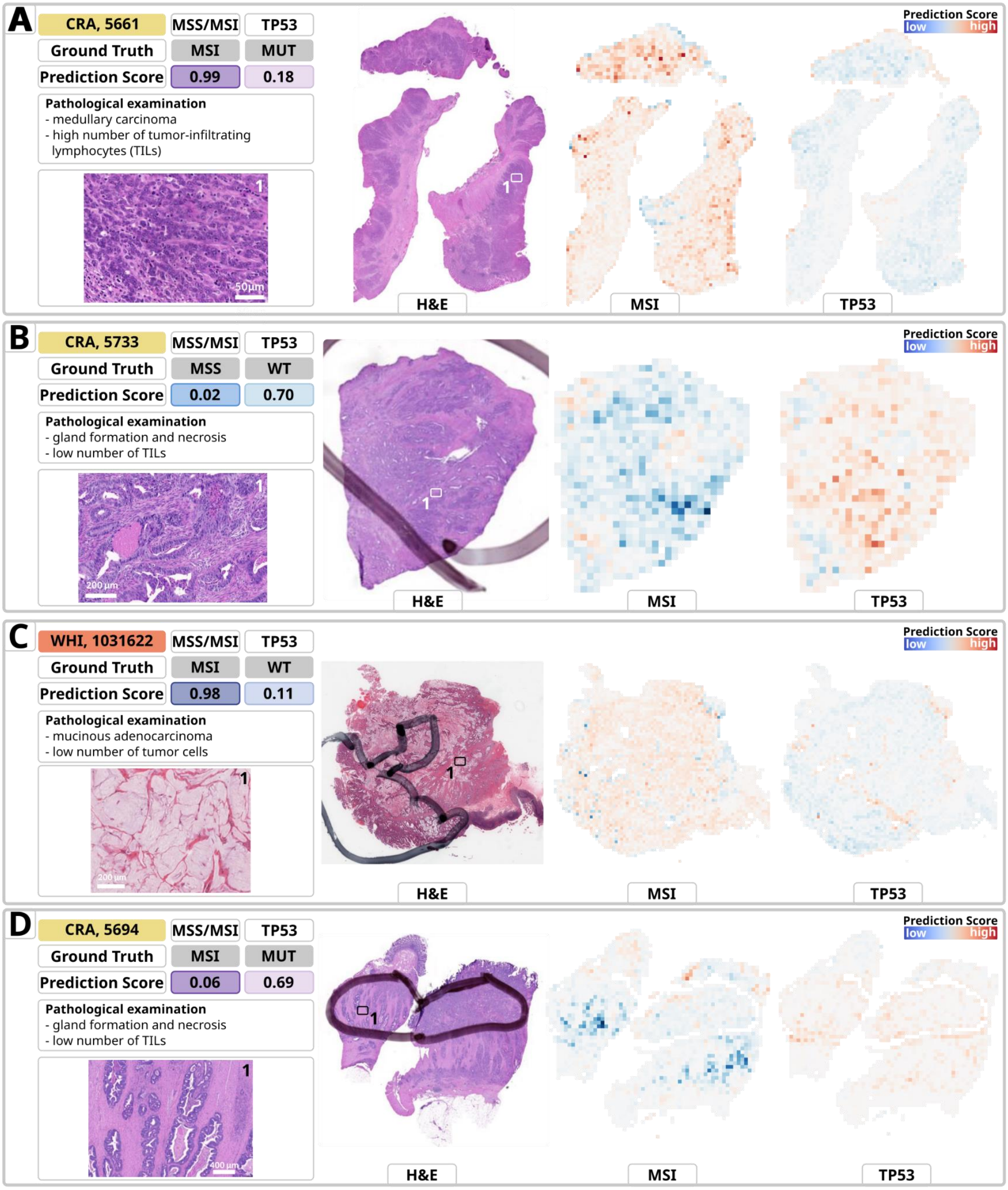

Fig. S9: **Heatmaps of representative samples for prediction of MSI and *TP53* from the external validation dataset**. The heatmaps are derived from the model with the median AUROC for MSI detection and the majority of prediction targets evaluated by sevenfold cross-validation. The cohort, Sample-ID, ground truth and prediction scores for MSI, along with the *TP53* mutational status, a brief pathological evaluation and magnified views of specific areas are provided for in-depth analysis. The heatmaps highlight relevant tumor areas for MSI (middle) and *TP53* mutation (right) prediction. Red signifies high importance and indicates MSI or *TP53* mutation (MUT), while blue signifies low importance and indicates MSS and *TP53* wild type (WT). The color intensity represents the model's attention to that particular area. The heatmaps highlight sections of tumor tissue highly similar for MSI and *TP53.* Nevertheless, corresponding regions in the heatmaps result in divergent scores for MSI (red) and *TP53* (blue). In certain cases, different regions seem to be of higher relevance for the forecasts of MSI and *TP53*, respectively. **A.** The heatmaps highlight tumor, rather than stroma, for MSI prediction, which is plausible and reassuring. Typical morphological features of MSI CRCs include sheets of tumor cells (medullary carcinoma) with a high number of tumor-infiltrating lymphocytes (TILs). Similar regions are highlighted for MSI and *TP53* prediction, with a low prediction score indicating *TP53* WT, despite the ground truth being *TP53* MUT. **B.** The tumor appears to be MSS showing typical morphological features for MSS in CRC, including conventional histology with gland formation (NOS, 'not otherwise specified') and necrosis, as well as a low number of TILs. Consequently, the low MSI predictive score looks plausible, but the high *TP53* predictive score contradicts the ground truth, which is *TP53* WT. **C.** Mucinous adenocarcinoma as seen in this sample is a typical morphological feature of MSI CRCs, characterized by over 50% mucinous differentiation with mucin lakes present and only occasional tumor cells. The MSI score is high and *TP53* score is low, both indicating the same as the ground truth. **D.** Higher intensity is observed for MSS in the heatmap. The morphological features are typical of MSS colorectal cancers, despite the ground truth being MSI. The conventional histology includes gland formation (NOS, 'not otherwise specified') and necrosis, as well as a low number of TILs. Therefore, the pathological examination results in a plausible similar outcome to the model, even though it contradicts the ground truth of MSI.

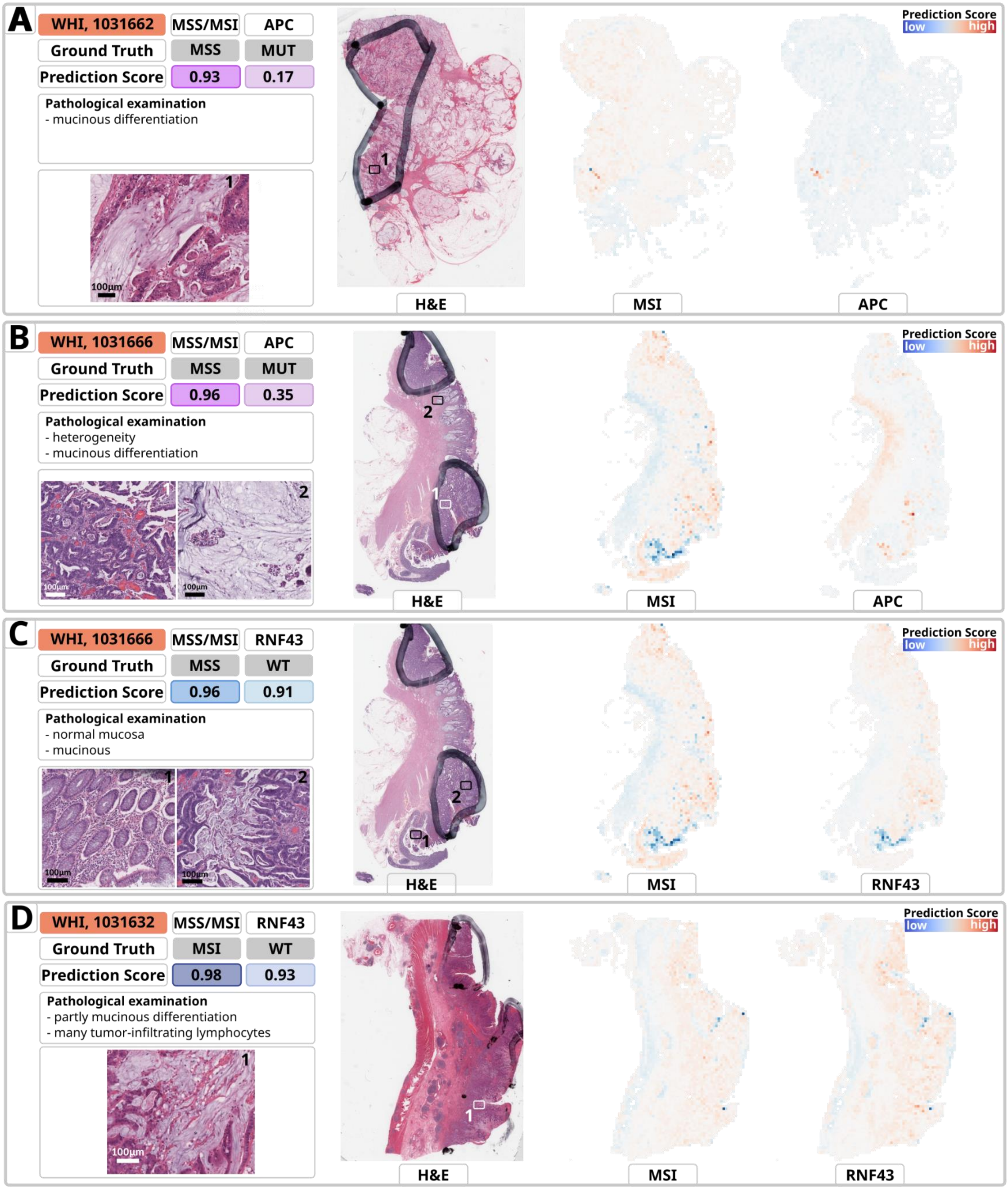

Fig. S10: **Heatmaps of representative samples for prediction of MSI, *APC* and *RNF43* from the external validation dataset**. The heatmaps are derived from the model with the median AUROC for MSI detection and the majority of prediction targets evaluated by sevenfold cross-validation. The cohort, Sample-ID, ground truth and prediction scores for MSI, along with the *APC* (**A**.-**B**.) and *RNF43* (**C**.-**D**.) mutational status, a brief pathological evaluation and magnified views of specific areas are provided for in-depth analysis. The heatmaps highlight relevant tumor areas for MSI (middle) and *APC/RNF43* mutation (right) prediction. Red signifies high importance and indicates MSI or *APC/RNF43* mutation (MUT), while blue signifies low importance and indicates MSS and *APC/RNF43* wild type (WT). The color intensity represents the model's attention to that particular area. All heatmaps highlight relatively concentrated sections of tumor tissue and are highly similar for MSI and *APC/RNF43*, respectively. However, while not only the regions but also the colors representing the scores are highly similar for MSI and *RNF43*, corresponding regions in the heatmaps for MSI and *APC* result in divergent colors and consequently also prediction scores. **A.-B.** The slide shows a case of mucinous adenocarcinoma with a rather heterogeneous appearance in B. The MSI score is high, despite the ground truth being MSS. On the other hand, the score for APC is low, indicating APC WT, although it is mutated. **C.** This case comprises normal mucosa and mucinous differentiation. Although the ground truth is WT, RNF43 is predicted with a high score, implying mutation, and despite being MSS, it receives a high prediction score indicating MSI. **D.** This example shows a case with partial mucinous differentiation and a high number of tumor infiltrating lymphocytes (TILs). The high MSI prediction is pathologically plausible and reflects the ground truth. Despite being WT, RNF43 receives a high score.

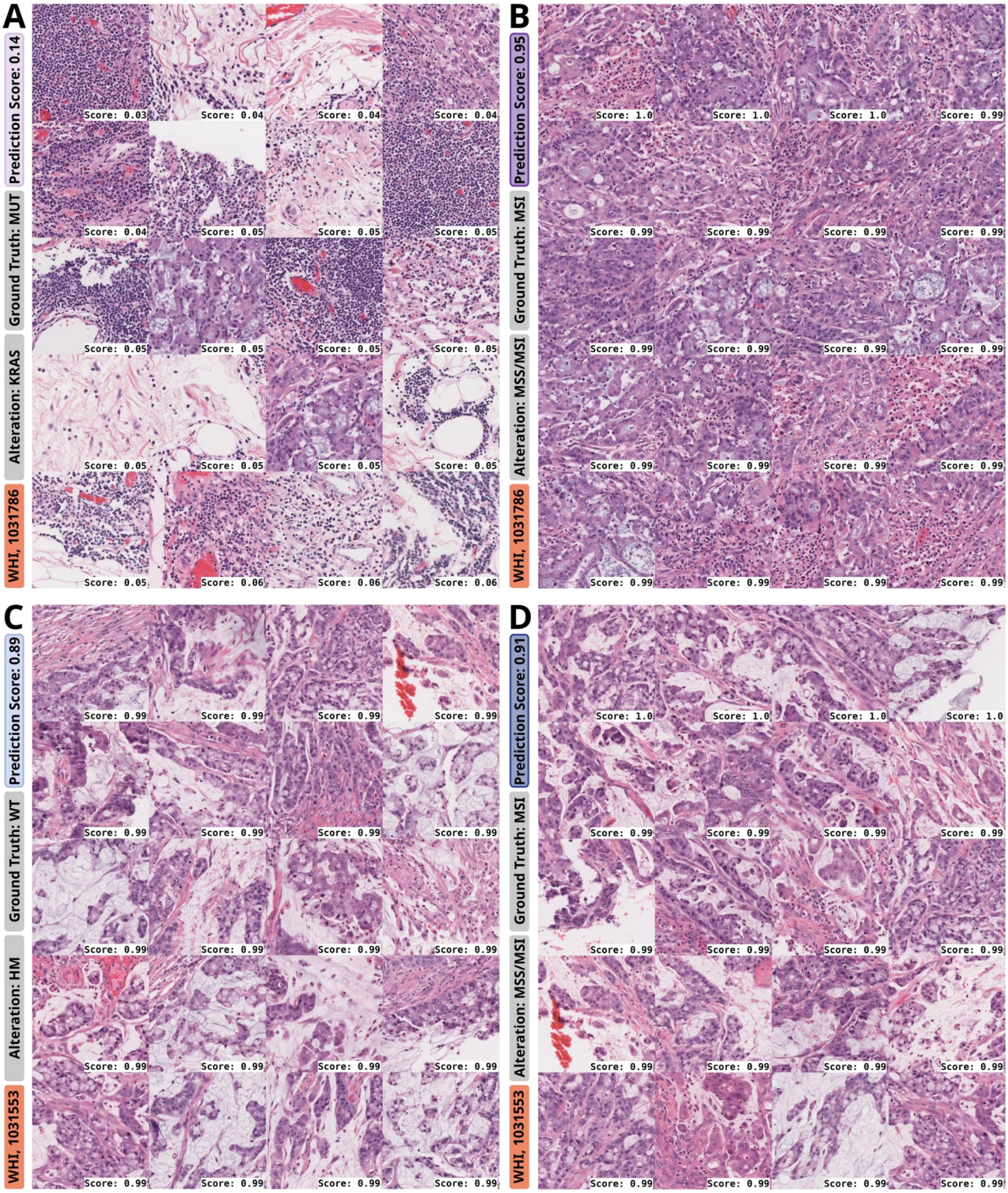

Fig. S11: **Top tiles for prediction of genetic alterations and MSI for two selected slides from Fig. 5.** Each row includes top tiles for one slide (heatmaps in Fig. 5), with tiles for KRAS (**A.**-**B.**)/Hypermutation (HM) (**C.**-**D.**) in the left and MSS/MSI in the right column. A detailed pathological assessment is given in Tab. S19. Abbreviations: HM: Hypermutation; MSI: Microsatellite instability; MSS: Microsatellite stability; MUT: Mutated; WT: Wild type.

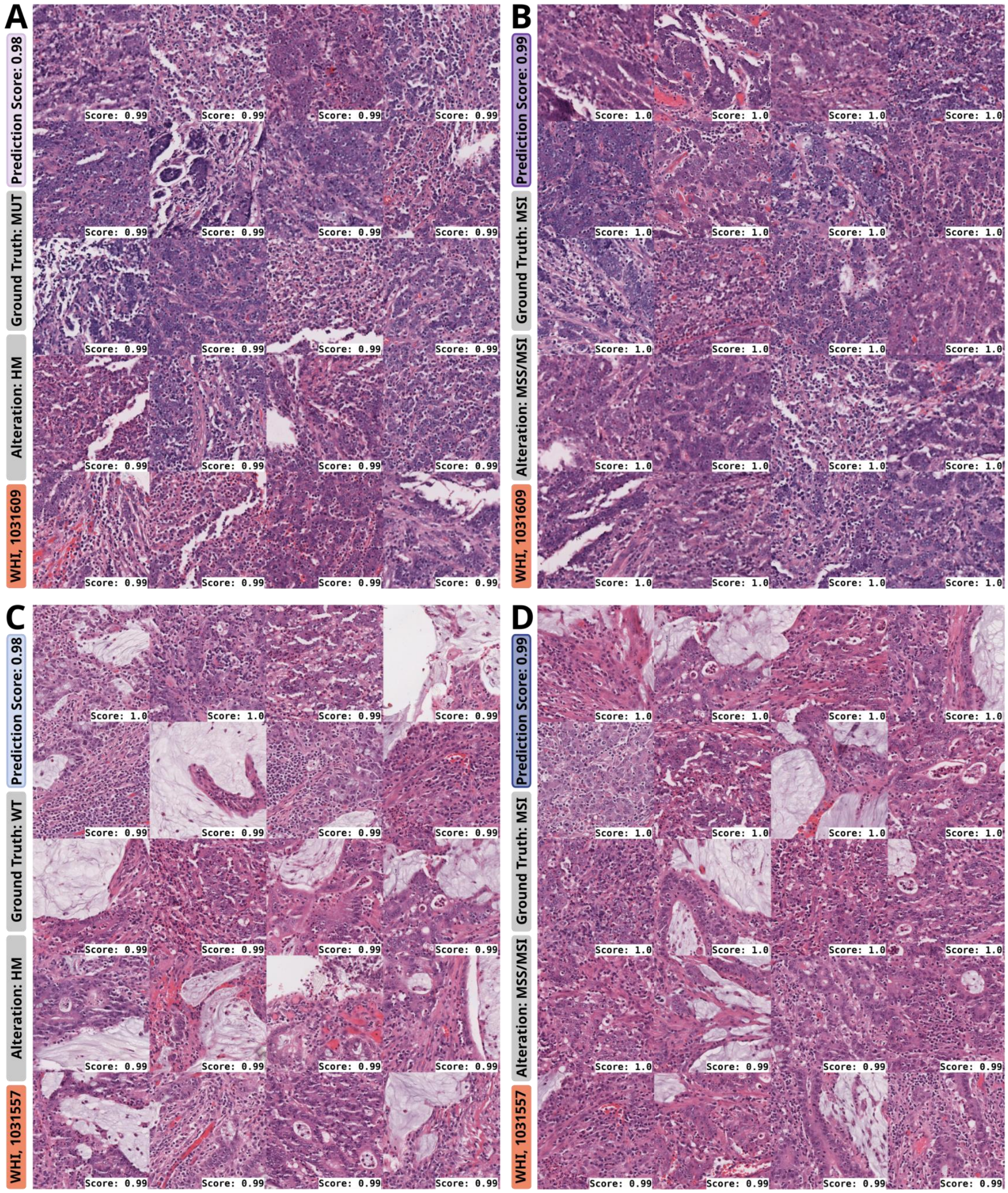

Fig. S12: **Top tiles for prediction of Hypermutation and MSI for two selected slides from Fig. S6.** Each row includes top tiles for one slide (heatmaps in Fig. S6), with tiles for Hypermutation (HM) in the left and MSS/MSI in the right column. A detailed pathological assessment is given in Tab. S19. Abbreviations: HM: Hypermutation; MSI: Microsatellite instability; MSS: Microsatellite stability; MUT: Mutated; WT: Wild type.

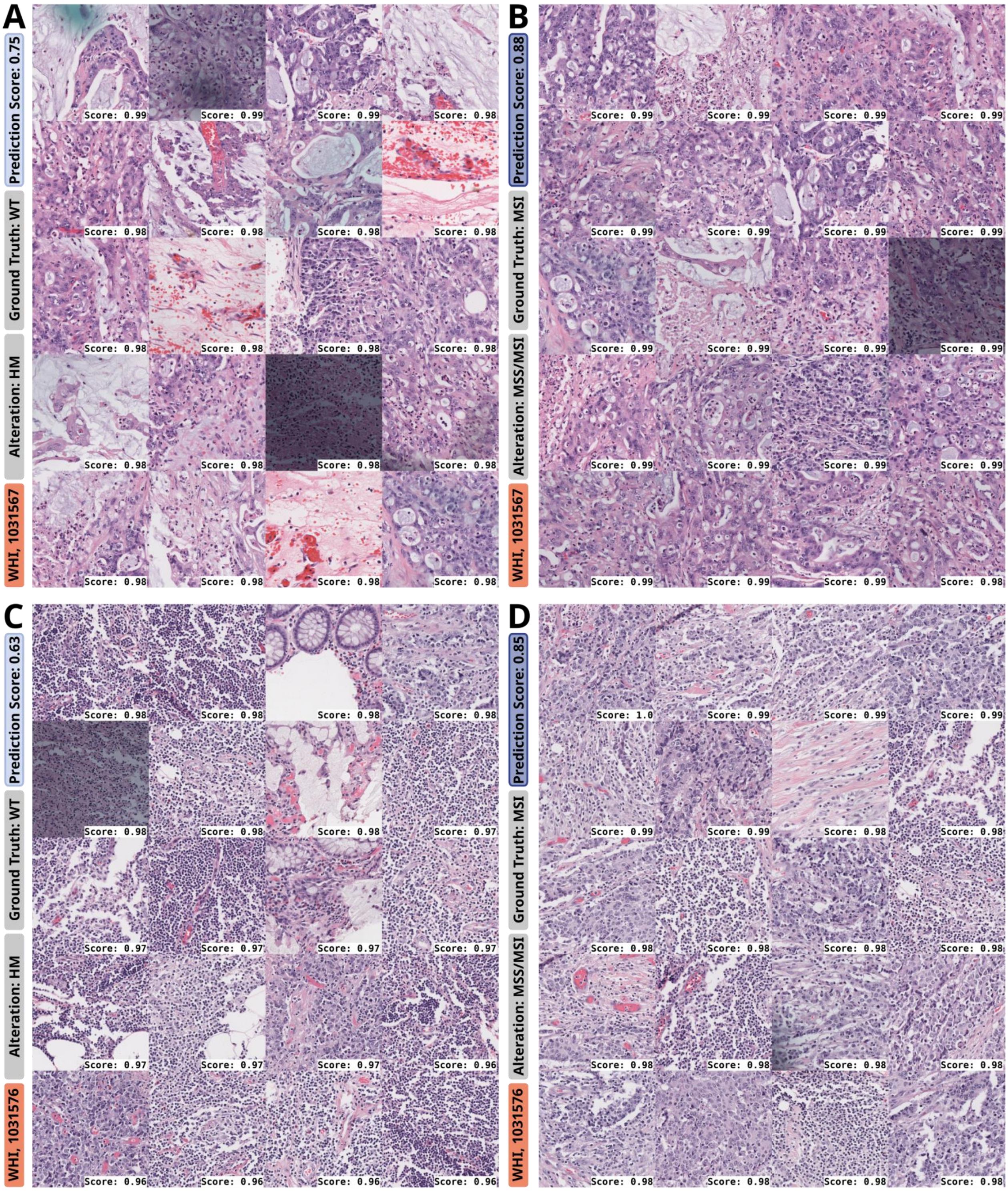

Fig. S13: **Top tiles for prediction of Hypermutation and MSI for two selected slides from Fig. S6.** Each row includes top tiles for one slide (heatmaps in Fig. S6), with tiles for Hypermutation (HM) in the left and MSS/MSI in the right column. A detailed pathological assessment is given in Tab. S19. Abbreviations: HM: Hypermutation; MSI: Microsatellite instability; MSS: Microsatellite stability; MUT: Mutated; WT: Wild type.

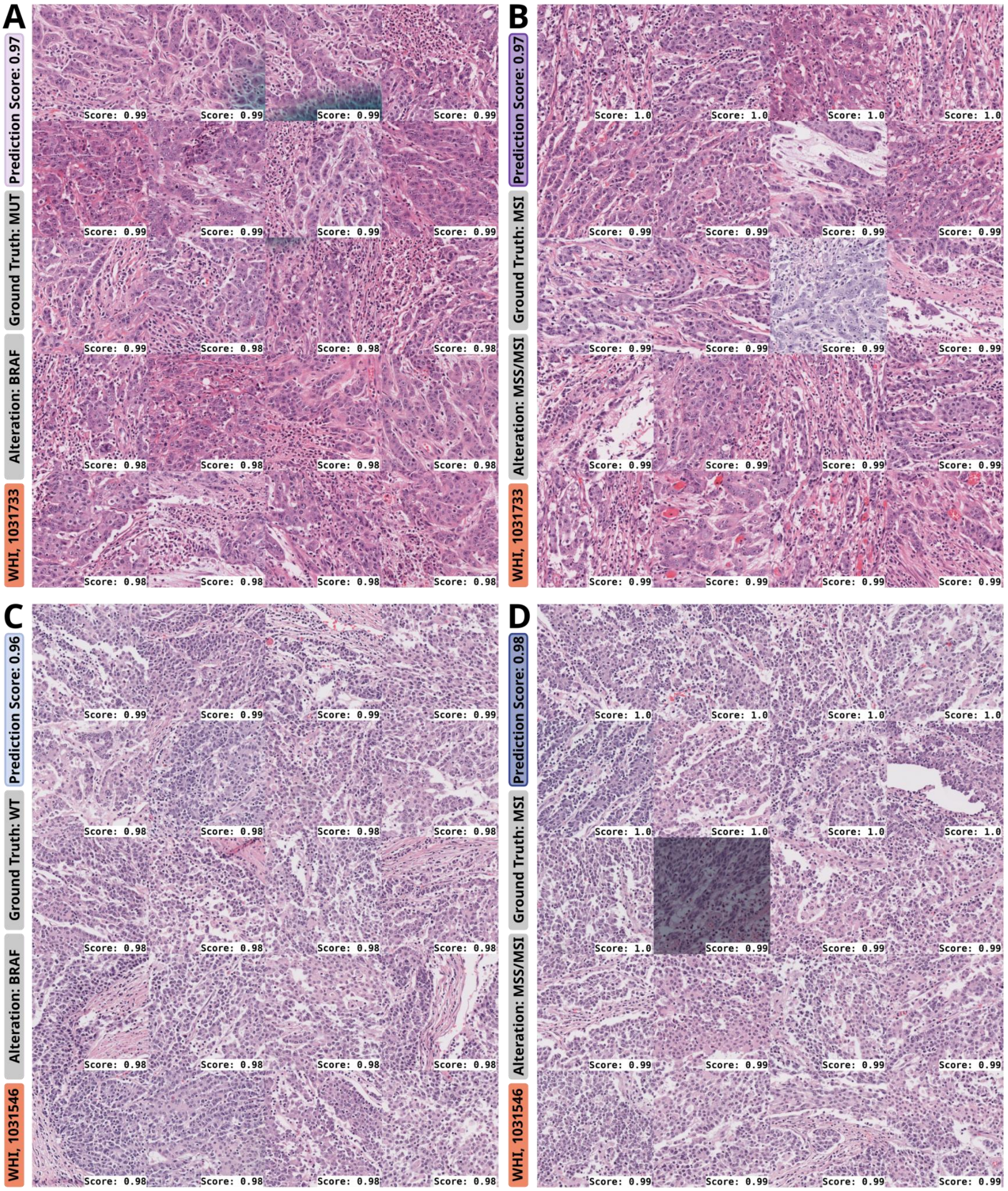

Fig. S14: **Top tiles for prediction of *BRAF* and MSI for two selected slides from Fig. S7.** Each row includes top tiles for one slide (heatmaps in Fig. S7), with tiles for *BRAF* in the left and MSS/MSI in the right column. A detailed pathological assessment is given in Tab. S19. Abbreviations: MSI: Microsatellite instability; MSS: Microsatellite stability; MUT: Mutated; WT: Wild type.

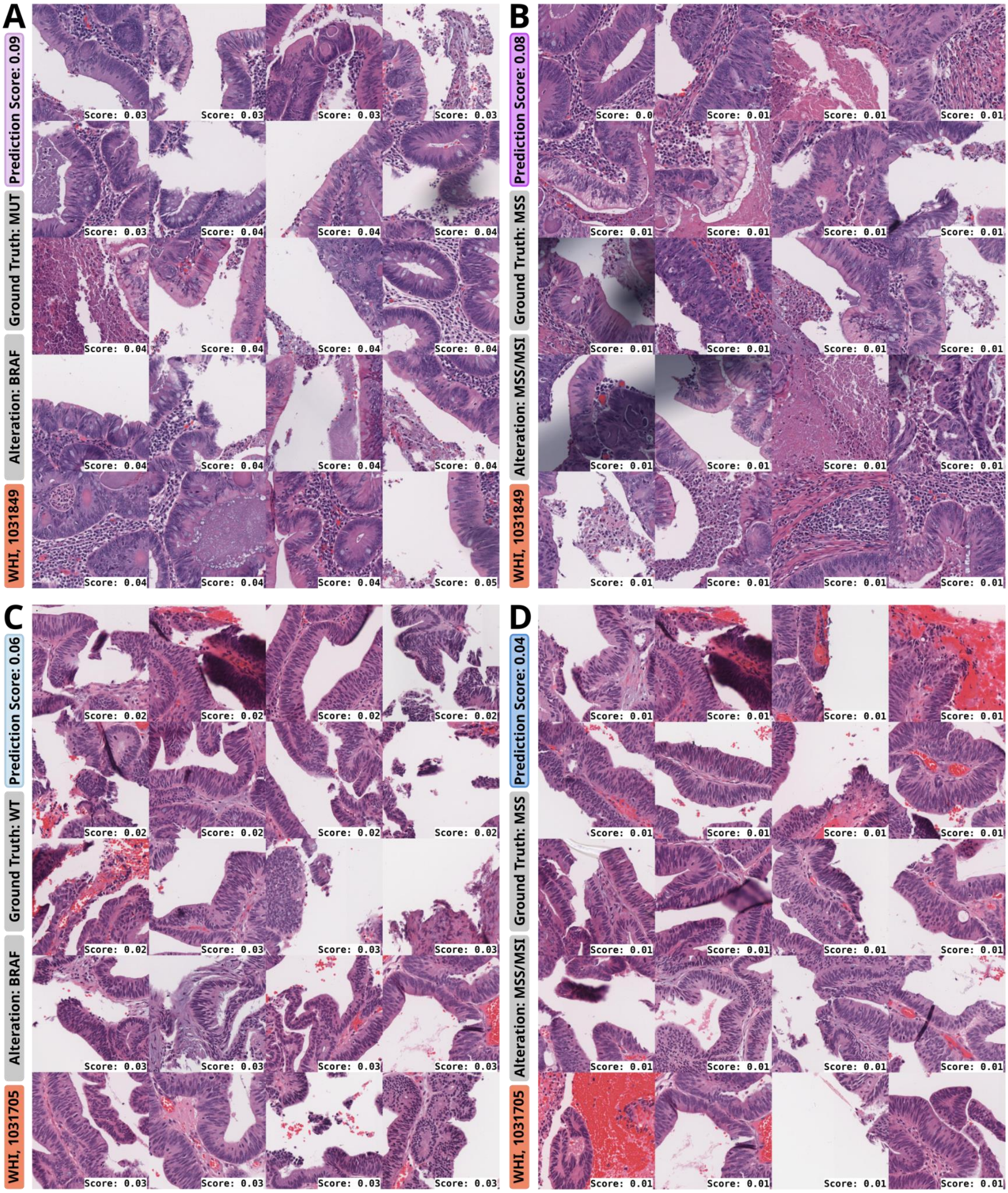

Fig. S15: **Top tiles for prediction of *BRAF* and MSI for two selected slides from Fig. S7.** Each row includes top tiles for one slide (heatmaps in Fig. S7), with tiles for *BRAF* in the left and MSS/MSI in the right column. A detailed pathological assessment is given in Tab. S19. Abbreviations: MSI: Microsatellite instability; MSS: Microsatellite stability; MUT: Mutated; WT: Wild type.

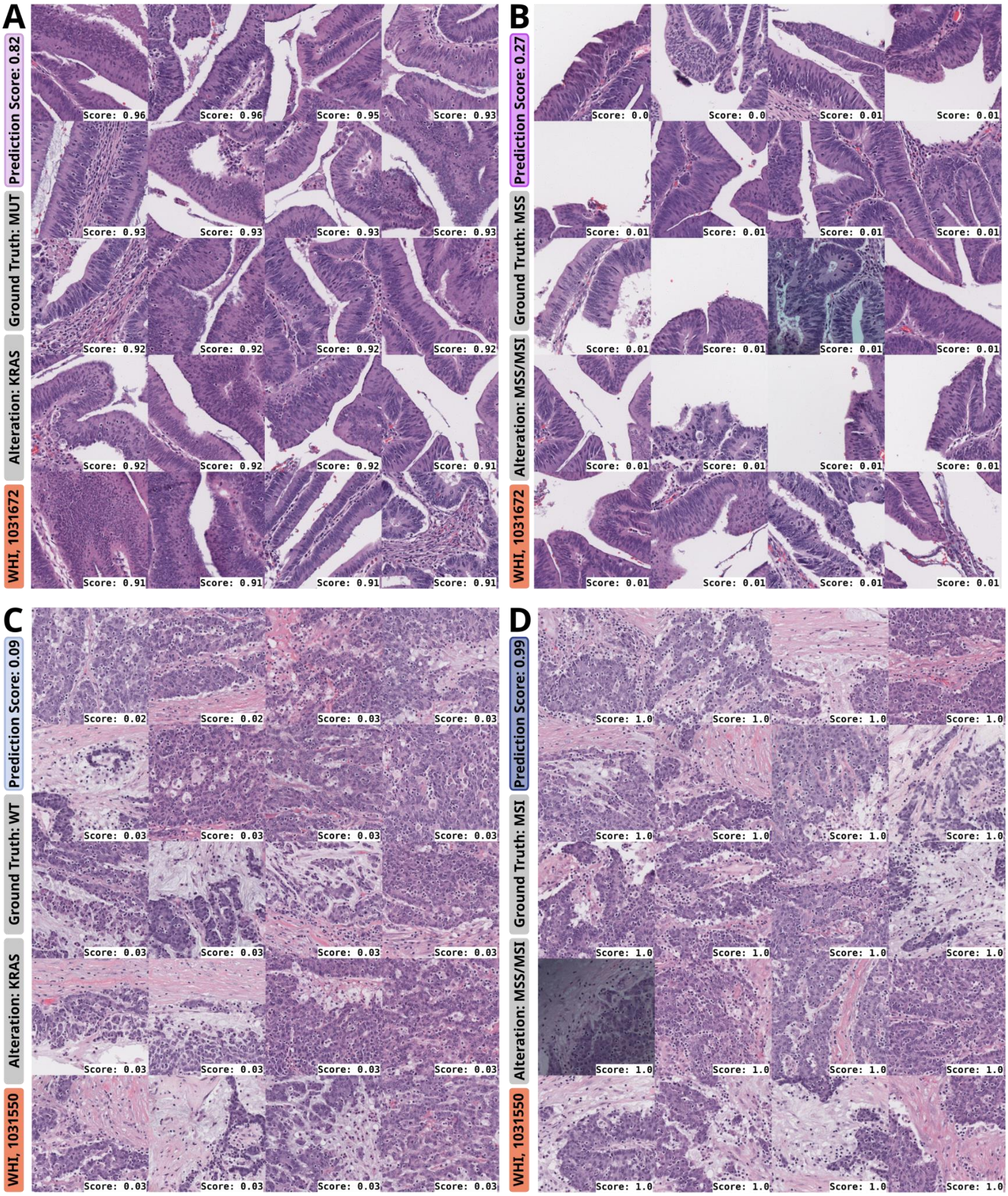

Fig. S16: **Top tiles for prediction of *KRAS* and MSI for two selected slides from Fig. S8.** Each row includes top tiles for one slide (heatmaps in Fig. S8), with tiles for *KRAS* in the left and MSS/MSI in the right column. A detailed pathological assessment is given in Tab. S19. Abbreviations: MSI: Microsatellite instability; MSS: Microsatellite stability; MUT: Mutated; WT: Wild type.

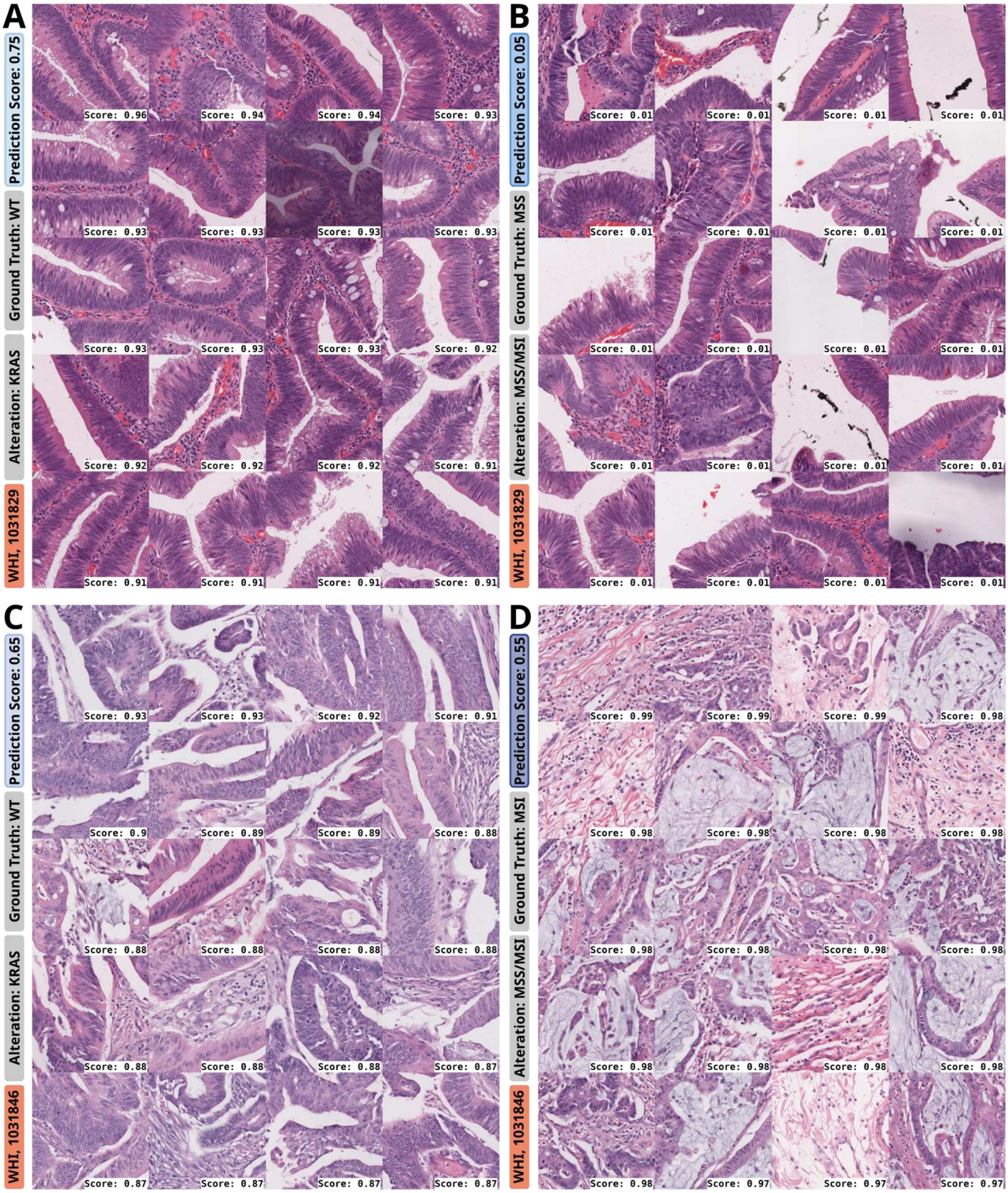

Fig. S17: **Top tiles for prediction of *KRAS* and MSI for two selected slides from Fig. S8.** Each row includes top tiles for one slide (heatmaps in Fig. S8), with tiles for *KRAS* in the left and MSS/MSI in the right column. A detailed pathological assessment is given in Tab. S19. Abbreviations: MSI: Microsatellite instability; MSS: Microsatellite stability; MUT: Mutated; WT: Wild type.

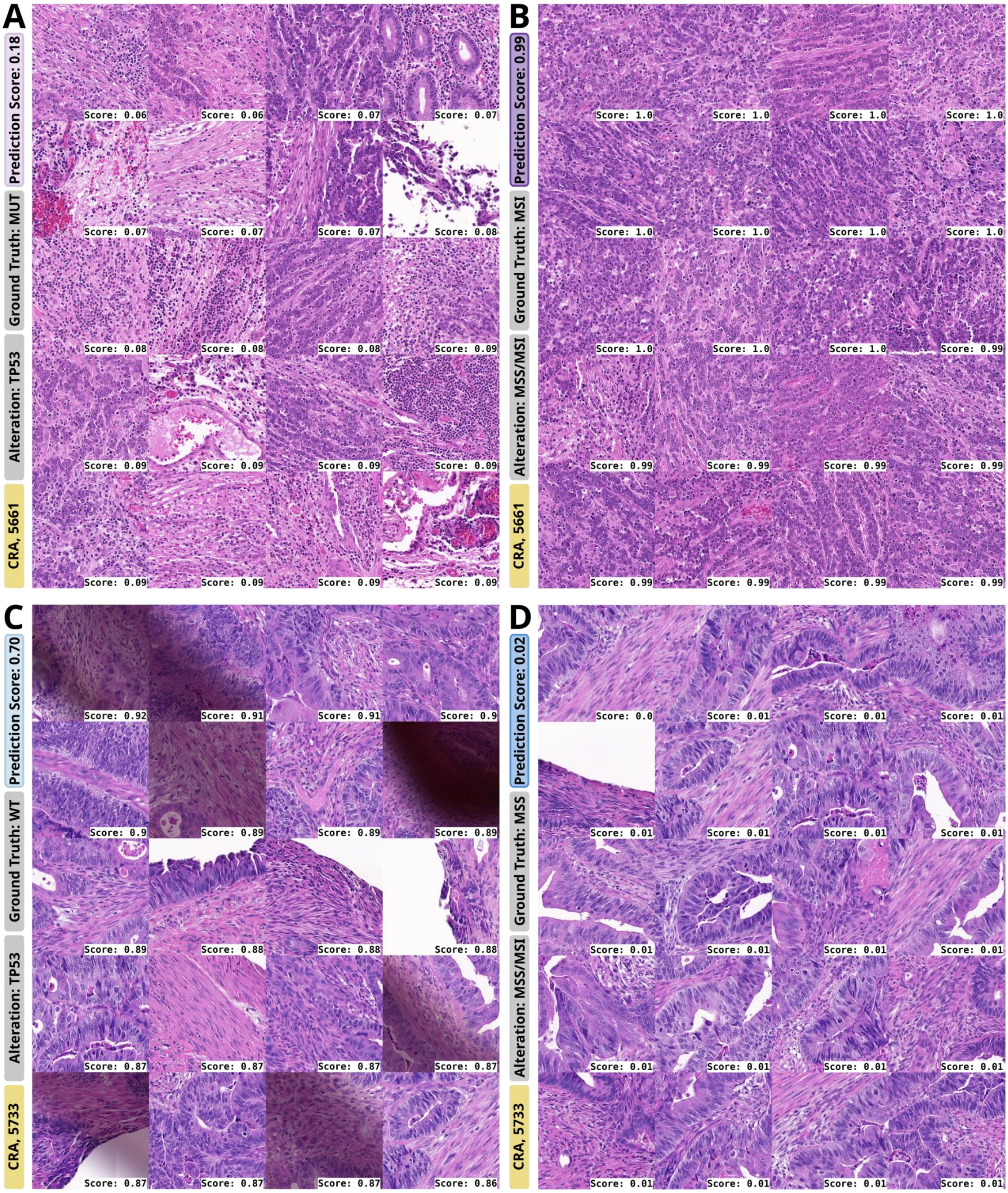

Fig. S18: **Top tiles for prediction of *TP53* and MSI for two selected slides from Fig. S9.** Each row includes top tiles for one slide (heatmaps in Fig. S9), with tiles for *TP53* in the left and MSS/MSI in the right column. A detailed pathological assessment is given in Tab. S19. Abbreviations: MSI: Microsatellite instability; MSS: Microsatellite stability; MUT: Mutated; WT: Wild type.

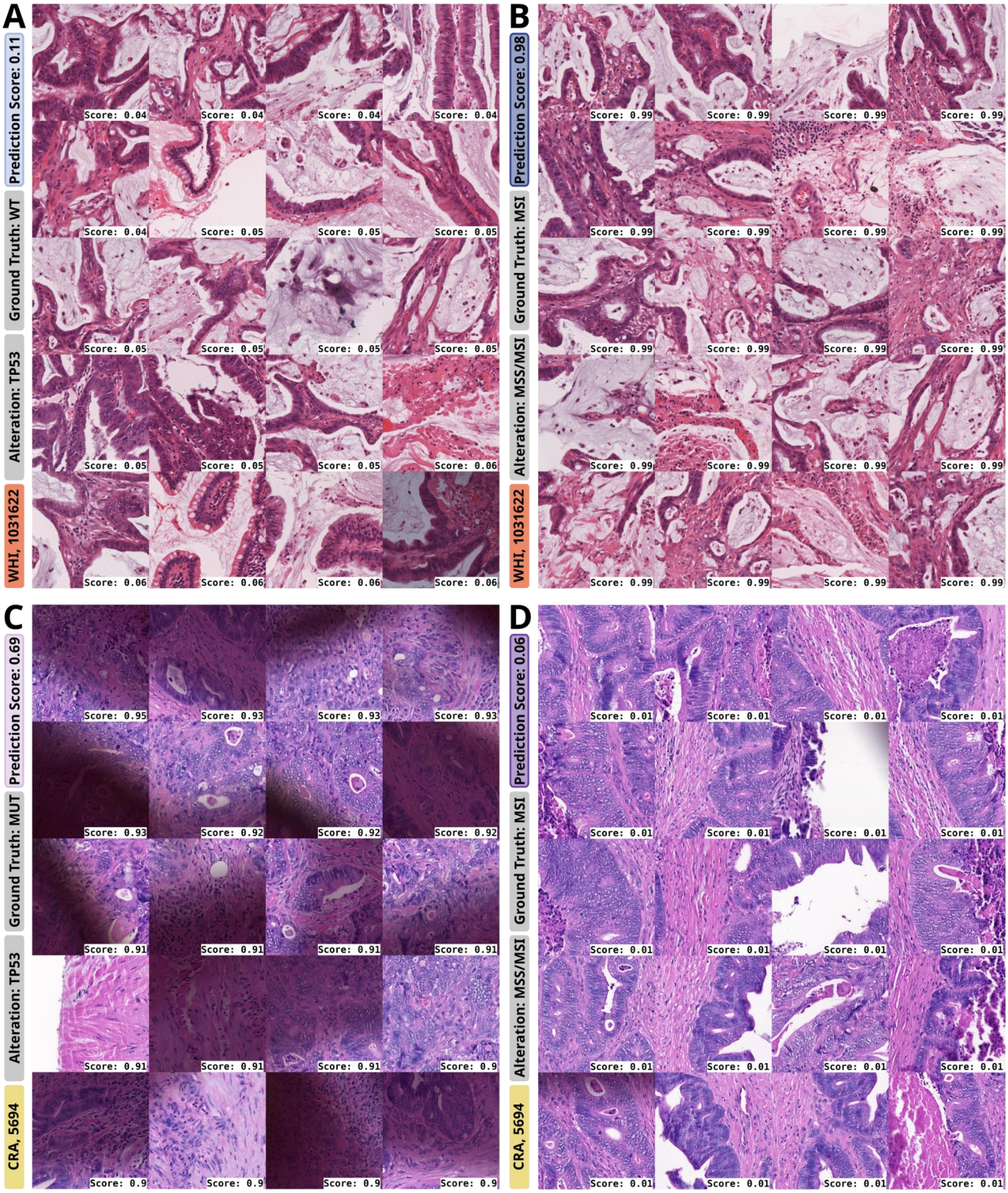

Fig. S19: **Top tiles for prediction of *TP53* and MSI for two selected slides from Fig. S9.** Each row includes top tiles for one slide (heatmaps in Fig. S9), with tiles for *TP53* in the left and MSS/MSI in the right column. A detailed pathological assessment is given in Tab. S19. Abbreviations: MSI: Microsatellite instability; MSS: Microsatellite stability; MUT: Mutated; WT: Wild type.

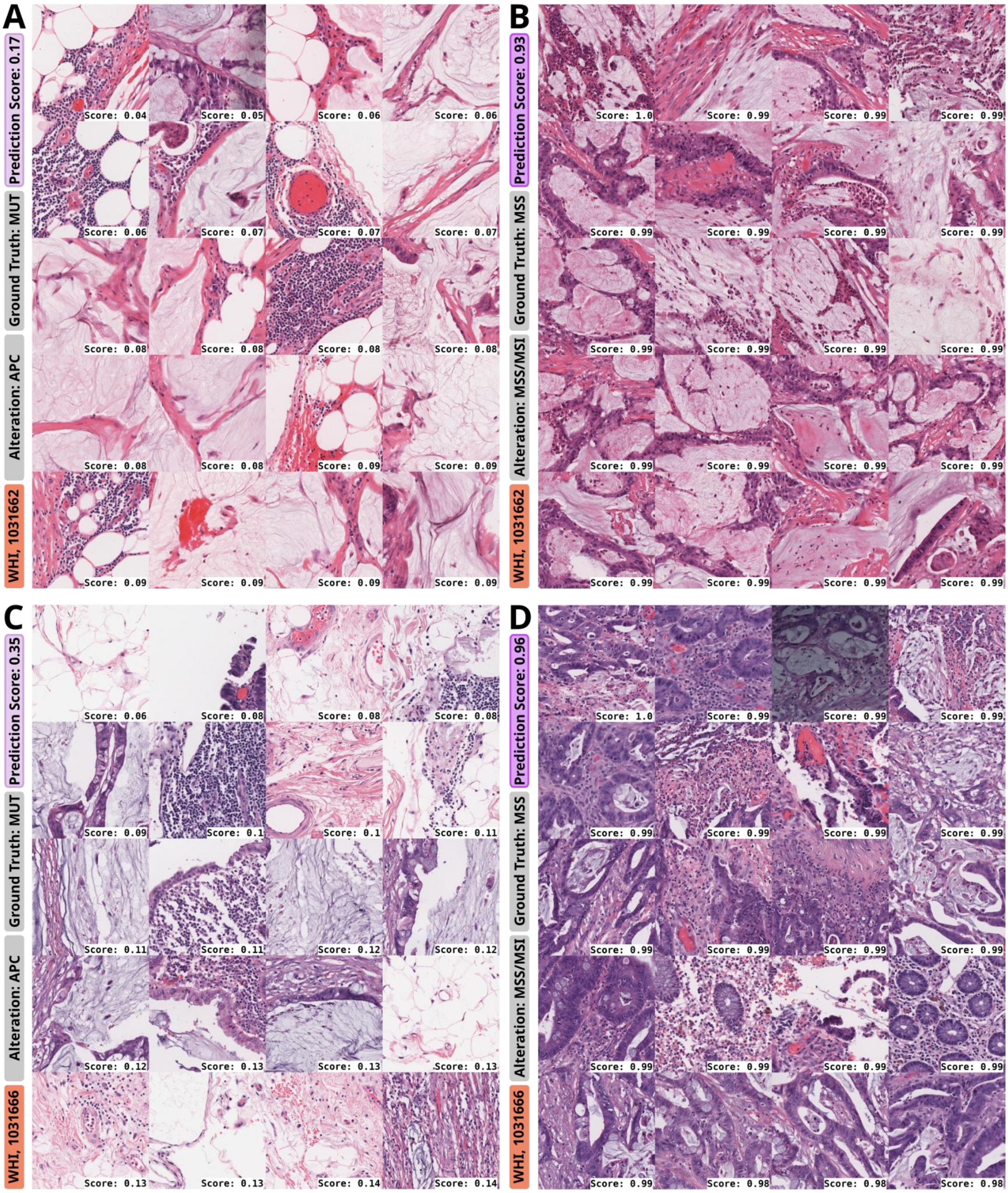

Fig. S20: **Top tiles for prediction of *APC* and MSI for two selected slides from Fig. S10.** Each row includes top tiles for one slide (heatmaps in Fig. S10), with tiles for *APC* in the left and MSS/MSI in the right column. A detailed pathological assessment is given in Tab. S19. Abbreviations: MSI: Microsatellite instability; MSS: Microsatellite stability; MUT: Mutated; WT: Wild type.

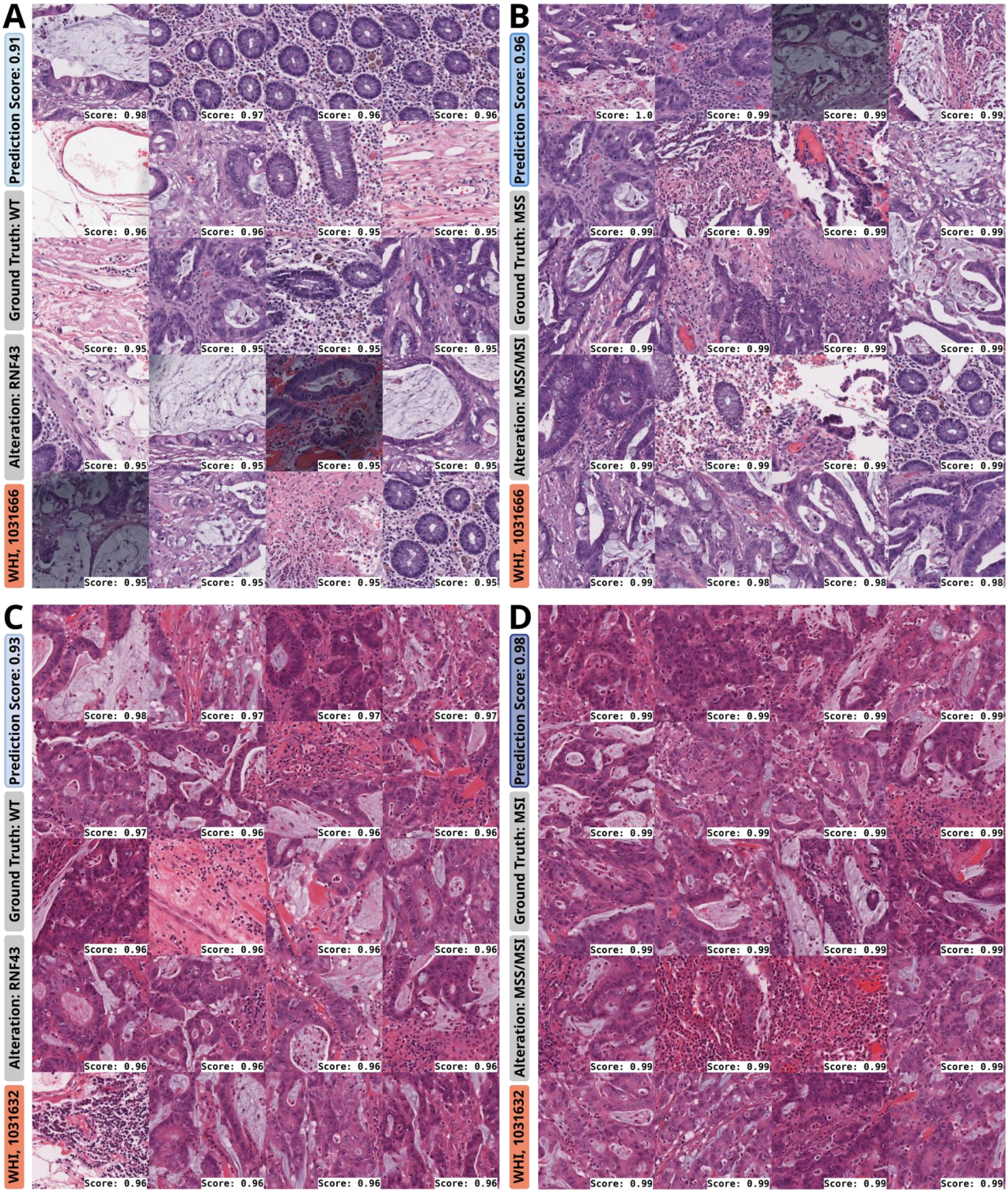

Fig. S21: **Top tiles for prediction of *RNF43* and MSI for two selected slides from Fig. S10.** Each row includes top tiles for one slide (heatmaps in Fig. S10), with tiles for *RNF43* in the left and MSS/MSI in the right column. A detailed pathological assessment is given in Tab. S19. Abbreviations: MSI: Microsatellite instability; MSS: Microsatellite stability; MUT: Mutated; WT: Wild type.

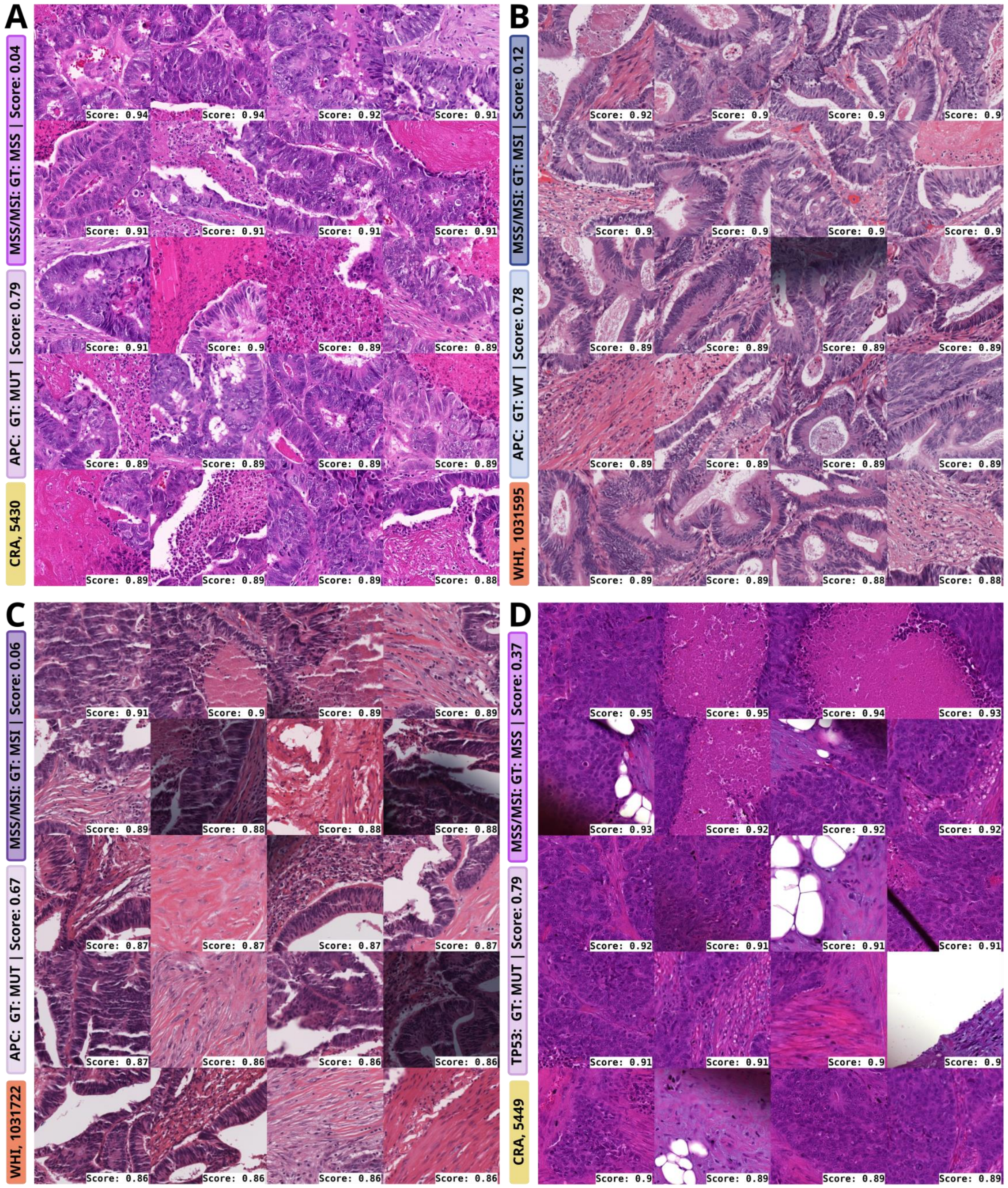

Fig. S22: **Top tiles for prediction of *APC* and *TP53* for selected slides showing tendencies of target specific morphology.** Prediction scores for *APC* and *TP53* seem to be driven by usual gland-forming adenocarcinoma (NOS, 'not otherwise specified') with cribriform architecture which can be broadly summarized as 'MSS-like' morphology. However, especially in **A** and **D**, extensive necrosis is highlighted. In prior studies, abnormal p53 expression as surrogate for *TP53* mutations was not associated with the extent of tumor necrosis in CRC [9] but in other entities such as lung cancer [10] Hence, for example, extensive necrosis could be considered a potentially interesting finding that requires further investigation in the context of the genetic alteration specificity (*TP53* and/or *APC*). However, the morphologic findings here are not distinct. The score for MSS/MSI and the genetic alteration is the prediction score assigned to the slide by the model. The individual tile prediction scores are given with the individual tiles. Abbreviations: GT: Ground Truth; MSI:

Microsatellite instability; MSS: Microsatellite stability; MUT: Mutated; NOS: Not otherwise specified; WT: Wild type.

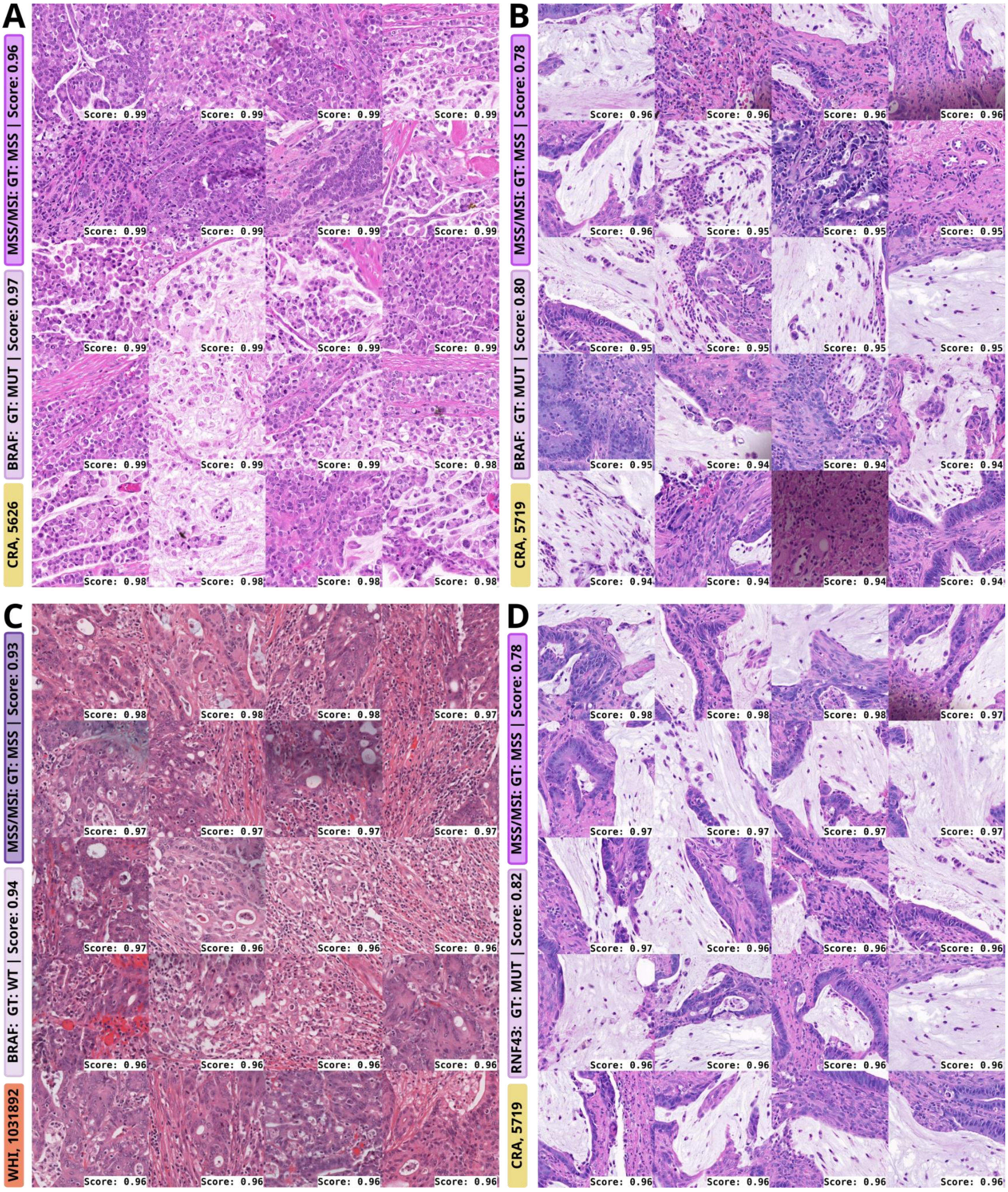

Fig. S23: **Top tiles for prediction of *BRAF* and *RNF43* for selected slides showing tendencies of target specific morphology.** Mucinous morphology and signet-ring cell differentiation displayed in **A** and **B** may be part of a *BRAF* specific morphology [11,12], potentially even beyond MSI, which is also associated with our findings. Moreover, poorly differentiated clusters and tumor budding are highlighted for *BRAF* prediction (**C**). Tumor budding was shown to be associated with *BRAF* mutations [13], and interestingly is inversely correlated with MSI [14] The prediction of *RNF43* MUT seems to be associated with mucinous histology, which is also a typical MSI-like morphology but also further associated with *RNF43* [15] Whether these morphologic findings are alteration specific beyond MSI requires further investigation. The score for MSS/MSI and the genetic alteration is the prediction score assigned to the slide by the model. The individual tile prediction scores are given with the individual tiles. Abbreviations: GT: Ground Truth; MSI: Microsatellite instability; MSS: Microsatellite stability; MUT: Mutated; WT: Wild type.

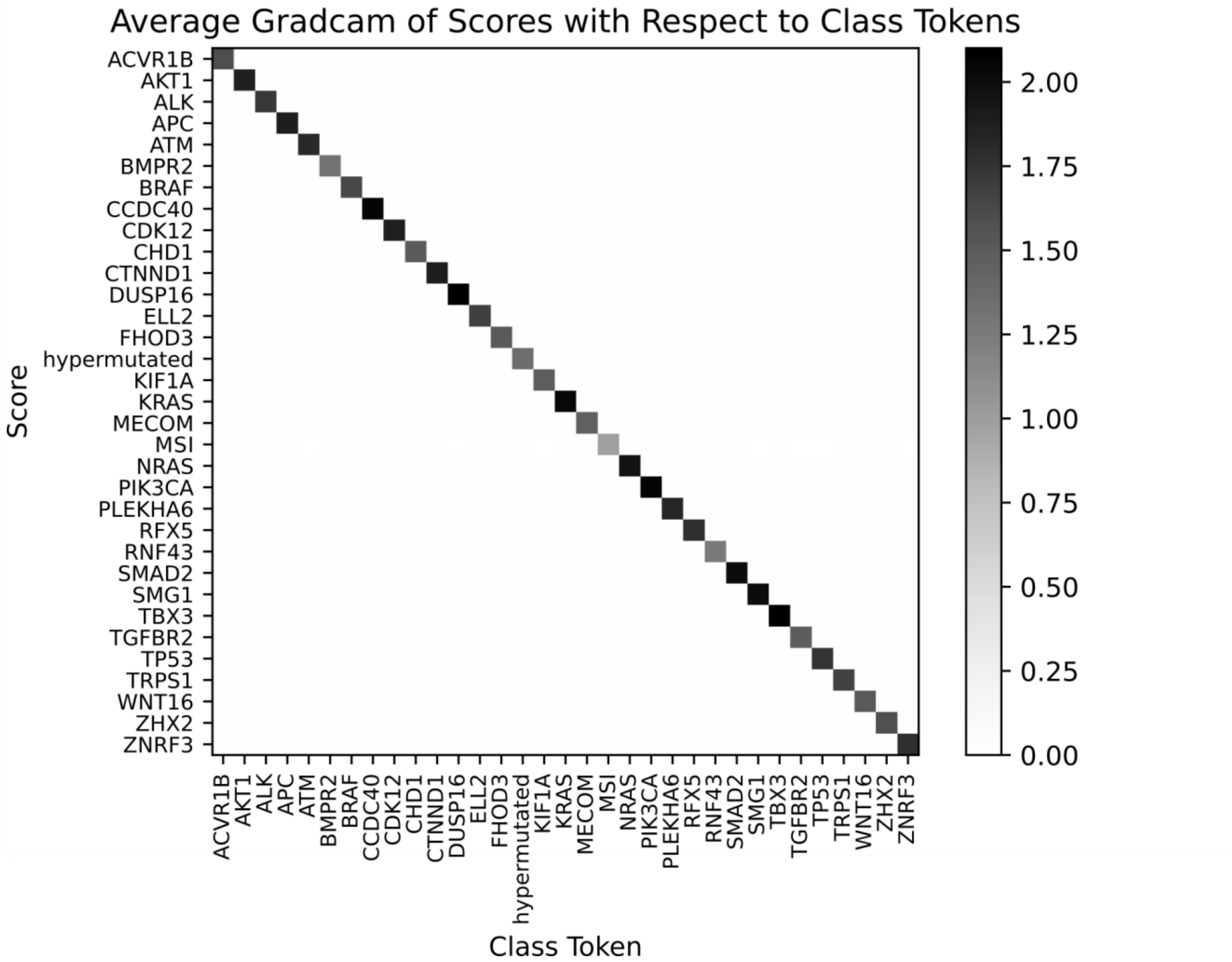

Fig. S24: **Average Grad-CAM of scores with respect to class tokens**: A heatmap illustrating the cross-correlation of Grad-CAM values between scores and class tokens across multiple genetic targets. The results demonstrate that the prediction of each target predominantly relies on its corresponding token in the decoder, indicating a clear token-specific attribution.
